# Activity intensity, volume, and duration and cardiovascular mortality by age, sex, and adiposity

**DOI:** 10.64898/2026.03.25.26349341

**Authors:** Fabian Schwendinger, Denis Infanger, Alex V. Rowlands, Arno Schmidt-Trucksäss

**Author notes:** Corresponding author: Fabian Schwendinger (FS); Grosse Allee 6, 4052 Basel, Switzerland. shared senior authors (Alex V. Rowlands, Arno Schmidt-Trucksäss). Denis Infanger (DI),; Alex V. Rowlands (AVR),; Arno Schmidt-Trucksäss (AST).

## Abstract

**Objective:** To investigate how age, sex, and body morphology modify associations of physical activity (PA) intensity, volume, and duration with cardiovascular disease (CVD) mortality.

**Patients and Methods:** We analysed wrist-worn accelerometer data from 8,607 (51.9% women; 268 CVD deaths) National Health and Nutrition Examination Survey participants. Intensity gradient and average acceleration quantified intensity and volume, respectively. Fine-Gray subdistribution hazards models estimated associations and interactions with age, sex, or waist-to-height ratio.

**Results:** Median (interquartile range) follow-up was 81 (69, 94) months. Subdistribution hazard ratios (SHR) compare 50^th^ with 25^th^ percentile. Relative risk associations were stronger in younger than older adults for intensity (45-year-olds: SHR=0.49, 95%CI:0.40–0.60 vs 75-year-olds: SHR=0.94, 95%CI:0.84–1.06; interaction *P*=.005) and more similar for volume (SHR=0.57, 95%CI:0.41–0.77 vs SHR=0.72, 95%CI:0.59–0.87; interaction *P*=.39). Absolute risk decreased more steeply at lower absolute intensities and more linearly with higher volume and duration in older adults. Evidence for sex differences was limited (interaction *P*=.79–.95), although precision varied by PA dimension. Relative risk associations weakened at higher values of waist-to-height ratio for intensity (interaction *P*=.09) and volume (0.5: SHR=0.51, 95%CI:0.38–0.70 vs 0.7: SHR=0.77, 95%CI:0.62–0.96; interaction *P*=.04). Absolute risk curves differed.

**Conclusion:** Higher PA intensity may be relevant across ages, with greater absolute risk reductions at lower absolute intensities among older adults despite weaker relative associations. Active older adults may benefit more from additional volume than higher intensity. Associations differed little by sex and were attenuated and imprecise at high central adiposity, where evidence for dimension-specific recommendations remains limited.

## 1. Introduction

Physical activity (PA) is a central lifestyle factor that can be modified to prevent cardiovascular disease (CVD) and reduce the risk of premature death.^1^ ^2^ Recent cohort studies using wrist-worn accelerometers have shown that the intensity of PA may be a stronger predictor of CVD outcomes than the volume or duration of PA.^1^ ^2^

Studies increasingly use cut-point-free metrics derived directly from raw accelerometer data, rather than relying solely on traditional category-based approaches (e.g., light, moderate, or vigorous PA).^1^ ^3^ These metrics provide a more nuanced picture of PA on continuous scales, enable more consistent comparisons across populations and devices,^4^ and help clarify the distinct roles of intensity, duration, and volume in shaping health outcomes.^1^ ^2^

To inform tailored PA recommendations, it is important to determine not only whether higher-intensity PA is associated with lower CVD mortality independently of other risk factors, but also whether and how this association may differ by age, sex, and markers of body morphology. These factors are associated with differences in cardiovascular health, physiological function, and responses to PA.^5^ They may therefore influence the extent to which adults benefit from different PA characteristics. Previous research, for instance, suggests mortality may be more strongly associated with PA intensity in healthier and/or younger populations, and with PA volume in less healthy and/or older populations.^6^ ^7^ However, evidence regarding potential modification by age, sex, and body morphology remains limited. Understanding these factors can help refine prevention strategies for specific population subgroups, including older adults, women and men, and adults living with overweight or obesity.

Therefore, this study investigated whether and how age, sex, and body morphology modify the association of PA intensity, duration, and volume with CVD mortality risk in the adult US general population. We hypothesised that the association of PA volume with lower CVD mortality would strengthen with age, but the association of PA intensity would weaken with age, that sex would show little modification of PA-related associations, and that adults with higher central adiposity would exhibit weaker associations of PA intensity with CVD mortality risk.

## 2. Methods

This study is reported following the Strengthening the Reporting of Observational Studies in Epidemiology (STROBE) guidelines (Supplemental Table 1).

### 2.1. Data source and population

We included participants aged ≥20 years of the National Health and Nutrition Examination Survey (NHANES) 2011-2014 with valid accelerometer data.^8^ NHANES evaluates the health and nutritional status of non-institutionalised civilian adults in the United States and was approved by the ethics review board of the National Center for Health Statistics (#2011-17).^8^ All participants gave written informed consent. Data were collected through interviewer-administered questionnaires and standardised health examinations.^8^

### 2.2. Outcome

Participant IDs were linked to death certificate records from the National Death Index to obtain mortality information up to 2019. The outcome was cardiovascular mortality, defined using ICD-10 codes I00-I09, I11, I13, I20-I51, and I60-I69. All-cause mortality was the secondary outcome. Follow-up time was defined from the date of examination until death or censoring.

### 2.3. Exposure

Participants wore triaxial ActiGraph GT3X+ accelerometers (ActiGraph, Pensacola, FL, USA; range: ±6 gravitational units [*g*]; 80 Hz) on the non-dominant wrist continuously for up to 7 days. Raw data were processed using the R-package GGIR v3.3.9 (Supplemental Table 2).^9-11^ We excluded participants who failed autocalibration (post-calibration error ≥0.05 *g*), had <3 valid days (>16 h wear/day), or lacked wear data for any 15-min period of the 24-h cycle.^2^ PA metrics were derived from the average magnitude of dynamic acceleration corrected for gravity and averaged over 5-s epochs (Euclidean Norm minus 1 *g* [ENMO in m*g*]; Supplemental Figure 3):

- Intensity: The intensity gradient (IG) describes the distribution of PA intensity across the 24-hour period. It is calculated by regressing the logged time spent in 25 m*g* acceleration intervals against the logged midpoint of each intensity bin.^2^ ^3^ Higher values reflect more time spent in higher-intensity activities.^2^ ^3^ Values outside -4.5 to -1.5 were excluded as implausible.^2^
- Volume: Average acceleration (AvAcc), the arithmetic mean 24-h ENMO in m*g*, served as a proxy for PA volume.^3^ Values >100 m*g* were excluded as implausible.^2^
- Duration: Time per 24-h day spent above 40 m*g*.^2^

### 2.4. Covariates

Covariates were chosen based on previous research and a directed acyclic graph (Supplemental Figure 1): age (continuous), sex (female/male), income-to-poverty ratio (continuous), ethnicity (Mexican American, other Hispanic, non-Hispanic White, non-Hispanic Black, other race—including multi-racial), education status (below high school, high school, and above high school), smoking status (never, former, current), and history of major chronic conditions (diabetes [yes/no, borderline=yes], cardiovascular disease [yes/no], and cancer [yes/no]).

We used three continuous body morphology markers with the highest discriminative ability for cardiometabolic risk: waist-to-height ratio (WtH), waist circumference, and body mass index (BMI).^12^ WtH was the primary indicator because it captures central adiposity and discriminates risk best.^12^

Blood pressure was the mean of the last two readings after exclusion of the first. Systolic values <70 mmHg and diastolic values <20 mmHg were set to missing. Mean arterial pressure was calculated as one-third systolic plus two-thirds diastolic pressure.^13^

### 2.5. Statistical analysis

Analyses were performed in R (v4.6.1; R Foundation for Statistical Computing, Vienna, Austria)^14^ using the packages survey (v4.5), rms (v8.1-1), survival (v3.8-6), mice (v3.19.0), and mitools (v2.7).

We used survey-weighted Fine-Gray subdistribution hazards models to estimate the associations of each PA metric with CVD mortality, treating non-CVD deaths (n=579) as competing risk, and to test whether these differed by age, sex, and body morphology. Joint design-based Rao-Scott likelihood ratio tests, run in each imputed dataset and combined with the median-P rule, quantified evidence for each interaction term.^15^ Events per model parameter are in Supplemental Table 5.

Separate models were fitted for PA intensity, volume, and duration. PA metrics, age, waist-to-height ratio, waist circumference, and BMI were modelled using restricted cubic splines with 3 knots (Supplemental Table 3). Association modification was assessed by including restricted interaction terms. Specifically, interactions were specified such that products involving nonlinear terms for both interacting variables were excluded. All models were adjusted for the covariates above. Where not the modifier, age and BMI (except in WtH and waist circumference models) were included as spline covariates. Schoenfeld residual plots showed little evidence of non-proportional hazards. Absolute 5-year CVD mortality risk was derived from the fitted Fine-Gray models as the cumulative incidence function via survfit() on an auxiliary weighted Cox model. These are shown as point predictions without confidence intervals (CI) as the auxiliary model’s standard errors do not account for the complex survey design. Absolute risks refer to the most common covariate categories (women, non-Hispanic White, more than high school education, never smoker, no CVD, cancer, or diabetes), age 50 years, and median continuous covariates, unless the variable was the modifier. Interaction tests refer to the relative scale; absolute risk differences are descriptive.

Dose-response curves range from the survey-weighted 2.5^th^ to 97.5^th^ percentile of each PA metric within each stratum (participants grouped at the midpoints between stratum values), extended to the reference where needed. Curves are relative to an inactive profile (intensity gradient −2.90; average acceleration 14 m*g*)^1^ or, for duration, the 25^th^ percentile (222 min). To visualise joint associations, we selected ages 45, 60, and 75 years, spanning the population most at risk, and WtH values of 0.5, 0.6, and 0.7. A WtH <0.5 indicates a favourable body morphology and lower cardiometabolic risk (upper boundary of tier 0 of the WtH classification).^16^ Values of 0.6 and 0.7 represent progressively higher central adiposity (upper boundary of tier 1 and within tier 2, respectively).^16^

We performed multiple imputation by chained equations (5 imputations, 25 iterations) to handle missing covariate data. We imputed continuous variables using predictive mean matching, binary variables using classification and regression trees, and categorical variables using logistic regression. All estimates were pooled across imputations (Rubin’s rules; absolute risks averaged).

#### 2.5.1. Sensitivity analyses

Sensitivity analyses were performed to reduce the risk of reverse causality and explore the impact of missing covariates:

I. Landmark analysis: we excluded participants with less than 2 years of follow-up.
II. We excluded participants with prevalent CVD.
III. Complete-case analyses included congestive heart failure, coronary heart disease, myocardial infarction, and angina pectoris as separate binary variables rather than combined (main analysis). Mean arterial pressure was not included due to missing data.
IV. We repeated the main analyses without adjusting for mean arterial blood pressure and diabetes status.
V. We repeated the main analyses with all-cause mortality as outcome using survey-weighted Cox proportional hazards models.
VI. For comparison, cause-specific Cox models (treating non-CVD death as censored) were also fitted (Supplemental Table 5).

#### 2.5.2. Translation of findings

The intensity and volume that would result from adding 10 min of brisk walking or 10 min of slow running were predicted following Rowlands et al.^17^. Acceleration values used to reflect brisk walking and slow running were 250 m*g*^18-20^ and 600 m*g*^19^, respectively. For volume, the introduced activities replaced time spent at the participant’s AvAcc as previously reported.^2^ ^21^ New volume = baseline volume + (added minutes / 1440) × (acceleration of added activity − AvAcc).^17^

For intensity, the activities replaced inactive time.^17^ Specifically, time from the lowest acceleration category (0-25 m*g*) was reallocated to the acceleration category corresponding to brisk walking (250-275 m*g*) or slow running, keeping total wear time constant.^17^

## 3. Results

The main analyses included 8,607 participants (51.9% women) with imputed covariate data, of whom 847 died (268 from CVD). Table 1 shows unweighted characteristics. Survey-weighted characteristics are in Supplemental Table 4, participant flow in Supplemental Figure 2, and exposure distributions by CVD status, age, sex and WtH in Supplemental Figures 4 and 5.

**Table 1.**
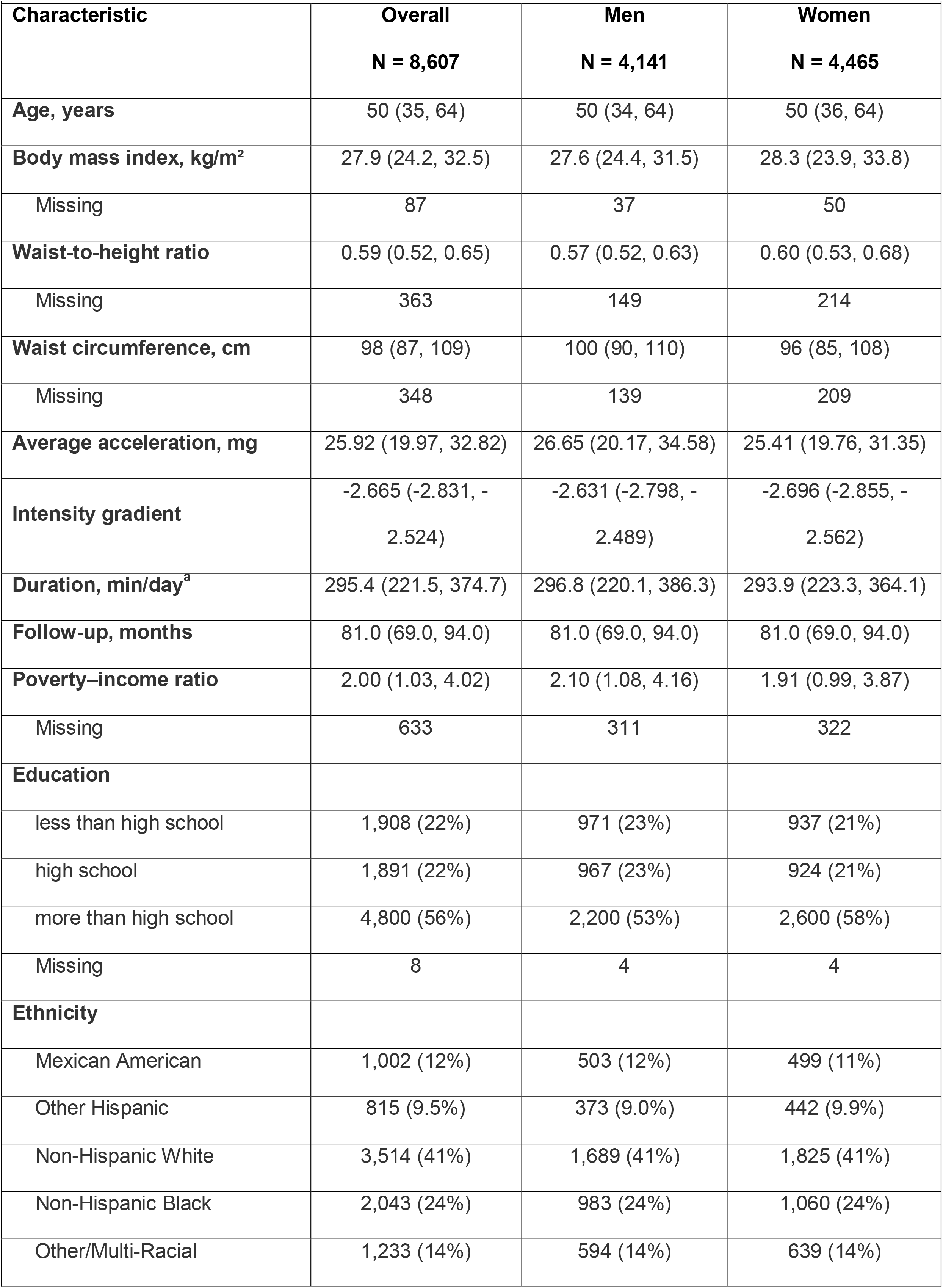
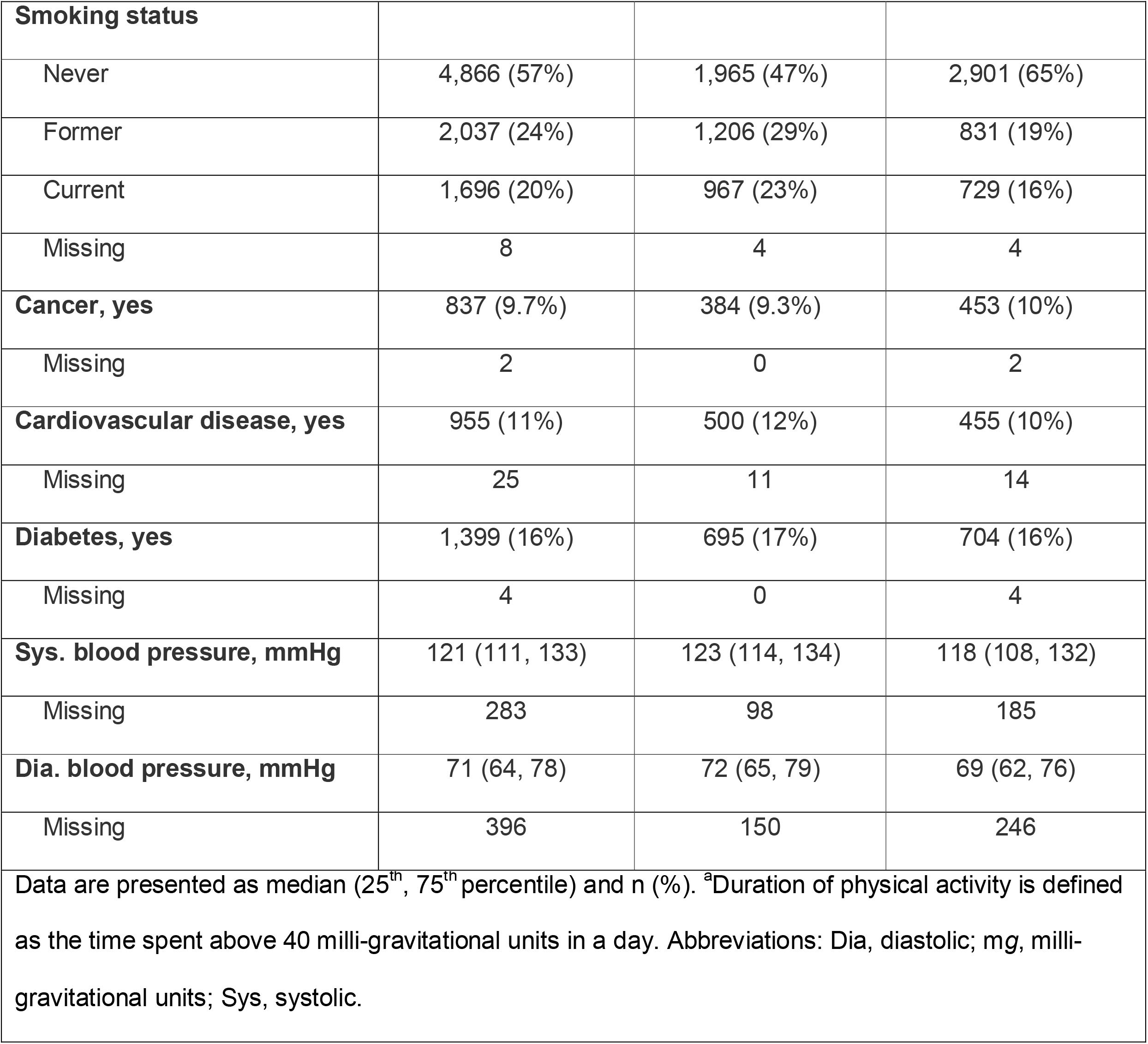
Unweighted cohort characteristics (overall and by sex)

### 3.1. Joint association of physical activity and age with CVD mortality

Risk estimates declined more steeply with higher intensity in younger than in older adults (Fig. 1, Supplemental Figure 6). In older adults, dose-response slopes were flatter and plateaued earlier, with imprecise estimates compatible with decreased and increased risk. The intensity–age interaction showed the strongest evidence of effect modification (*P*=.005). Compared with the 25^th^ intensity percentile, the 50^th^ was associated with 51% (95% CI: 40%–60%) lower risk at age 45, 26% (95% CI: 8%–40%) lower risk at age 60, and 6% (95% CI: −6% to 16%) lower risk at age 75. Corresponding predicted 5-year cumulative risk profiles differed in shape, with the steepest decrease in risk observed at older ages.

**Figure 1.**
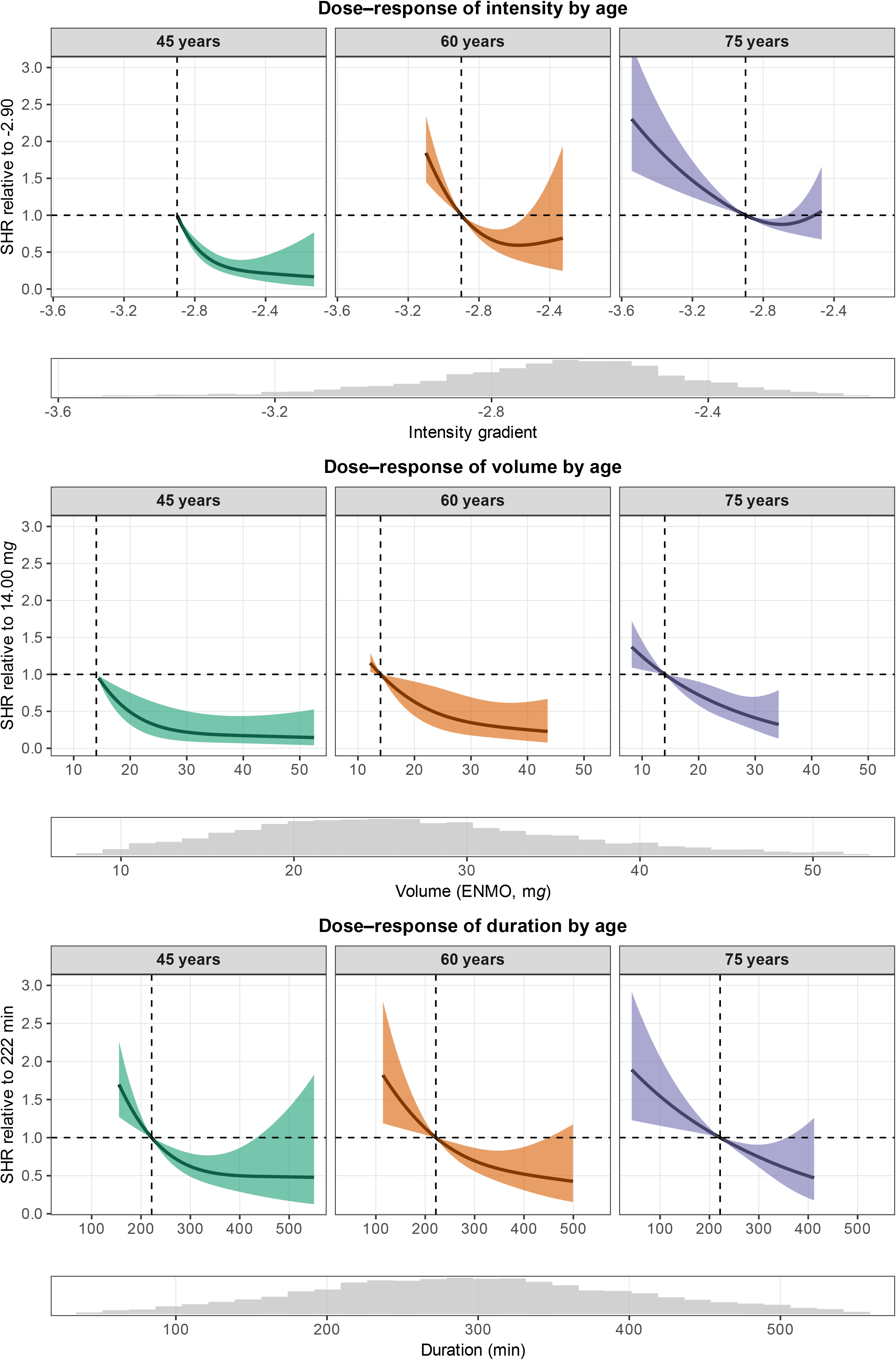
Dose-response relationship between physical activity intensity (top), volume (middle), and duration (bottom) and cardiovascular disease mortality risk, predicted for ages 45, 60, and 75 years. Models included an interaction term between each physical activity metric and age, both modelled continuously using restricted cubic splines, and were adjusted for sex, education, income-to-poverty ratio, ethnicity, smoking status, mean arterial blood pressure, history of cardiovascular disease, cancer history, diabetes, and body mass index. Histograms depict the distribution of the corresponding exposure variable. Estimates at the distribution tails should be interpreted with caution due to sparse data. Abbreviations: ENMO, Euclidean Norm minus 1 *g*; m*g*, milli-gravitational units; SHR, subdistribution hazard ratio.

For volume, visual patterns were more comparable. Higher PA volume was associated with lower risk throughout (Fig. 1, Supplemental Figure 7). In younger and middle-aged adults, the initial decline was steeper but levelled off earlier. In absolute terms, however, differences were larger at older ages, despite limited evidence for a volume–age interaction (*P*=.39).

For duration, patterns were visually comparable (Fig. 1, Supplemental Figure 8). The absolute risk dose-response curve again showed a stronger relationship in older adults, although evidence for a duration–age interaction was limited (*P*=.40). At age 75, the 75^th^ vs 25^th^ percentile was associated with lower risk for volume (SHR 0.48, 95% CI: 0.23–1.01; 1.41% vs 3.47%) and duration (SHR 0.55, 95% CI: 0.28–1.07; 1.82% vs 3.27%) but not intensity (SHR 1.05, 95% CI: 0.75–1.47; 2.81% vs 2.67%).

### 3.2. Joint association of physical activity and sex with CVD mortality

For intensity, dose-response curves were similar in shape but more precise in men than in women, with little evidence for an intensity–sex interaction (*P*=.93; Fig. 2, Supplemental Figure 9, Supplemental Table 5). Compared to the 25^th^ percentile, the 50^th^ was associated with 20% (95% CI: 3%–34%) lower risk for men and 18% (95% CI: −27% to 46%) for women. Absolute risk estimates were higher among men with dose-response patterns comparable to women.

**Figure 2.**
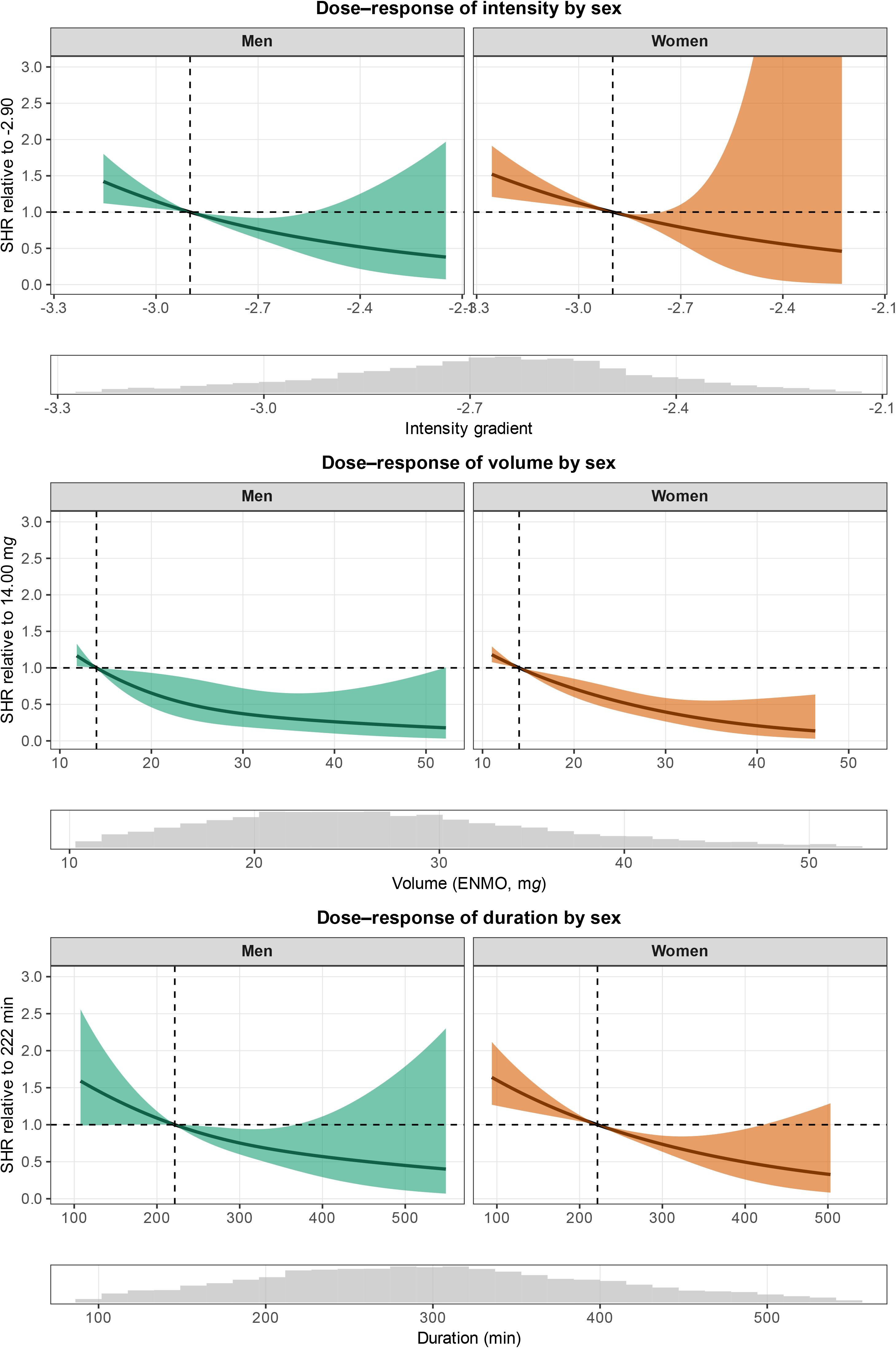
Dose-response relationship between physical activity intensity (top), volume (middle), and duration (bottom) and cardiovascular disease mortality risk, predicted for men and women. Models included an interaction term between each physical activity metric (modelled continuously using restricted cubic splines) and sex, and were adjusted for age, education, income-to-poverty ratio, ethnicity, smoking status, mean arterial blood pressure, history of cardiovascular disease, cancer history, diabetes, and body mass index. Histograms depict the distribution of the corresponding exposure variable in the whole sample. Estimates at the distribution tails should be interpreted with caution due to sparse data. Abbreviations: ENMO, Euclidean Norm minus 1 *g*; m*g*, milli-gravitational units; SHR, subdistribution hazard ratio.

For volume, risk appeared to decline more steeply from low to moderate volumes and then plateaued in men as opposed to a more linear decrease in women (Fig. 2, Supplemental Figure 9). However, there was limited evidence for volume–sex interaction (*P*=.79).

For duration, dose-response curves were more precise in women. In men, durations beyond approximately 500 min/day were compatible with both risk reduction and risk increase (Fig. 2, Supplemental Figure 9). Evidence for a duration–sex interaction was limited (*P*=.95). Absolute differences were larger in men for all volume and duration but dose-response patterns were similar, reflecting their higher baseline risk compared to women.

### 3.3. Joint association of physical activity and body morphology with CVD mortality

Visually, inverse associations of intensity, volume, and duration with CVD mortality were stronger among adults with lower WtH than among those with high WtH (Fig. 3, Supplemental Figures 10–12, Supplemental Table 5). There was evidence for interaction for intensity (*P*=.09), volume (*P*=.04), and duration (*P*=.04). Absolute risks were higher at WtH 0.7 than 0.5. While the absolute risk dose-response curve appeared somewhat attenuated for intensity. However, more distinct patterns were seen for volume and duration. Adults with low to moderate WtH had a steeper decline followed by a plateau compared to a more steady but attenuated decline in those with high WtH.

**Figure 3.**
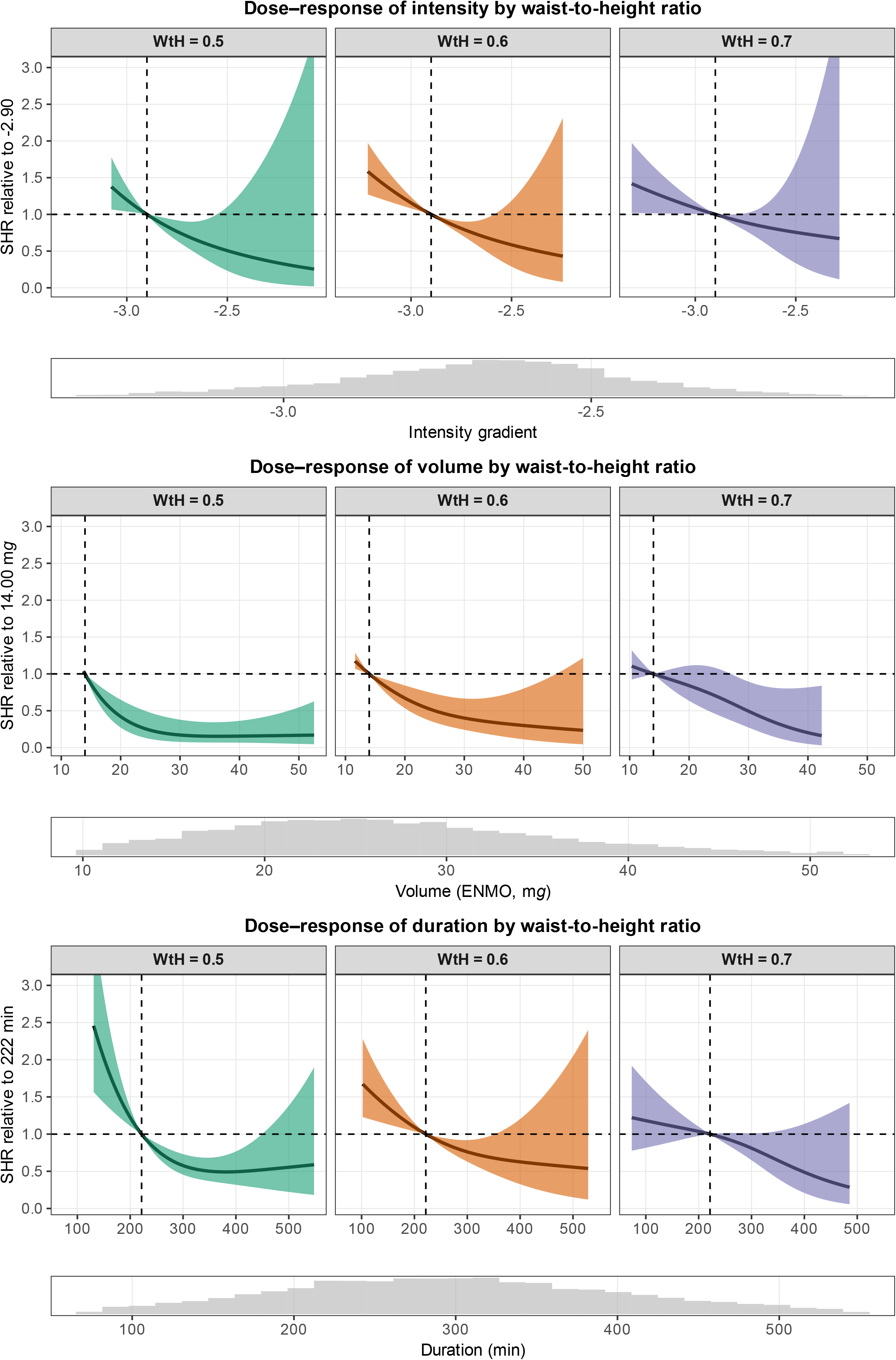
Dose-response relationship between physical activity intensity (top), volume (middle), and duration (bottom) and cardiovascular disease mortality risk, predicted for waist-to-height ratio (WtH) 0.5, 0.6, and 0.7. A waist-to-height ratio <0.5 is considered indicative of a favourable body morphology and lower cardiometabolic risk.^16^ It corresponds to the upper boundary of WtH tier 0.^16^ Values of 0.6 and 0.7 were selected to represent progressively higher levels of central adiposity: 0.6 corresponds to the upper boundary of tier 1, while 0.7 falls within tier 2.^16^ Models included an interaction term between each physical activity metric and WtH, both modelled continuously using restricted cubic splines, and were adjusted for age, sex, education, income-to-poverty ratio, ethnicity, smoking status, mean arterial blood pressure, history of cardiovascular disease, cancer history, and diabetes. Histograms depict the distribution of the corresponding exposure variable. Estimates at the distribution tails should be interpreted with caution due to sparse data. Abbreviations: ENMO, Euclidean Norm minus 1 *g*; m*g*, milli-gravitational units; SHR, subdistribution hazard ratio; WtH, waist-to-height ratio.

Associations were attenuated at larger waist circumference, though less markedly than for WtH (Supplemental Figures 13–16, Supplemental Table 5). Across BMI levels, patterns were somewhat less pronounced than for WtH but still visible (Supplemental Figures 17–20, Supplemental Table 5).

### 3.4. Translating findings into public health messages

Fig. 4 translates the dose-response relationships for PA intensity and volume into comparisons reflecting everyday activity patterns (Supplemental Table 6). These are illustrative comparisons between individuals, not estimates of intervention effects. Among inactive adults (intensity gradient −2.90; 14 m*g*), the intensity model estimated relative risk reductions of 22% (95% CI: 16%–28%), 12% (95% CI: 7%–16%), and 5% (95% CI: 3%–7%) at ages 45, 60, and 75, respectively. Relative reductions also decreased with age in the volume model.

**Figure 4.**
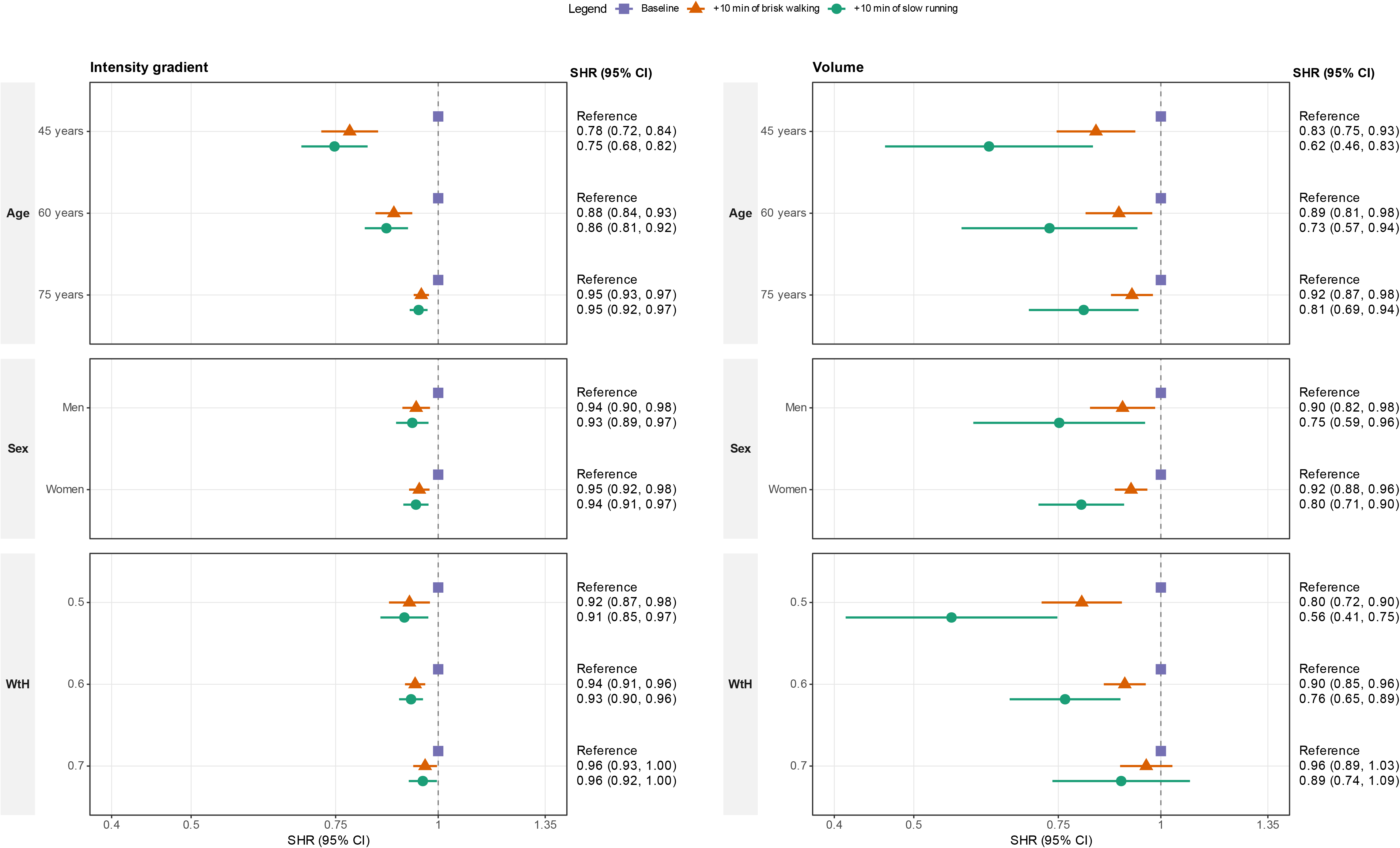
Estimated subdistribution hazard ratios (SHR) for inactive activity profiles across age, sex, and adiposity interactions. This figure summarises SHR scenarios from the dose–response models across selected interactions and strata. For each activity example (inactive, defined as 14 m*g* and -2.9 for volume and intensity, respectively), SHRs are shown for the baseline value, a scenario of adding 10 minutes of brisk walking, and a scenario of adding 10 minutes of slow running. Resulting intensity and volume changes are taken from Rowlands, et al. ^17^. The baseline–inactive combination serves as reference (SHR = 1.0). Abbreviations: CI, confidence interval; m*g*, milli-gravitational units; SHR, subdistribution hazard ratio; WtH, waist-to-height ratio.

Within the inactive group, sex differences were small for intensity (men: 6%, 95% CI: 2%–10%; women: 5%, 95% CI: 2%–8%) and modest for volume (men: 10%, 95% CI: 2%–18%; women: 8%, 95% CI: 4% to 12%).

Differences across body morphology were also modest. Among inactive adults, the intensity model estimated 8% lower risk (95% CI: 2%–13%) at WtH 0.5 vs 6% at WtH 0.6 (95% CI: 4%–9%), with greater differences for volume.

### 3.5. Sensitivity analyses

Complete-case analyses (n = 7,838; events = 226) yielded results consistent with the main findings, with wider 95% CIs (Supplemental Figures 29–33). Similar patterns emerged in the landmark analysis (n = 8,422; events = 216) indicating robustness against reverse causality (Supplemental Figures 21–24). Excluding participants with prevalent CVD (n = 7,650–7,652; events = 131) produced similar but more uncertain results for the age and WtH interaction analyses. CIs narrowed only for women in the sex analyses (Supplemental Figures 25–28). Results for WtH were consistent with the main models (*P*=.01–.03). Results were similar without adjustment for mean arterial pressure and diabetes (Supplemental Figures 34–38). Cause-specific models gave stronger evidence for interactions with adiposity (Supplemental Table 5). For all-cause mortality, dose-response curves were consistent with the results for CVD mortality (*P*=.03–.44; Supplemental Table 7, Supplemental Figures 39–43) and sexes (*P*=.35 to >.99).

## 4. Discussion

In U.S. adults, the relative association between PA intensity and CVD mortality appeared stronger at younger (e.g., ∼45 years) compared to older ages (e.g., ∼75 years). For volume and duration, dose-response patterns were broadly similar across ages but appeared more linear in older adults and showed larger absolute risk differences. We found little evidence for sex-specific differences but noted less precise estimates for the intensity–mortality association in women and duration– mortality association in men. Adults with higher WtH and waist circumference tended to show weaker, less precise PA–mortality associations, whereas patterns by BMI were less clear. This possible subgroup heterogeneity could help tailor PA recommendations for cardiovascular longevity.

### 4.1. Joint association of physical activity and age with CVD mortality

Our findings suggest that the dose-response associations with CVD and all-cause mortality varied across PA dimensions. Whereas the intensity–mortality association plateaued at lower absolute intensities among older adults, the association with volume and duration became more linear with higher age. Among older adults, the flatter, earlier-plateauing dose-response curve for intensity compared to the more linear shapes of volume and duration suggests that inactive adults might benefit from both higher intensity and longer duration. However, those above approximately the population median PA level may gain more from volume and duration than from intensity. Because baseline risk rises steeply with age, the apparently weaker relative associations at older ages translate into substantially steeper declines in absolute risk. This was especially evident for volume and duration, where risk reduction persisted across much of the observed PA range in older adults as opposed to the plateau in younger adults.

This is partly consistent with recent evidence from the Swedish SNAC-K cohort.^6^ Lager et al.^6^ found a stronger but uncertain association between light-intensity walking among ≥80-year-olds compared with 66-year-olds with major adverse cardiovascular events. Evidence for an interaction between age and moderate-to-vigorous PA was inconclusive.^6^ Although point estimates suggested that higher-intensity PA might confer stronger benefits in older adults, 95% CIs encompassed beneficial, null, and harmful associations (HR=0.92, 95% CI: 0.79–1.09).^6^ Unlike cut-point-based moderate-to-vigorous PA, our cut-point-free metrics combined with absolute risk estimates emphasise substantially greater absolute risk reductions at lower PA intensities among older adults. This pattern may be hidden by conventional intensity thresholds and relative risk estimates alone.

Hallmarks of cardiovascular ageing^5^ may explain weaker relative associations at older ages. Mitochondrial dysfunction, cellular senescence, and chronic low-grade inflammation reduce endothelial and myocardial adaptability and promote vascular stiffness.^5^ However, age-related reductions in cardiorespiratory fitness may imply a greater relative physiological stimulus for a given activity. Together with the elevated baseline CVD risk, this may explain why greater absolute risk reductions were observed at lower absolute intensity in older adults despite somewhat weaker relative associations.

### 4.2. Joint association of physical activity and sex with CVD mortality

Chen et al.^22^ found that 250 min/week of device-measured moderate-to-vigorous PA was associated with 30% lower coronary heart disease incidence in women, whereas men required 530 min/week for a similar reduction. In contrast, Zaccardi et al.^21^ found that a daily 10-min brisk walk added to an inactive lifestyle was associated with 1.4 years longer life expectancy in men and 0.9 years in women.^21^ These discrepancies might partly reflect the PA dimensions considered, the cohort characteristics, and the risk scale. Similar relative associations might translate into larger absolute gains in men, whose baseline risk is higher, as in our data.

Our modelled scenario demonstrates how the choice of dimension may shape sex comparisons. The same added PA was associated with similar risk reductions in men and women in the intensity model (6%, 95% CI: 2%–10% vs 5%, 95% CI: 2%–8%), but a larger reduction in men in the volume model (10%, 95% CI: 2%–18% vs 8%, 95% CI: 4% to 12%). Adding 10 min of brisk walking shifts the volume percentile more than the intensity percentile.^17^

Dimension-specific patterns also emerged in the dose-response curves. Estimates were more precise in men for intensity and in women for duration. Excluding prevalent CVD improved precision for women (intensity) but not for men (duration). For volume, risk declined steeply then plateaued in men but declined more steadily in women, despite little evidence of interaction, suggesting that benefits in men may not extend proportionally beyond moderate volume. Our findings might thus temper, rather than resolve, the debate on sex differences by highlighting the role of PA level and dimension.^21^ ^22^ If these patterns reflect true biology, possible explanations include sex differences in cardiovascular regulation, vascular responsiveness, and hormonal milieu,^23^ ^24^ which may further interact with sex differences in cardiovascular ageing.^23^

### 4.3. Joint association of physical activity and body morphology with CVD mortality

Our findings suggest that promoting PA remains important for adults with moderately elevated central adiposity, in whom associations were clearer for volume and duration than for intensity, so accumulating volume through longer durations may be particularly relevant. Among adults with high central adiposity, relative associations were attenuated for all PA dimensions, most clearly for volume, with 95% CIs including 1. However, more linear patterns in absolute risk suggest that, despite weaker associations, more PA duration and volume may still lead to meaningful risk reductions. Promoting PA therefore remains important, although stronger evidence is needed before dimension-specific recommendations can be made.

Device-measured and self-reported PA show differing patterns in how adiposity modifies mortality risk.^25^ ^26^ A meta-analysis of device-measured PA found a stronger inverse association of light-intensity PA with all-cause mortality in adults with normal BMI compared to those living with obesity (BMI: ≥30 kg/m^2^).^26^ For moderate-to-vigorous PA, attenuation was more modest and uncertain.^26^ Notably, when adiposity was defined using waist circumference rather than BMI, associations for moderate-to-vigorous activity were largely similar across groups.^26^

In contrast, longitudinal self-reported data from Taiwan’s MJ Cohort showed lower CVD mortality risk in adults who increased PA volume while reducing waist circumference and BMI than in those who increased PA without changing adiposity.^25^ However, 95% CIs overlapped substantially and little difference was observed when groups were defined using body fat percentage.^25^ Our study adds a complementary perspective by jointly examining intensity, volume, and duration of PA. Our findings suggest that dose-response associations across these PA parameters vary according to adiposity levels and that more research is needed focussing on adults living with high central adiposity.

Research on cardiovascular and inflammatory responses to acute exercise indicates that, beyond higher baseline and post-exercise blood pressure, adults living with obesity experience higher cardiovascular strain,^27^ ^28^ as well as larger increases in inflammatory and oxidative stress markers compared to individuals with normal weight.^29^ These physiological responses might contribute to the attenuated associations between PA and CVD mortality among adults with higher central adiposity.

### 4.4. Study generalisability, strengths, and limitations

We analysed joint associations of PA with age, sex, and body morphology for CVD mortality in a large U.S. population-representative cohort. Analyses of all-cause mortality suggested that some findings may be CVD-specific. While there was still evidence for an interaction with WtH, differences in dose-response patterns across age and sex lessened with limited evidence for formal interaction.

Cut-point-free metrics make results more comparable across populations, wrist-worn accelerometers,^4^ and avoid bias from categorisation as with e.g., moderate-to-vigorous PA.^3^

Despite extensive sensitivity analyses, we cannot completely rule out reverse causality. Moreover, NHANES is an observational study with accelerometer-based PA measured at baseline. We thus do not know whether the measured week reflects habitual PA. However, a recent analysis of up to 4 repeated accelerometer measurements after 3-4 years reported moderate to good reproducibility of PA phenotypes.^30^ Accelerometers may also not accurately capture activities like resistance exercise. Residual confounding is also possible. Lastly, AvAcc and IG are absolute measures of PA.^3^ They thus do not reflect relative intensity or differences in cardiorespiratory fitness.^3^ This likely affects comparisons between ages, sexes, and adiposity measures. Interactions were tested on the relative scale only, and absolute risks refer to one covariate profile without CIs. Finally, the PA metrics were modelled separately and volume and duration were highly correlated, so contrasts between dimensions are not mutually adjusted.

## 5. Conclusion

Higher PA intensity might reduce CVD mortality risk in younger and middle-aged adults and older inactive adults. Regardless of weaker relative associations, older adults showed stronger absolute risk associations at lower absolute intensities. More active older adults might benefit more from accumulating additional volume through longer durations than further increasing intensity. Evidence for sex differences was limited, but dose-response patterns varied by dimension (shape for volume and precision for intensity and duration). Associations were inverse across PA dimensions at a favourable waist-to-height ratio and clearer for volume and duration than intensity at moderately elevated central adiposity. At high central adiposity, relative associations were attenuated and imprecise for all dimensions, yet comparable absolute risk reductions might require greater PA volume.

## Abbreviations

AvAcc: average acceleration
BMI: body mass index
CI: confidence interval
CVD: cardiovascular disease
ENMO: Euclidean norm minus 1 *g*
HR: hazard ratio
IG: intensity gradient
m*g*: milli-gravitational units
NHANES: National Health and Nutrition Examination Survey
PA: physical activity
SHR: subdistribution hazard ratio
WtH: waist-to-height ratio.

## Declarations

### Ethics approval and consent to participate

The National Health and Nutrition Examination Survey was approved by the ethics review board of the National Center for Health Statistics (#2011-17). All participants provided written informed consent prior to study participation.

### Availability of data and materials

The data underlying this article are available on the NHANES website at https://wwwn.cdc.gov/nchs/nhanes/Default.aspx and can be accessed openly. All code supporting this manuscript is publicly accessible on GitHub upon publication: https://github.com/FSchwendinger/pa-age-sex-bmi-cvd.

### CRediT authorship contribution statement

Fabian Schwendinger: Conceptualisation, Data curation, Formal analysis, Methodology, Project administration, Software, Visualisation, Writing – original draft. Denis Infanger: Formal analysis, Validation, Writing – review & editing. Alex V. Rowlands: Supervision, Writing – review & editing. Arno Schmidt-Trucksäss: Supervision, Writing – review & editing. All authors have read and approved the final version of the manuscript, and agree with the order of presentation of the authors. The corresponding author attests that all listed authors had access to the data, meet authorship criteria, and that no others meeting the criteria have been omitted.

## Acknowledgements

Raw accelerometer data processing was performed by FS at sciCORE (http://scicore.unibas.ch/) scientific computing centre at the University of Basel. For the purpose of open access, the authors have applied a Creative Commons Attribution license (CC BY) to any Author Accepted Manuscript version arising from this submission.

## Declaration of generative AI and AI-assisted technologies in the manuscript preparation process

During the preparation of this work, the authors used Claude Opus 5.5 (Anthropic) in order to support language refinement as well as review and correction of the analysis code. After using these tools, the authors reviewed and edited the content as needed and take full responsibility for the content of the published article.

