## Supplementary File 1 for "Activity intensity, volume, and duration and cardiovascular mortality by age, sex, and adiposity"

[§] AVR and AST are joint senior authors.

### Table of contents

**1 Checklist: Strengthening the reporting of observational studies in epidemiology (STROBE) 3**

**2 Directed acyclic graph 7**

**3 Flow of participants 8**

**4 GGIR variable names 10**

**5 Restricted cubic spline placement 11**

**6 Population characteristics 13**

**7 Main analyses 14**

**9 GGIR configuration 72**

**References 77**

### 1 Checklist: Strengthening the reporting of observational studies in epidemiology (STROBE)

See Table 1 for the STROBE checklist.

**TABLE 1** STROBE Statement—Checklist of items that should be included in reports of cohort studies

| | Item No | Recommendation | Page |
|---|---|---|---|
| Title and abstract | 1 | (a) Indicate the study's design with a commonly used term in the title or the abstract | Title; Abstract |
| | | (b) Provide in the abstract an informative and balanced summary of what was done and what was found | Abstract |
| Introduction | | | |
| Background/rationale | 2 | Explain the scientific background and rationale for the investigation being reported | Introduction |
| Objectives | 3 | State specific objectives, including any prespecified hypotheses | Introduction (last paragraph) |
| Methods | | | |
| Study design | 4 | Present key elements of study design early in the paper | Methods 2.1 |
| Setting | 5 | Describe the setting, locations, and relevant dates, including periods of recruitment, exposure, follow-up, and data collection | Methods 2.1–2.2 |
| Participants | 6 | (a) Give the eligibility criteria, and the sources and methods of selection of participants. Describe methods of follow-up | Methods 2.1–2.2 |
| | | (b) For matched studies, give matching criteria and number of exposed and unexposed | n/a |

| | Item No | Recommendation | Page |
|---|---|---|---|
| Variables | 7 | Clearly define all outcomes, exposures, predictors, potential confounders, and effect modifiers. Give diagnostic criteria, if applicable | Methods 2.2–2.4 |
| Data sources/ measurement | 8* | For each variable of interest, give sources of data and details of methods of assessment (measurement). Describe comparability of assessment methods if there is more than one group | Methods 2.3–2.4 |
| Bias | 9 | Describe any efforts to address potential sources of bias | Methods 2.4, 2.5.1 |
| Study size | 10 | Explain how the study size was arrived at | Suppl. Fig. S2 |
| Quantitative variables | 11 | Explain how quantitative variables were handled in the analyses. If applicable, describe which groupings were chosen and why | Methods 2.5 |
| Statistical methods | 12 | (a) Describe all statistical methods, including those used to control for confounding | Methods 2.4–2.5 |
| | | (b) Describe any methods used to examine subgroups and interactions | Methods 2.5 |
| | | (c) Explain how missing data were addressed | Methods 2.5 |
| | | (d) If applicable, explain how loss to follow-up was addressed | Methods 2.2 |
| | | (e) Describe any sensitivity analyses | Methods 2.5.1 |
| Results | | | |

| | Item No | Recommendation | Page |
|---|---|---|---|
| Participants | 13* | (a) Report numbers of individuals at each stage of study—eg numbers potentially eligible, examined for eligibility, confirmed eligible, included in the study, completing follow-up, and analysed | Suppl. Fig. S2 |
| | | (b) Give reasons for non-participation at each stage | Suppl. Fig. S2 |
| | | (c) Consider use of a flow diagram | Suppl. Fig. S2 |
| Descriptive data | 14 | (a) Give characteristics of study participants (eg demographic, clinical, social) and information on exposures and potential confounders | Results 3; Table 1; Suppl. Table S4 |
| | | (b) Indicate number of participants with missing data for each variable of interest | Table 1 |
| | | (c) Summarise follow-up time (eg, average and total amount) | Abstract; Table 1; Suppl. Table S4 |
| Outcome data | 15 | Report numbers of outcome events or summary measures over time | Results 3 |
| Main results | 16 | (a) Give unadjusted estimates and, if applicable, confounder-adjusted estimates and their precision (eg, 95% confidence interval). Make clear which confounders were adjusted for and why they were included | Results 3.1–3.3; Figs. 1–3; Methods 2.4 |
| | | (b) Report category boundaries when continuous variables were categorized | n/a |
| | | (c) If relevant, consider translating estimates of relative risk into absolute risk for a meaningful time period | Results 3.1–3.4; Suppl. Figs. S6–S12 |

| | Item No | Recommendation | Page |
|---|---|---|---|
| Other analyses | 17 | Report other analyses done—eg analyses of subgroups and interactions, and sensitivity analyses | Results 3.5; Supplementary material 1 |
| Discussion | | | |
| Key results | 18 | Summarise key results with reference to study objectives | Discussion (first paragraph) |
| Limitations | 19 | Discuss limitations of the study, taking into account sources of potential bias or imprecision. Discuss both direction and magnitude of any potential bias | Discussion 4.4 |
| Interpretation | 20 | Give a cautious overall interpretation of results considering objectives, limitations, multiplicity of analyses, results from similar studies, and other relevant evidence | Discussion 4.1–4.4 |
| Generalisability | 21 | Discuss the generalisability (external validity) of the study results | Discussion 4.4 |
| Other information | | | |
| Funding | 22 | Give the source of funding and the role of the funders for the present study and, if applicable, for the original study on which the present article is based | Declarations (Funding) |

#### 2 Directed acyclic graph

We constructed a directed acyclic graph to visualise assumed causal relationships between physical activity and cardiovascular disease (CVD) mortality and to identify potential confounding pathways. Based on this structure, covariates were selected to block non-causal backdoor paths between the exposure and outcome (see Figure 1).

The directed acyclic graph identified education,[1] income,[2] ethnicity, smoking status,[3,4] blood pressure, and prevalent diseases (CVD, cancer, diabetes) as potential confounders based on available evidence and subject matter knowledge.[5] Because blood pressure and diabetes may also lie on the causal pathway between physical activity and CVD mortality, we repeated all analyses without adjusting for them (Section 8.4). While fruit and vegetable consumption has been shown to be associated with CVD mortality,[6] we assume no causal pathway to physical activity. Moreover, we assume an effect of age, sex, and body morphology on both physical activity and CVD mortality.[4] While the relationship between body morphology and physical activity may be bidirectional, we considered body morphology primarily as a confounder. Adults with less favourable body morphology generally engage in lower levels of physical activity and experience greater mortality risk.[4,7] Baseline measures are likely to reflect pre-existing overweight or obesity that predates the assessed physical activity exposure. Lastly, we included genetic and environmental factors as potential unobserved confounders in the DAG.[8,9]

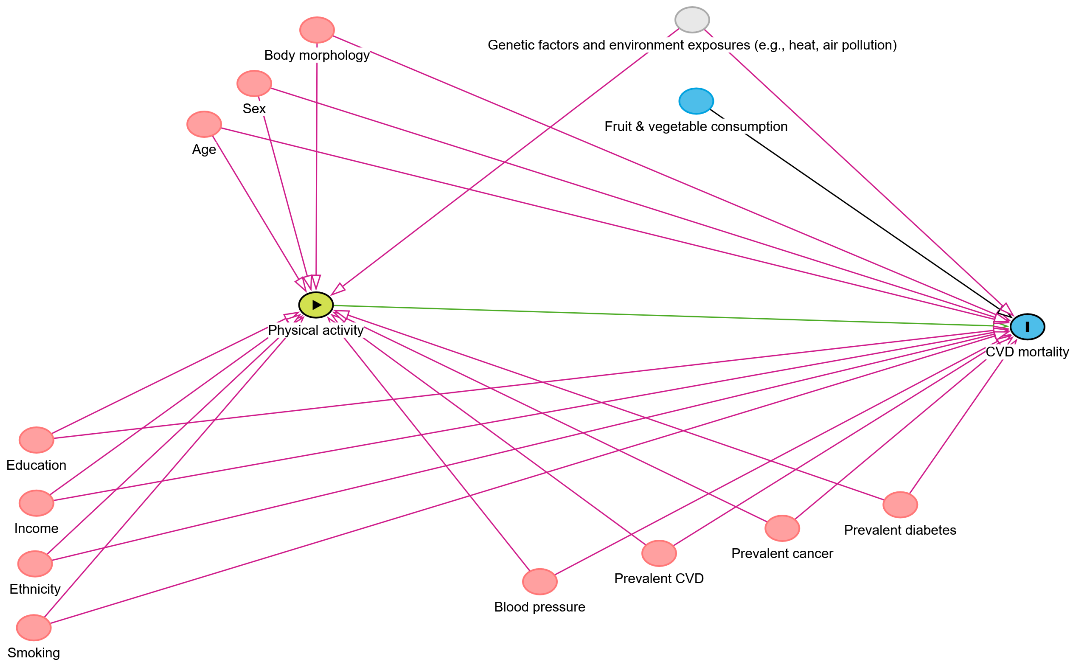

**FIG 1** Directed acyclic graph illustrating assumed causal relationships between physical activity and cardiovascular disease (CVD) mortality. The DAG was used to identify potential confounding pathways and inform covariate selection. Red lines illustrate potential biasing paths. Green reflects the assumed causal path. Grey colour highlights unobserved variables.

### 3 Flow of participants

The flow of participants is shown in Figure 2.

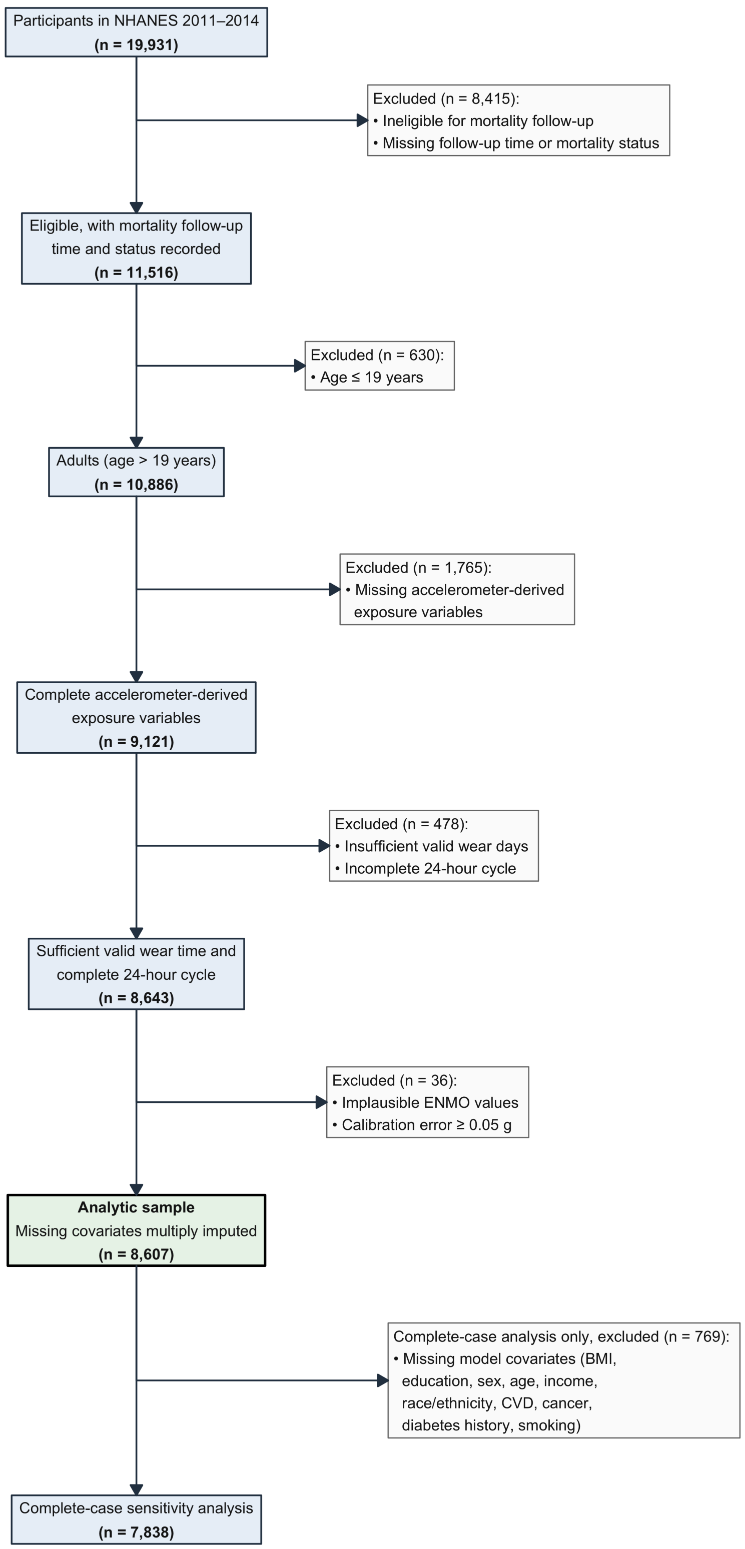

**FIG 2** Flow of participants.

#### 4 GGIR variable names

Table 2 contains the description and names of the variables used in the analyses. All variables are output of GGIR's part 2 person summary .csv file.[10] The intensity gradient and average acceleration are defined as described by Rowlands et al.[11]. The config.csv file to reproduce the GGIR analyses is at the end of the Supplementary file.

**TABLE 2** GGIR variables used in the analyses and corresponding naming in the manuscript.

| Name in manuscript | Name in GGIR | Definition |
|---|---|---|
| Intensity gradient / intensity of physical activity | AD_ig_gradient_ENMO_0.24hr | Intensity distribution of physical activity across a 24-h day. A higher IG indicates less time spent at low intensity and/or more time spent at higher intensity. |
| Intercept | AD_ig_intercept_ENMO_0.24hr | Intercept of the regression line from which the intensity gradient is derived. |
| $R^2$ of intensity gradient regression line | AD_ig_rsquared_ENMO_0.24hr | Describes the model fit of the linear regression used to derive the intensity gradient. |
| Average acceleration / volume of physical activity | AD_mean_ENMO_mg_0.24hr | Arithmetic average of Euclidean Norm Minus One (ENMO) across a 24-h day in m*g* (proxy for physical activity volume). |
| Duration of physical activity | AD_MVPA_E5S_T40_ENMO_0.24hr | Time spent above 40 m*g* in a 24-h day, averaged across all valid days. |

Figure 3 illustrates the concepts of the cut-point-free metrics used in this study: intensity gradient and average acceleration.

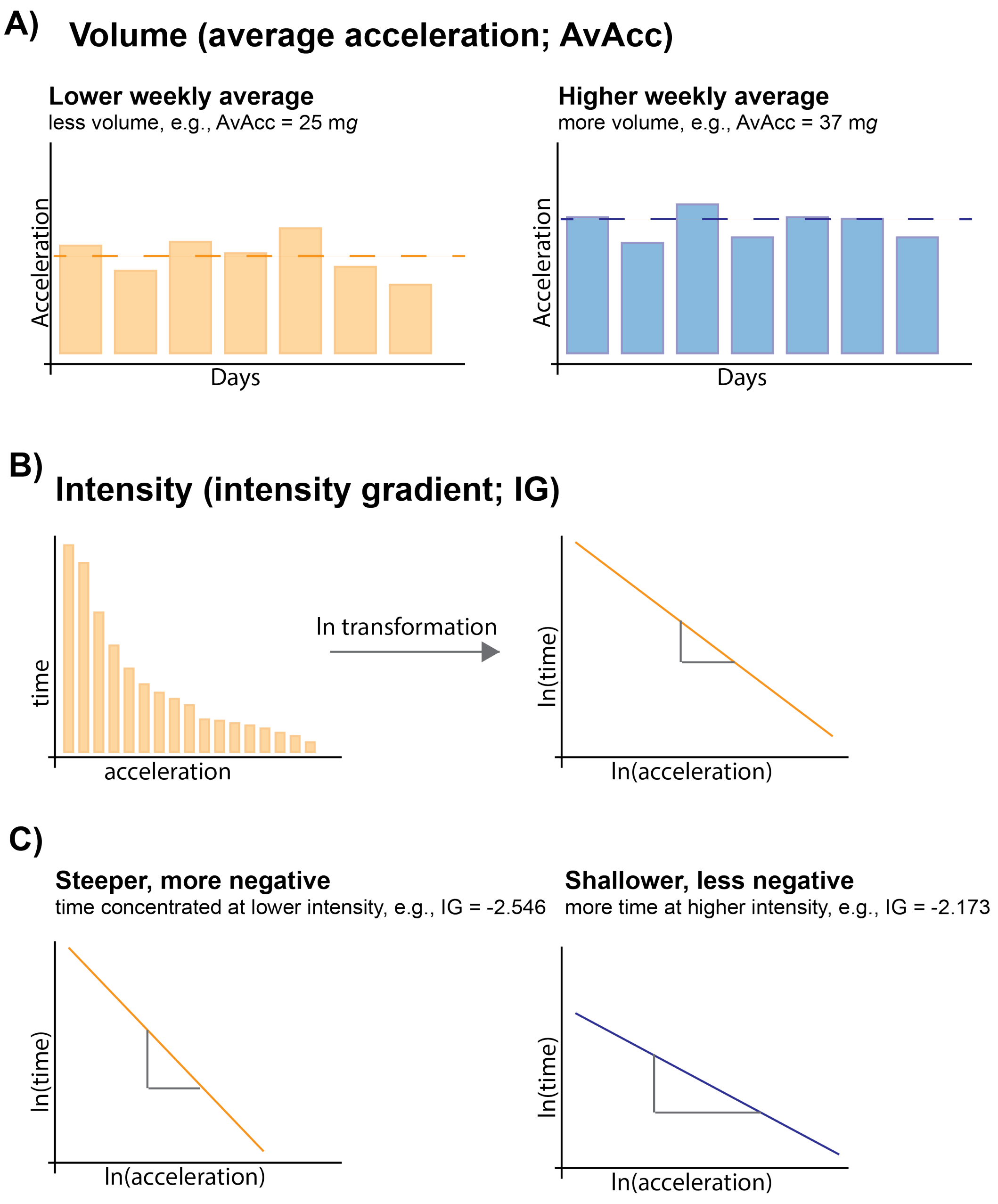

**FIG 3** Conceptual figure of the cut-point-free metrics: intensity gradient and average acceleration. A) Comparison of an inactive and active individual using the average acceleration. B) Calculation of the intensity gradient by fitting a linear regression to the time-intensity relationship on a natural log-log scale. C) Comparison of an inactive and active individual using the intensity gradient.

#### 5 Restricted cubic spline placement

We used restricted cubic splines with three knots to model the continuous variables shown in Table 3. Knot positions were selected using the `rcspline.eval()` function from the `Hmisc` R package v5.3.0.[12]

**TABLE 3** Restricted cubic spline knot placements for all survival models.

| Variable | Knot values |
|---|---|
| Intensity gradient | -3.021, -2.665, -2.383 |
| Average acceleration | 15.11, 25.92, 40.38 |
| Duration | 155.8, 295.4, 453 |
| Age | 48, 70, 79 |
| Waist-to-height ratio | 0.4754, 0.5869, 0.7283 |
| Body mass index | 21.5, 27.9, 38.1 |
| Waist circumference | 79.5, 98, 120.7 |

### 6 Population characteristics

Table 4 shows the imputed, survey-weighted cohort characteristics.

**TABLE 4** Survey-weighted cohort characteristics (overall and by sex) with imputed covariates

| Characteristic | **Overall** N = 179,836,080[1] | **Men** N = 85,310,147[1] | **Women** N = 94,525,932[1] |
|---|---|---|---|
| **Age, years** | 48 (35, 62) | 47 (34, 60) | 49 (36, 62) |
| **Body mass index, kg/m²** | 28.0 (24.3, 32.5) | 28.0 (24.9, 31.8) | 28.0 (23.8, 33.4) |
| **Waist-to-height ratio** | 0.58 (0.52, 0.65) | 0.58 (0.52, 0.63) | 0.59 (0.53, 0.67) |
| **Waist circumference, cm** | 98 (88, 109) | 101 (92, 111) | 96 (85, 107) |
| **Average acceleration, mg** | 26.23 (20.50, 32.87) | 27.31 (20.97, 34.79) | 25.54 (20.08, 31.20) |
| **Intensity gradient** | -2.648 (-2.805, -2.513) | -2.614 (-2.757, -2.476) | -2.681 (-2.837, -2.547) |
| **Duration, min/day** | 296.5 (226.3, 371.5) | 301.2 (228.1, 385.5) | 293.1 (225.6, 359.7) |
| **Follow-up, months** | 82.0 (69.0, 95.0) | 82.0 (69.0, 96.0) | 82.0 (70.0, 95.0) |
| **Poverty–income ratio** | 2.78 (1.31, 4.96) | 2.96 (1.39, 5.00) | 2.64 (1.26, 4.77) |
| **Education** | | | |
| less than high school | 28,337,063 (16%) | 14,143,342 (17%) | 14,193,721 (15%) |
| high school | 38,170,577 (21%) | 19,310,365 (23%) | 18,860,212 (20%) |
| more than high school | 113,328,440 (63%) | 51,856,441 (61%) | 61,472,000 (65%) |
| **Ethnicity** | | | |
| Mexican American | 14,976,190 (8.3%) | 7,778,466 (9.1%) | 7,197,724 (7.6%) |
| Other Hispanic | 10,689,026 (5.9%) | 5,096,952 (6.0%) | 5,592,073 (5.9%) |
| Non-Hispanic White | 120,462,848 (67%) | 57,001,757 (67%) | 63,461,091 (67%) |
| Non-Hispanic Black | 20,448,175 (11%) | 9,015,752 (11%) | 11,432,422 (12%) |
| Other/Multi-Racial | 13,259,842 (7.4%) | 6,417,219 (7.5%) | 6,842,622 (7.2%) |
| **Smoking status** | | | |
| Never | 100,863,586 (56%) | 42,583,899 (50%) | 58,279,687 (62%) |
| Former | 44,245,288 (25%) | 24,202,779 (28%) | 20,042,509 (21%) |
| Current | 34,727,206 (19%) | 18,523,469 (22%) | 16,203,737 (17%) |
| **Cancer, yes** | 19,826,372 (11%) | 8,393,256 (9.8%) | 11,433,116 (12%) |
| **Cardiovascular disease, yes** | 16,929,099 (9.4%) | 8,638,577 (10%) | 8,290,523 (8.8%) |
| **Diabetes, yes** | 23,289,346 (13%) | 11,422,345 (13%) | 11,867,000 (13%) |
| **Sys. blood pressure, mmHg** | 119 (111, 131) | 121 (113, 131) | 117 (108, 130) |
| **Dia. blood pressure, mmHg** | 71 (64, 78) | 72 (65, 79) | 70 (63, 77) |

[1] Median (Q1, Q3); n (%)

Duration of physical activity is defined as the time spent above 40 milli-gravitational units in a day. Abbreviations: Sys, systolic; Dia, diastolic; mg, milli-gravitational units.

### 7 Main analyses

Since our cohort included both adults with and without a history of CVD, we visualised the distribution of exposure variables (i.e., intensity, volume, and duration of PA) stratified by history of CVD, defined as binary variable. Figure 4 shows that there is sufficient overlap in the distribution of exposures between strata, suggesting that there is support for the models to reflect both adults with and without chronic non-communicable diseases in the cohort.

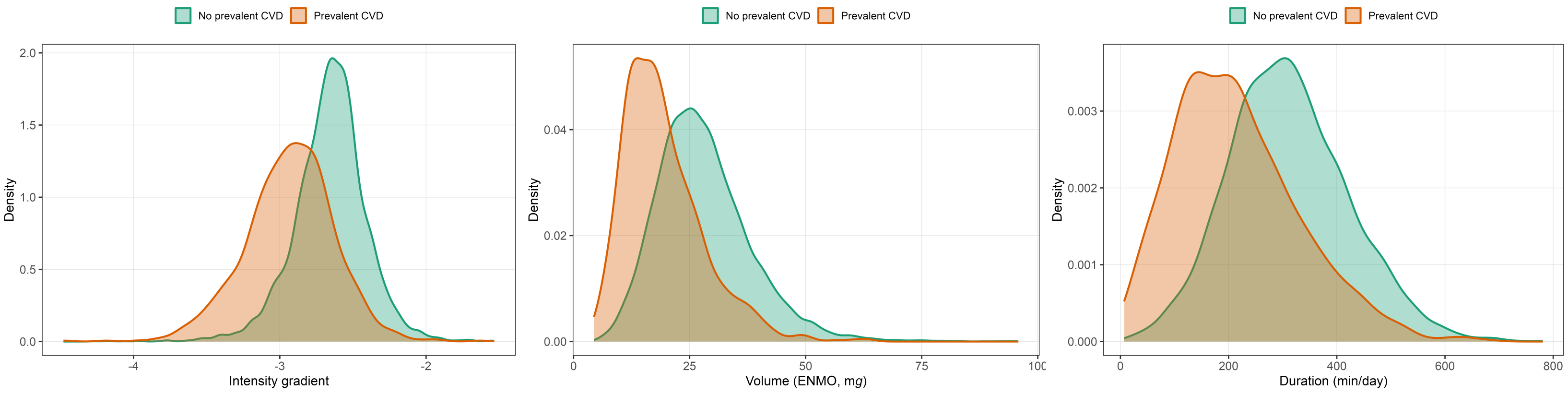

**(a)** Intensity gradient **(b)** Average acceleration **(c)** Duration

**FIG 4** Distribution of physical activity intensity (top), volume (middle), and duration (bottom) by history of cardiovascular disease (CVD). Density curves are overlaid to visualise the distribution shape and were created based on one imputed dataset.

Figure 5 shows the distribution of exposure variables across age, sex, and waist-to-height ratio.

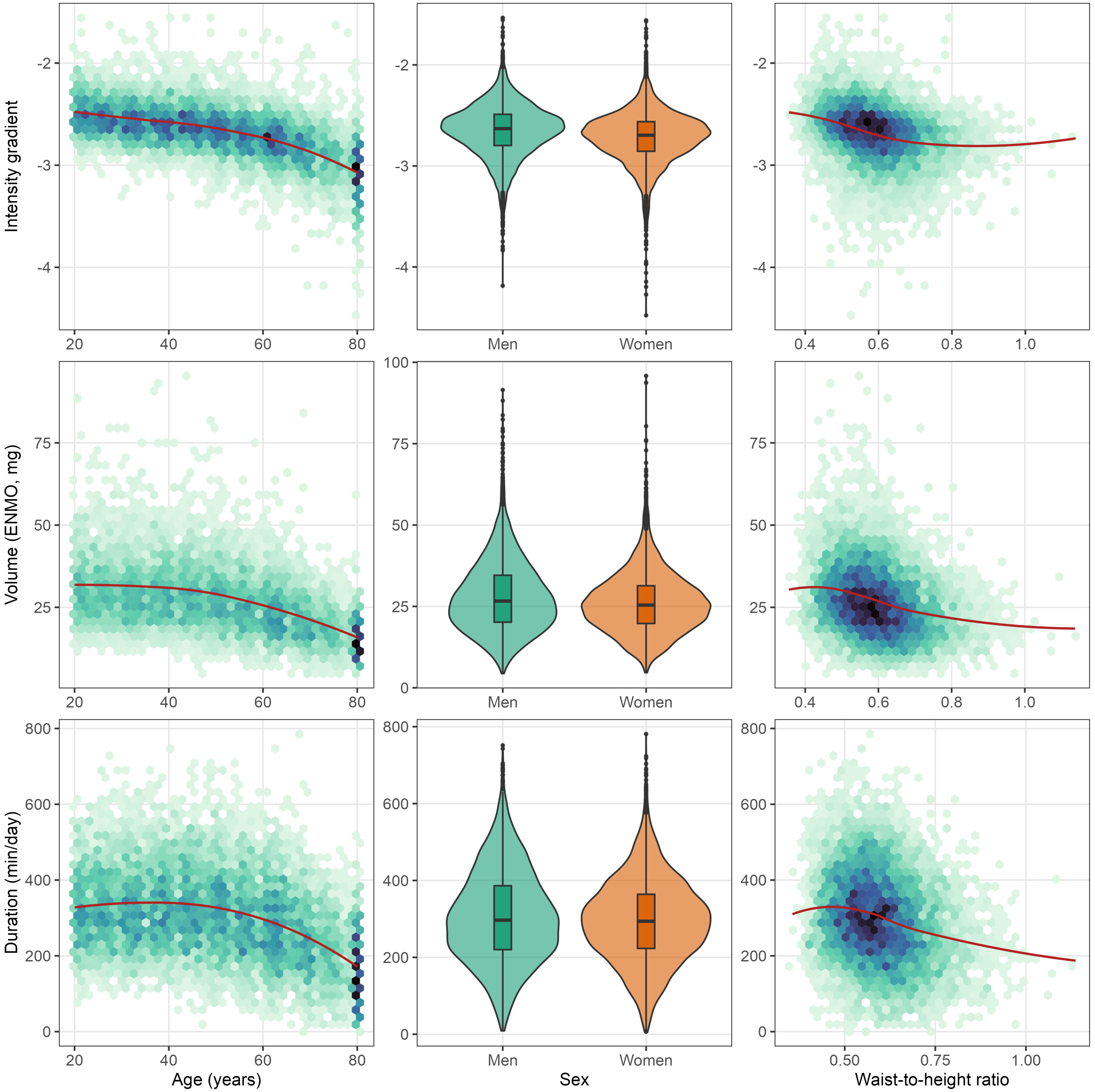

**FIG 5** Distribution of physical activity intensity (top), volume (middle), and duration (bottom) by age (left), sex (middle), and waist-to-height ratio (right). For age and waist-to-height ratio, hexagonal bins represent the density of observations (darker shading indicates a higher concentration of participants), with a locally estimated scatterplot smoothing curve. For sex, violin plots show the full distribution of each outcome for men and women, with boxplots indicating the median and interquartile range.

##### 7.1 Comparison of cause-specific and subdistribution hazard interaction tests

**TABLE 5** Comparison of joint interaction tests under cause-specific Cox (CSH) and Fine-Gray subdistribution hazard (SHR) models.

| Model | | Model size | | | Interaction test p-value | | |
|---|---|---|---|---|---|---|---|
| PA metric | Association modifier | CVD events | Parameters | Events/param. | Cause-specific | Fine-Gray (SHR) | Δ (FG - CSH)[1] |
| Intensity | Age | 268 | 23 | 11.7 | 0.006 | 0.005 | -0.001 |
| Volume | Age | 268 | 23 | 11.7 | 0.526 | 0.389 | -0.137 |
| Duration | Age | 268 | 23 | 11.7 | 0.633 | 0.404 | -0.229 |
| Intensity | Sex | 268 | 22 | 12.2 | 0.855 | 0.931 | 0.075 |
| Volume | Sex | 268 | 22 | 12.2 | 0.537 | 0.794 | 0.257 |
| Duration | Sex | 268 | 22 | 12.2 | 0.811 | 0.948 | 0.137 |
| Intensity | Waist-to-height ratio | 268 | 23 | 11.7 | 0.021 | 0.093 | 0.071 |
| Volume | Waist-to-height ratio | 268 | 23 | 11.7 | 0.006 | 0.039 | 0.033 |
| Duration | Waist-to-height ratio | 268 | 23 | 11.7 | 0.005 | 0.035 | 0.031 |
| Intensity | Body mass index | 268 | 23 | 11.7 | 0.115 | 0.713 | 0.599 |
| Volume | Body mass index | 268 | 23 | 11.7 | 0.016 | 0.127 | 0.111 |
| Duration | Body mass index | 268 | 23 | 11.7 | 0.011 | 0.092 | 0.080 |
| Intensity | Waist circumference | 268 | 23 | 11.7 | 0.090 | 0.224 | 0.134 |
| Volume | Waist circumference | 268 | 23 | 11.7 | 0.018 | 0.111 | 0.093 |
| Duration | Waist circumference | 268 | 23 | 11.7 | 0.015 | 0.092 | 0.077 |

[1] Δ (FG - CSH) is the change in p-value when moving from the cause-specific to the Fine-Gray approach; positive values indicate weaker evidence for the interaction after accounting for non-CVD mortality as a competing risk.

The main text reports associations between physical activity and CVD mortality using Fine-Gray subdistribution hazard models, which account for non-CVD death as a competing risk. As recommended by Latouche et al. [13], we additionally fitted the corresponding cause-specific Cox models for all models reported in the main text, plus the waist circumference and body mass index models reported in the supplement, to characterise how inference about these interactions depends on the choice of competing-risk approach. The cause-specific model treats non-CVD deaths as censored observations and estimates CVD mortality risk among adults currently alive. The Fine-Gray model retains adults who died of non-CVD causes in an extended risk set with a time-dependent weight and estimates the association of covariates with the cumulative incidence of CVD mortality.[14] Table 5 summarises the joint design-based test of the interaction term under both approaches for each of the 15 models. p-values are from Rao-Scott likelihood ratio tests of the interaction term, run in each of the five imputed datasets and combined with the median-P rule.[15,16] Evidence for interactions with adiposity measures was stronger in the cause-specific models (e.g., waist-to-height ratio: p = 0.005–0.021) than in the Fine-Gray models (p = 0.035–0.093).

#### 7.2 Joint analyses of physical activity and age

In all panels below, subdistribution hazard ratios and absolute risks were pooled across the five imputed datasets. Absolute risks were predicted at 60 months (5 years) for a reference profile with each categorical covariate at its most common category and each continuous covariate at its median: women aged 50 years (unless age or sex was the association modifier) of non-Hispanic White ethnicity with more than high school education, who had never smoked, had no prevalent CVD, cancer, or diabetes, and had median income-to-poverty ratio (1.97), body mass index (27.9 kg/m$^2$; not included in the waist-to-height ratio and waist circumference models), and mean arterial pressure (87.3 mmHg). Points in the absolute-risk line plots mark the 25$^{th}$ and 50$^{th}$ percentile of each exposure in the analytic sample. Contour and absolute-risk surface plots span the survey-weighted 2.5$^{th}$ to 97.5$^{th}$ percentiles of each exposure in the full cohort; each line in the dose-response and absolute-risk line plots spans these percentiles within its stratum (participants grouped at the midpoints between stratum values). Ranges were extended to include the reference value where needed. Plot ranges do not affect the models, which used the full range of each exposure.

Figure 6, Figure 7, and Figure 8 show the continuous interaction between age and physical activity intensity, volume, and duration for CVD mortality, accounting for non-CVD mortality as a competing risk, alongside the corresponding predicted absolute risk.

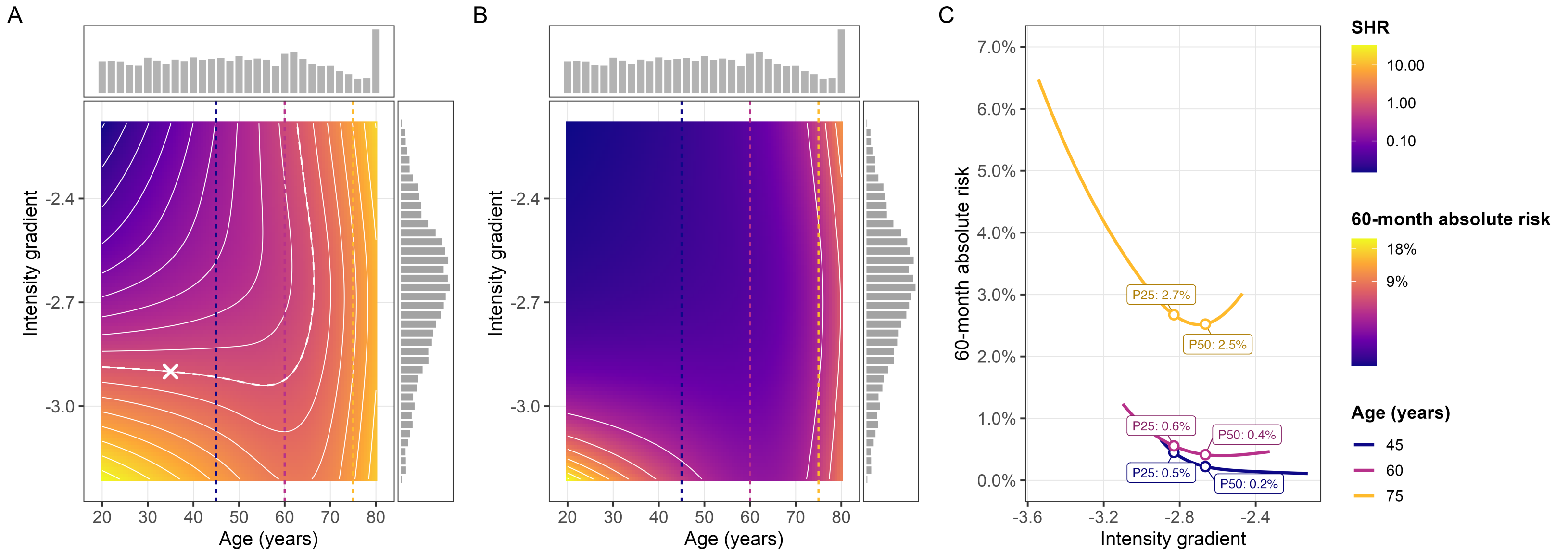

**FIG 6** Association between age and physical activity intensity and CVD mortality, from a survey-weighted Fine-Gray subdistribution hazard model accounting for non-CVD mortality as a competing risk. (A) Subdistribution hazard ratio (SHR) contour plot (isohazard curves; X marks the reference point, SHR = 1). (B) Predicted 60-month absolute CVD mortality risk over the same age/exposure space. (C) Predicted 60-month absolute risk by exposure at ages 45, 60, and 75. Dashed vertical lines in panels A and B mark these same three ages.

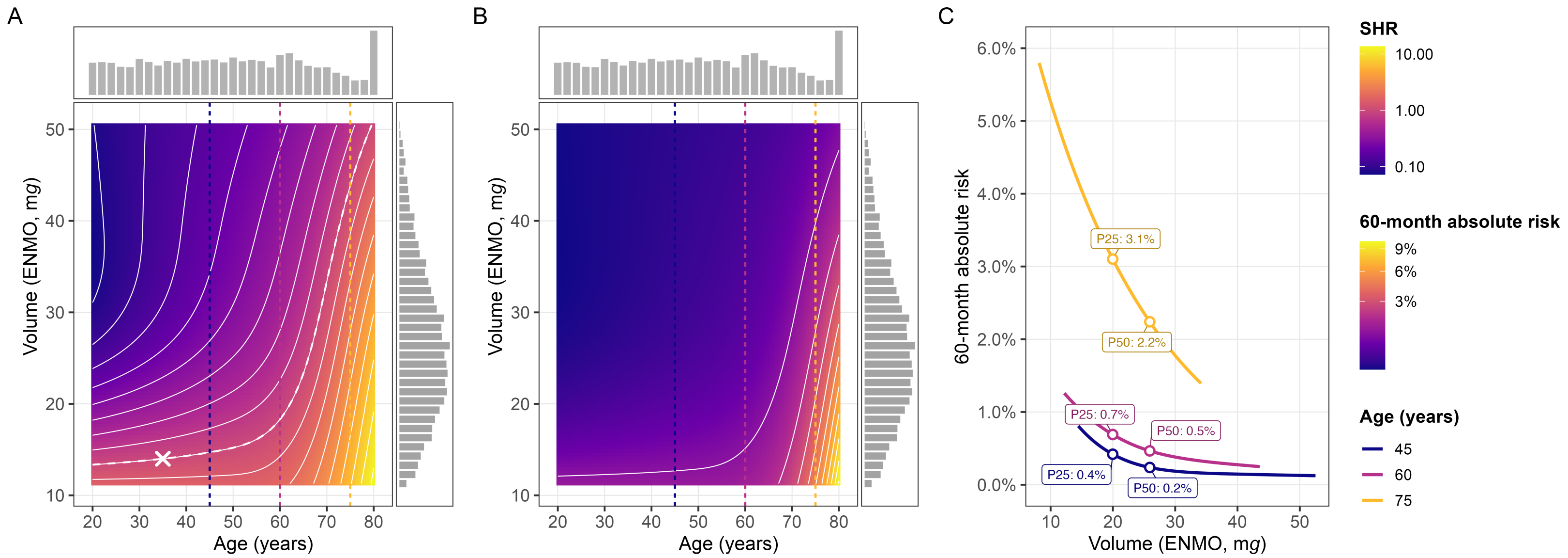

**FIG 7** Association between age and physical activity volume and CVD mortality, from a survey-weighted Fine-Gray subdistribution hazard model accounting for non-CVD mortality as a competing risk. (A) Subdistribution hazard ratio (SHR) contour plot (isohazard curves; X marks the reference point, SHR = 1). (B) Predicted 60-month absolute CVD mortality risk over the same age/exposure space. (C) Predicted 60-month absolute risk by exposure at ages 45, 60, and 75. Dashed vertical lines in panels A and B mark these same three ages.

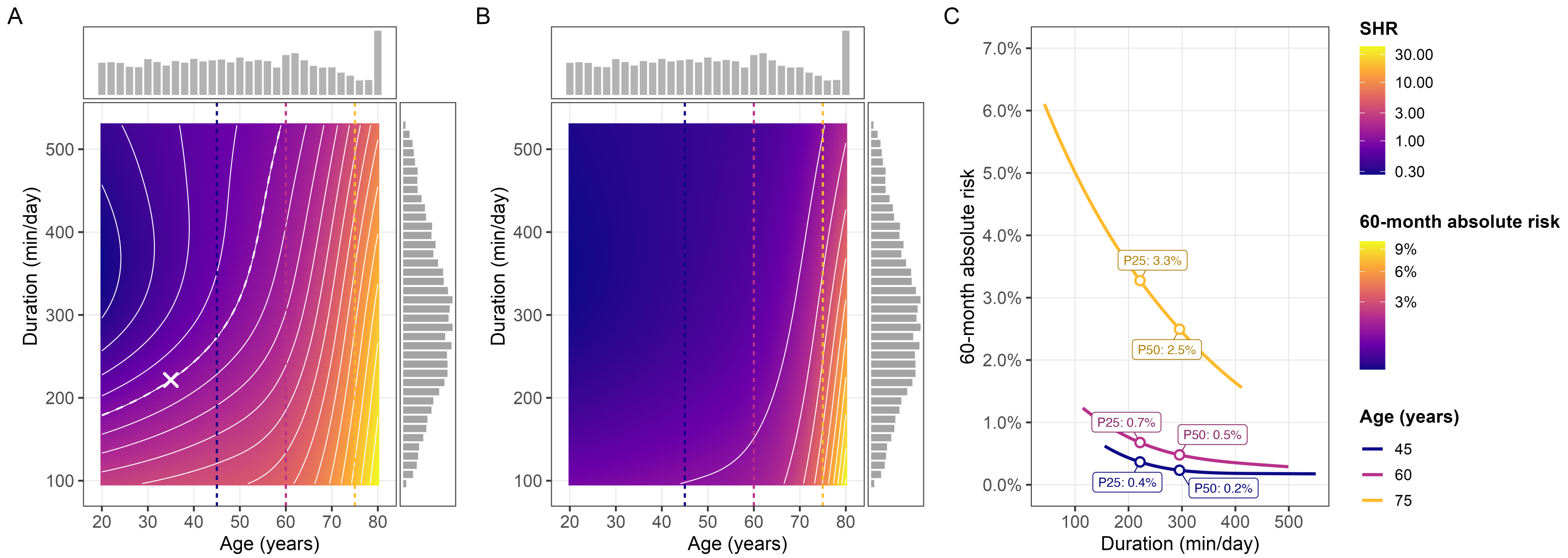

**FIG 8** Association between age and physical activity duration and CVD mortality, from a survey-weighted Fine-Gray subdistribution hazard model accounting for non-CVD mortality as a competing risk. (A) Subdistribution hazard ratio (SHR) contour plot (isohazard curves; X marks the reference point, SHR = 1). (B) Predicted 60-month absolute CVD mortality risk over the same age/exposure space. (C) Predicted 60-month absolute risk by exposure at ages 45, 60, and 75. Dashed vertical lines in panels A and B mark these same three ages.

#### 7.3 Joint analyses of physical activity and sex

Figure 9 shows the predicted absolute risk estimates of CVD mortality based on the interaction between sex and physical activity intensity, volume, and duration.

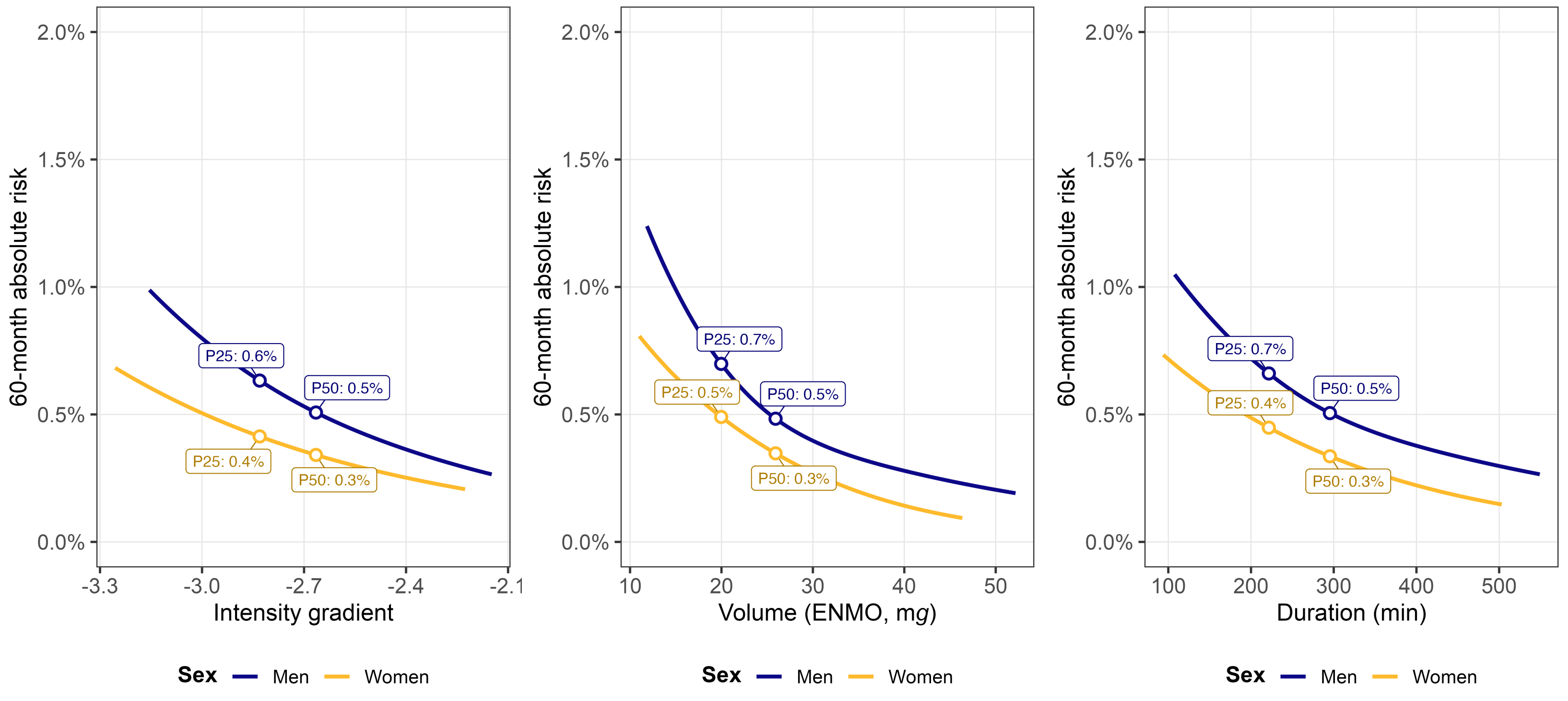

**(a)** Intensity gradient **(b)** Volume **(c)** Duration

**FIG 9** Predicted 60-month absolute CVD mortality risk by sex across intensity, volume, and duration of physical activity, from survey-weighted Fine-Gray subdistribution hazard models accounting for non-CVD mortality as a competing risk. Points and labels mark the predicted absolute risk at the 25$^{th}$ and 50$^{th}$ percentile of each exposure in the analytic sample, separately for men and women.

#### 7.4 Joint analyses of physical activity and body morphology

We investigated the joint association of physical activity and body morphology with CVD mortality using a range of body morphology measures including waist-to-height ratio, waist circumference, and body mass index. Waist-to-height ratio captures central adiposity while accounting for body morphology. Moreover, a recent meta-analysis has demonstrated superiority over waist circumference and body mass index for detecting cardiometabolic risk factors in both sexes, supporting its use as a screening tool.[17]

##### 7.4.1 Waist-to-height ratio

Figure 10, Figure 11, and Figure 12 show the continuous interaction between waist-to-height ratio and physical activity intensity, volume, and duration for CVD mortality, accounting for non-CVD mortality as a competing risk, alongside the corresponding predicted absolute risk.

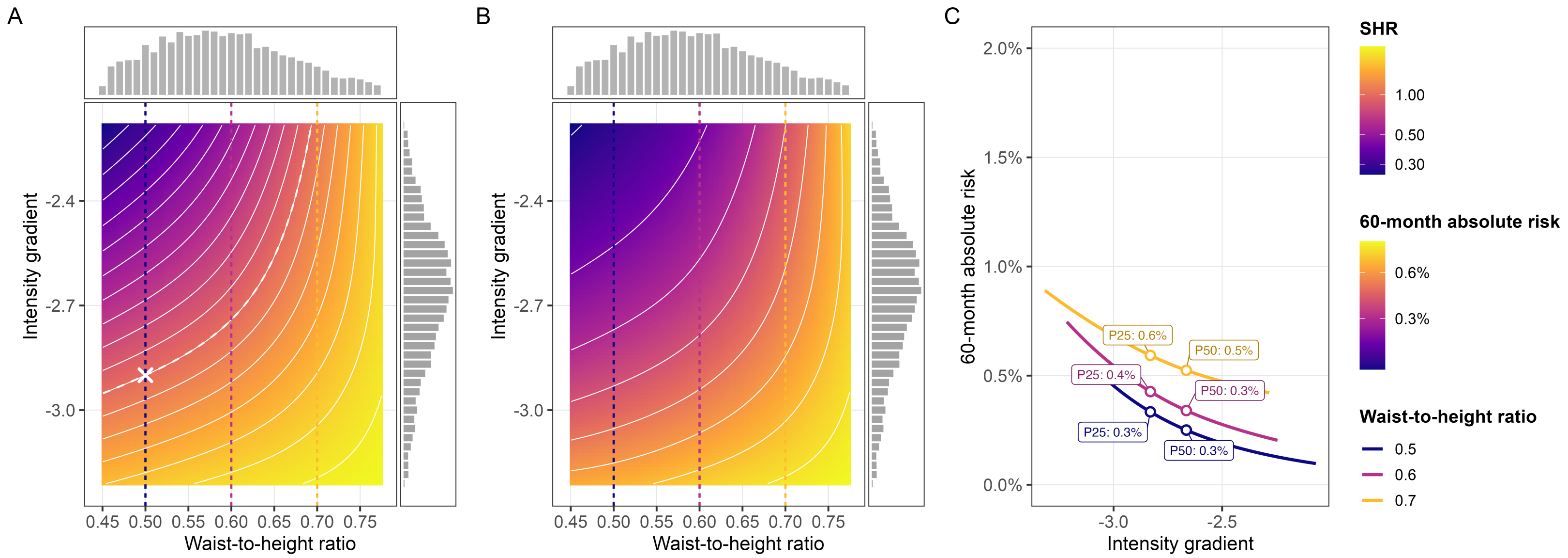

**FIG 10** Association between waist-to-height ratio and physical activity intensity and CVD mortality, from a survey-weighted Fine-Gray subdistribution hazard model accounting for non-CVD mortality as a competing risk. (A) Subdistribution hazard ratio (SHR) contour plot (isohazard curves; X marks the reference point, SHR = 1). (B) Predicted 60-month absolute CVD mortality risk over the same waist-to-height ratio/exposure space. (C) Predicted 60-month absolute risk by exposure at waist-to-height ratios of 0.5, 0.6, and 0.7. Dashed vertical lines in panels A and B mark these same three values.

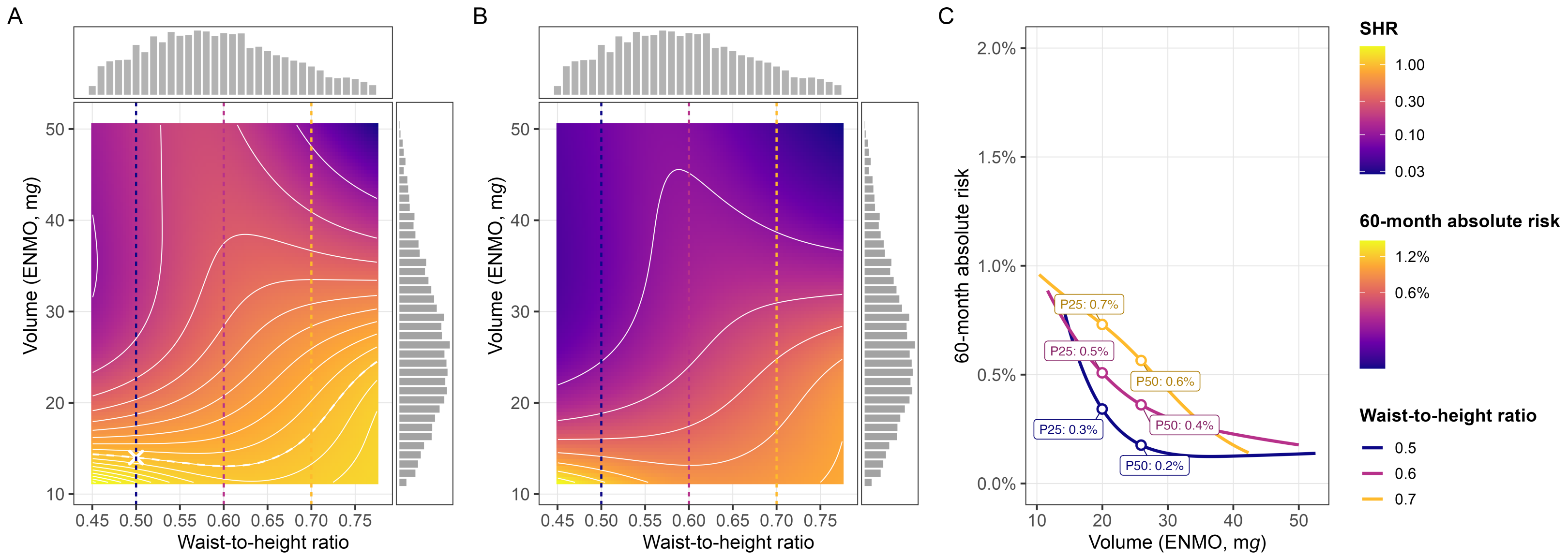

**FIG 11** Association between waist-to-height ratio and physical activity volume and CVD mortality, from a survey-weighted Fine-Gray subdistribution hazard model accounting for non-CVD mortality as a competing risk. (A) Subdistribution hazard ratio (SHR) contour plot (isohazard curves; X marks the reference point, SHR = 1). (B) Predicted 60-month absolute CVD mortality risk over the same waist-to-height ratio/exposure space. (C) Predicted 60-month absolute risk by exposure at waist-to-height ratios of 0.5, 0.6, and 0.7. Dashed vertical lines in panels A and B mark these same three values.

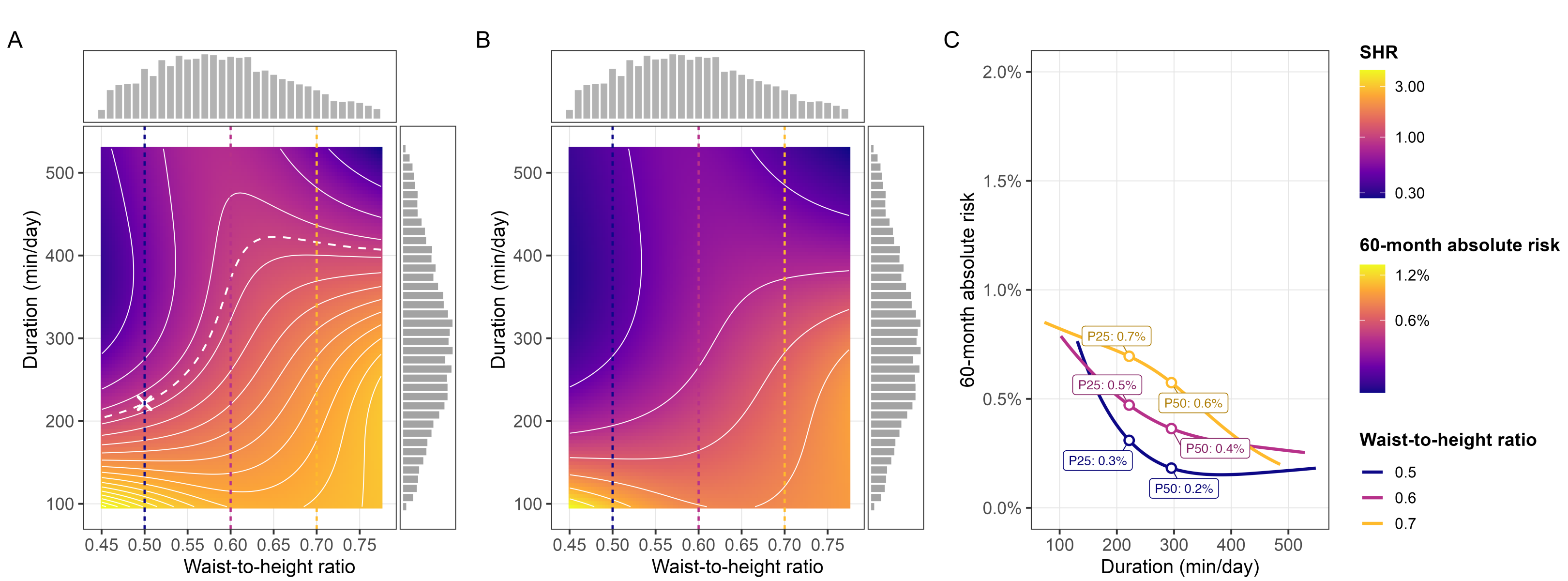

**FIG 12** Association between waist-to-height ratio and physical activity duration and CVD mortality, from a survey-weighted Fine-Gray subdistribution hazard model accounting for non-CVD mortality as a competing risk. (A) Subdistribution hazard ratio (SHR) contour plot (isohazard curves; X marks the reference point, SHR = 1). (B) Predicted 60-month absolute CVD mortality risk over the same waist-to-height ratio/exposure space. (C) Predicted 60-month absolute risk by exposure at waist-to-height ratios of 0.5, 0.6, and 0.7. Dashed vertical lines in panels A and B mark these same three values.

##### 7.4.2 Waist circumference

The following analyses show the joint associations of physical activity and waist circumference with CVD mortality (see Figure 13). Waist circumferences of 88, 98, and 109 cm correspond to the 25$^{th}$, 50$^{th}$, and 75$^{th}$ percentiles of the cohort distribution. The absolute-risk panels below use 80, 100, and 120 cm.

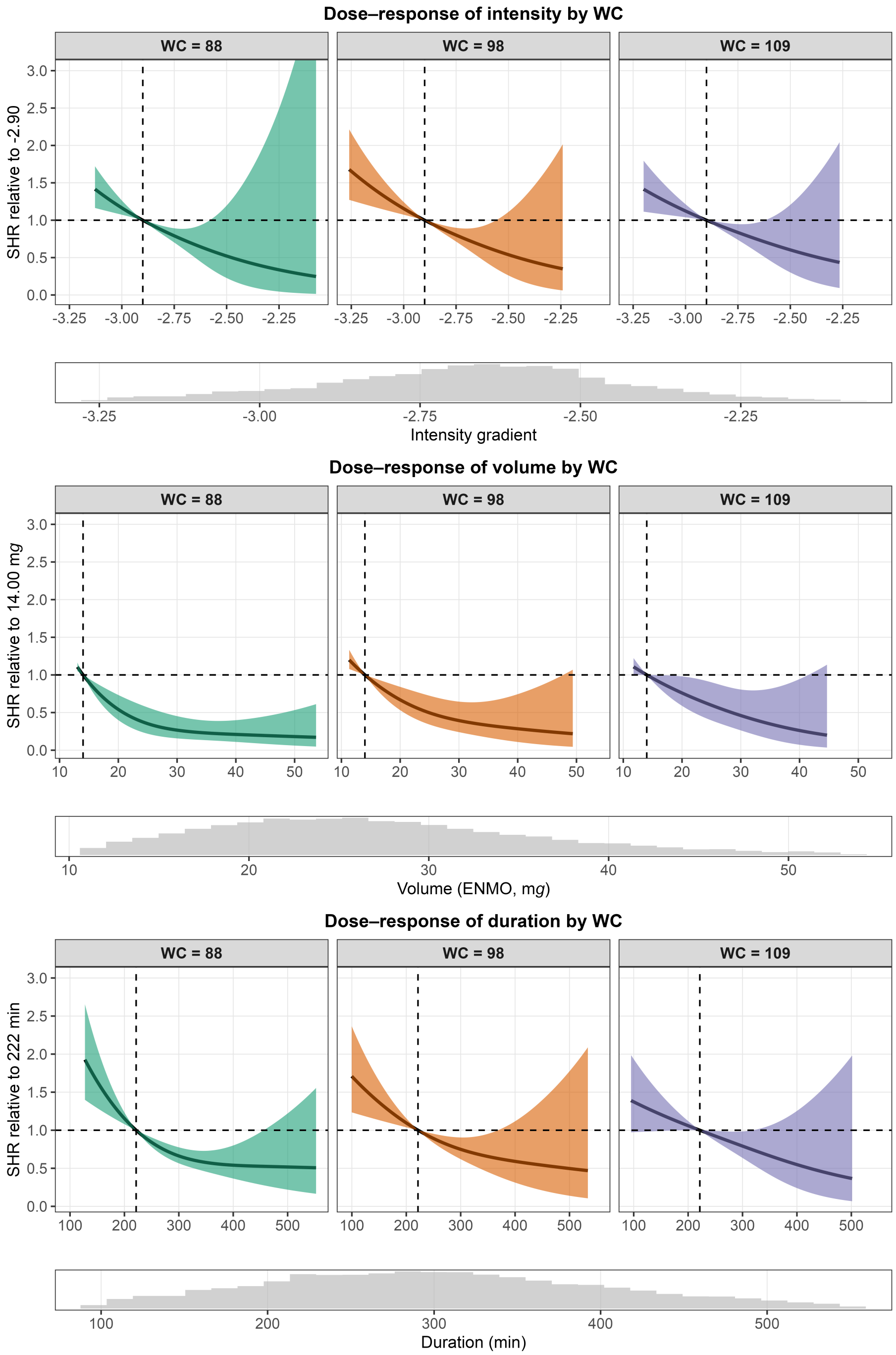

**FIG 13** Dose-response relationship between physical activity intensity (top), volume (middle), and duration (bottom) and cardiovascular disease mortality subdistribution hazard ratio (SHR), predicted for waist circumference (WC) 88, 98, and 109 cm, from survey-weighted Fine-Gray models accounting for non-CVD mortality as a competing risk. Models included an interaction term between each physical activity metric and WC. Histograms depict the distribution of the corresponding exposure variable.

#### 7.5 Joint analyses of physical activity and waist circumference

Figure 14, Figure 15, and Figure 16 show the continuous interaction between waist circumference and physical activity intensity, volume, and duration for CVD mortality, accounting for non-CVD mortality as a competing risk, alongside the corresponding predicted absolute risk.

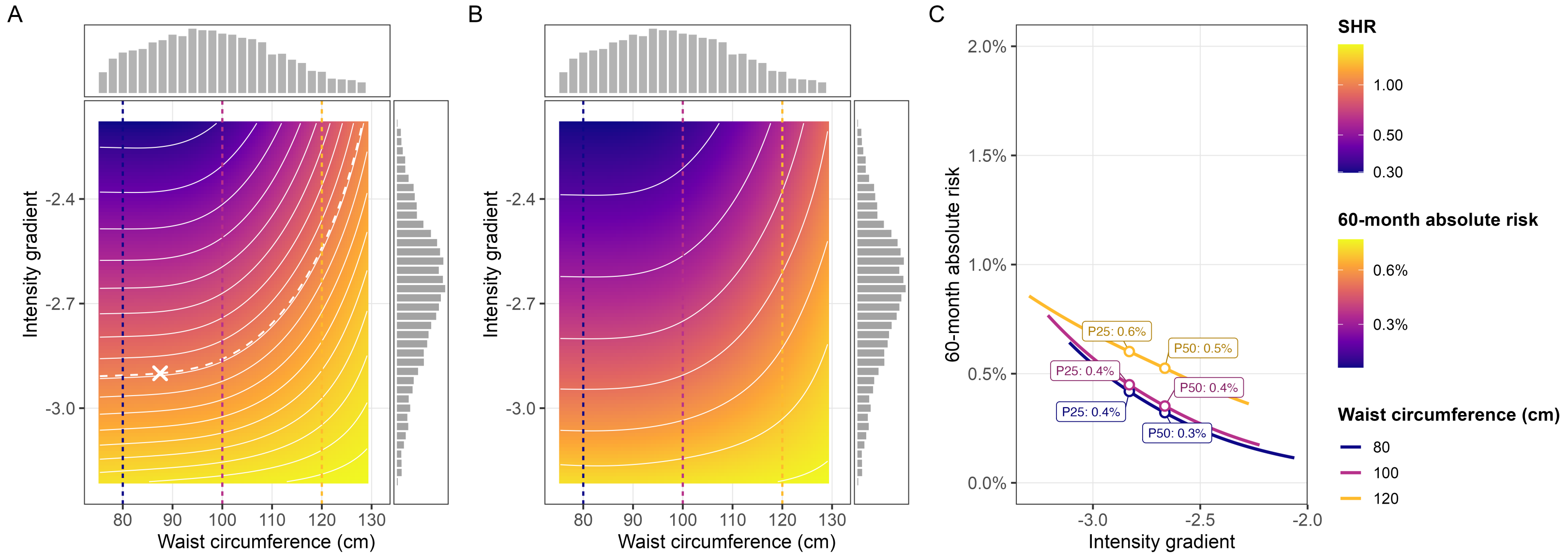

**FIG 14** Association between waist circumference and physical activity intensity and CVD mortality, from a survey-weighted Fine-Gray subdistribution hazard model accounting for non-CVD mortality as a competing risk. (A) Subdistribution hazard ratio (SHR) contour plot (isohazard curves; X marks the reference point, SHR = 1). (B) Predicted 60-month absolute CVD mortality risk over the same waist circumference/exposure space. (C) Predicted 60-month absolute risk by exposure at waist circumferences of 80, 100, and 120 cm. Dashed vertical lines in panels A and B mark these same three values.

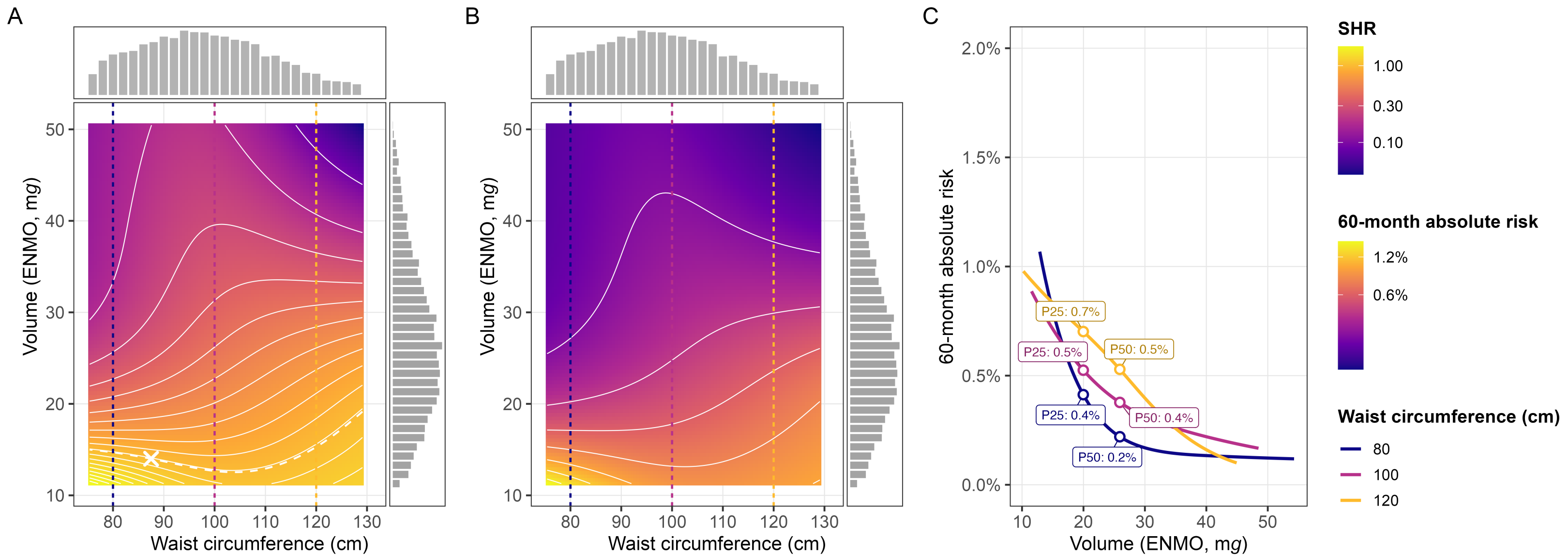

**FIG 15** Association between waist circumference and physical activity volume and CVD mortality, from a survey-weighted Fine-Gray subdistribution hazard model accounting for non-CVD mortality as a competing risk. (A) Subdistribution hazard ratio (SHR) contour plot (isohazard curves; X marks the reference point, SHR = 1). (B) Predicted 60-month absolute CVD mortality risk over the same waist circumference/exposure space. (C) Predicted 60-month absolute risk by exposure at waist circumferences of 80, 100, and 120 cm. Dashed vertical lines in panels A and B mark these same three values.

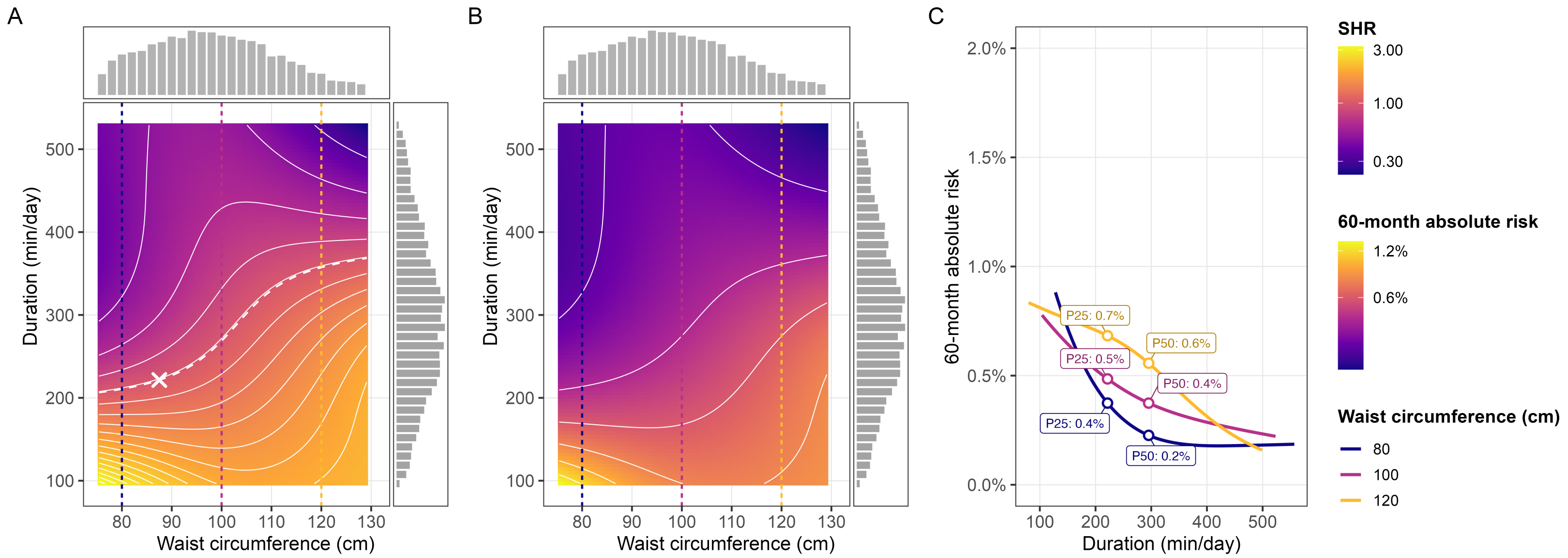

**FIG 16** Association between waist circumference and physical activity duration and CVD mortality, from a survey-weighted Fine-Gray subdistribution hazard model accounting for non-CVD mortality as a competing risk. (A) Subdistribution hazard ratio (SHR) contour plot (isohazard curves; X marks the reference point, SHR = 1). (B) Predicted 60-month absolute CVD mortality risk over the same waist circumference/exposure space. (C) Predicted 60-month absolute risk by exposure at waist circumferences of 80, 100, and 120 cm. Dashed vertical lines in panels A and B mark these same three values.

###### 7.5.1 Body mass index

The following analyses show the joint associations of physical activity and body mass index with CVD mortality (see Figure 17). The body mass index values of 20, 30, and 40 kg/$m^2$ correspond to healthy weight, obesity class 1, and obesity class 3, respectively.

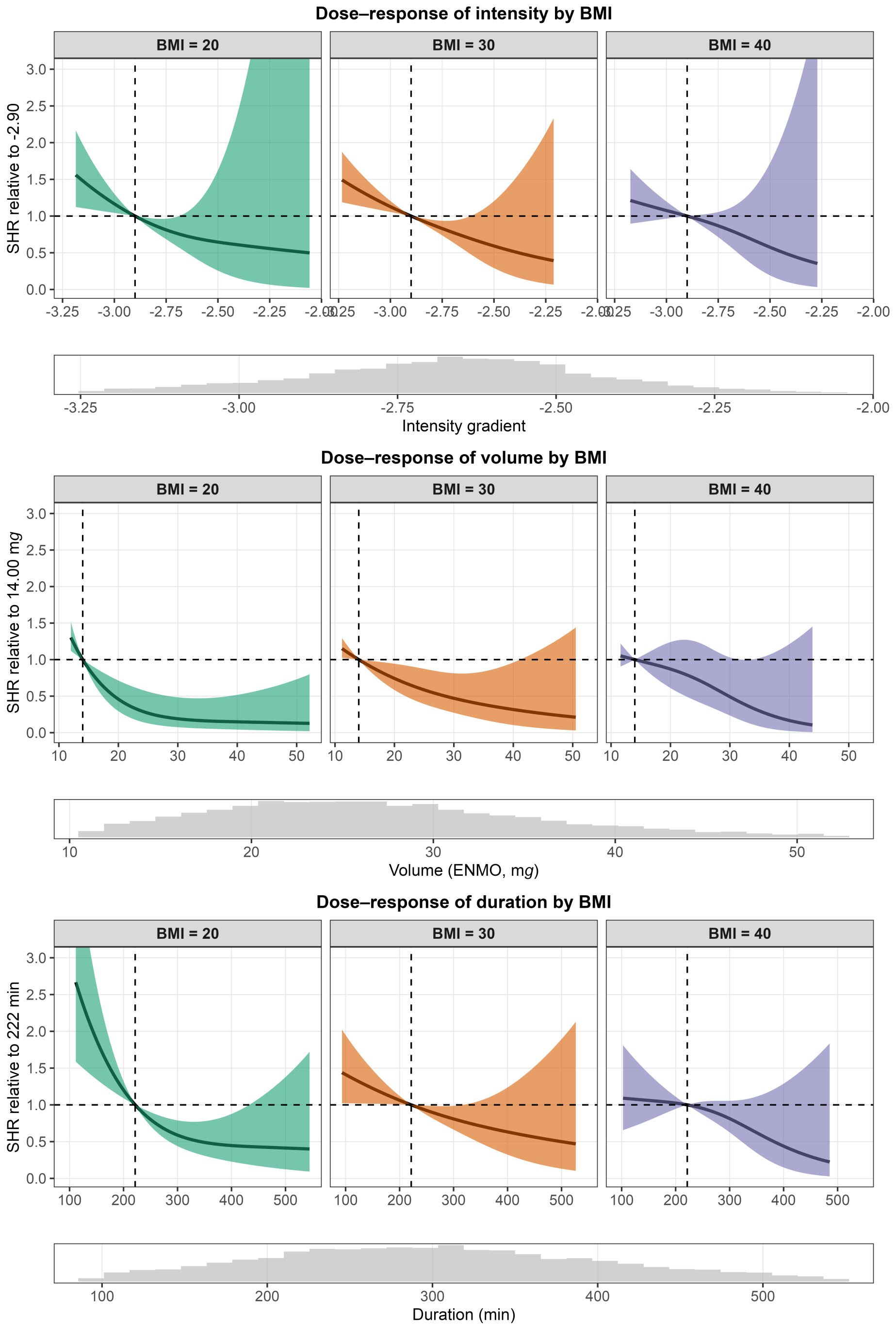

**FIG 17** Dose-response relationship between physical activity intensity (top), volume (middle), and duration (bottom) and cardiovascular disease mortality subdistribution hazard ratio (SHR), predicted for body mass index (BMI) 20, 30, and 40 kg/m$^2$, from survey-weighted Fine-Gray models accounting for non-CVD mortality as a competing risk. Models included an interaction term between each physical activity metric and BMI. Histograms depict the distribution of the corresponding exposure variable.

#### 7.6 Joint analyses of physical activity and body mass index

Figure 18, Figure 19, and Figure 20 show the continuous interaction between BMI and physical activity intensity, volume, and duration for CVD mortality, accounting for non-CVD mortality as a competing risk, alongside the corresponding predicted absolute risk.

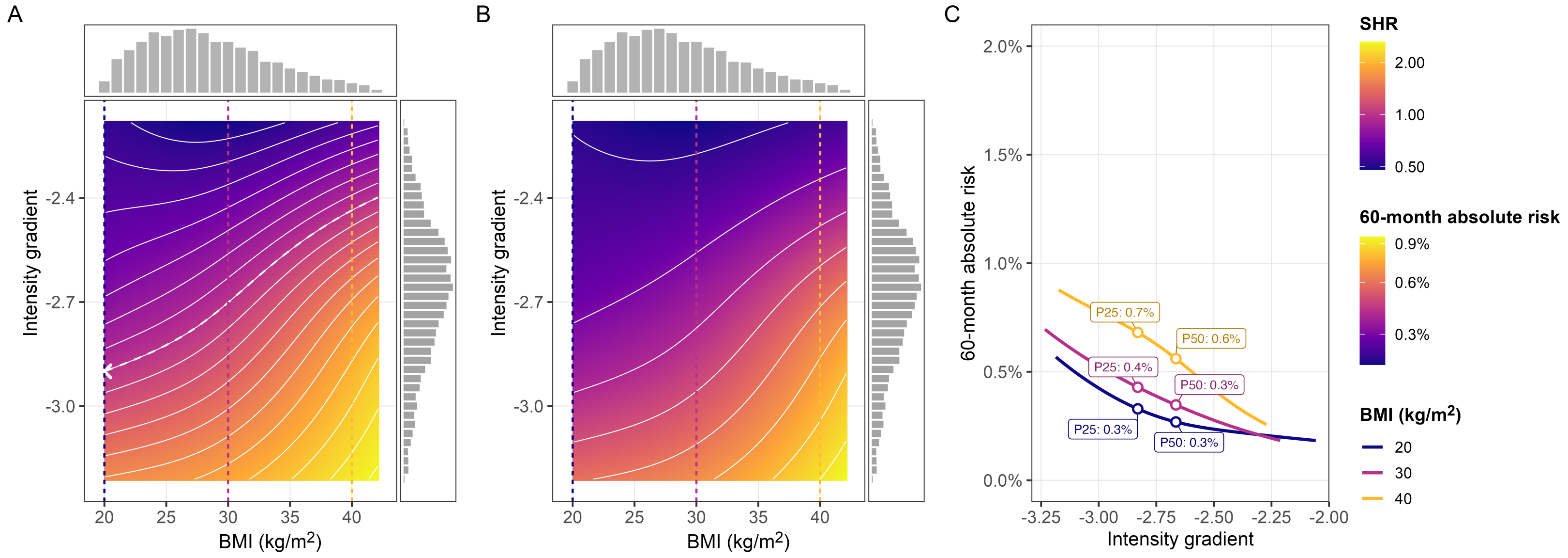

**FIG 18** Association between BMI and physical activity intensity and CVD mortality, from a survey-weighted Fine-Gray subdistribution hazard model accounting for non-CVD mortality as a competing risk. (A) Subdistribution hazard ratio (SHR) contour plot (isohazard curves; X marks the reference point, SHR = 1). (B) Predicted 60-month absolute CVD mortality risk over the same BMI/exposure space. (C) Predicted 60-month absolute risk by exposure at BMI 20, 30, and 40 kg/m². Dashed vertical lines in panels A and B mark these same three values.

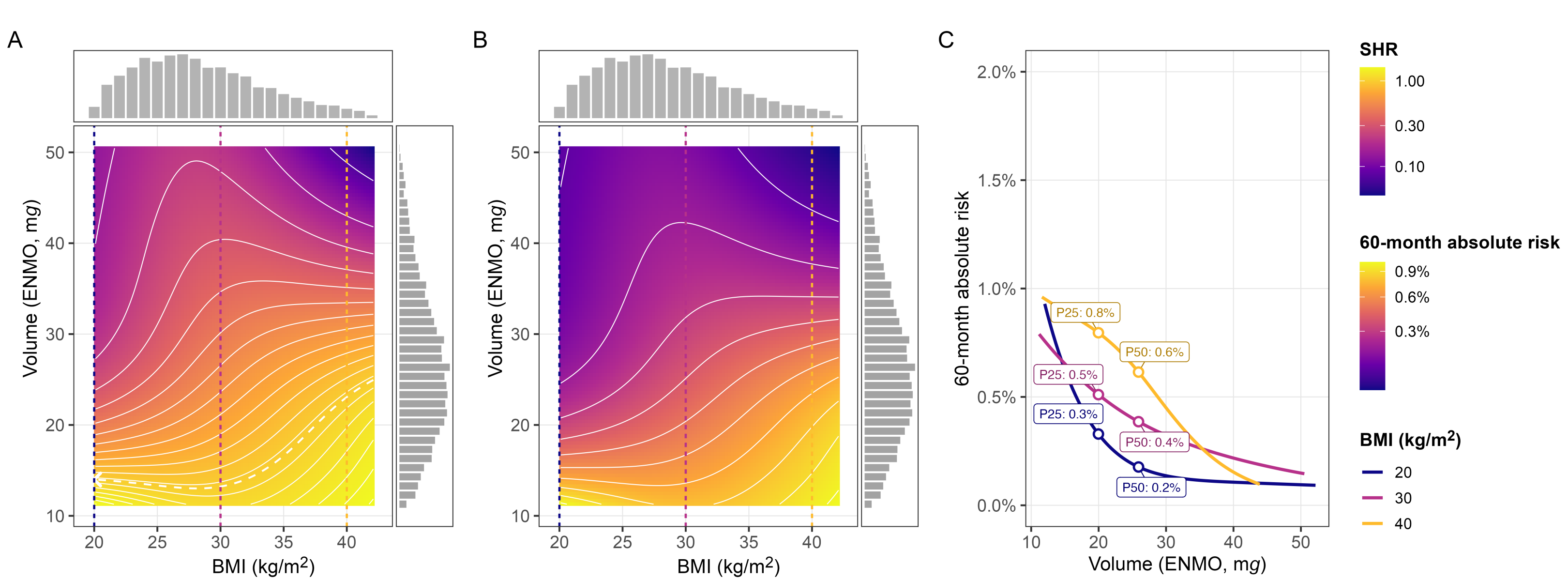

**FIG 19** Association between BMI and physical activity volume and CVD mortality, from a survey-weighted Fine-Gray subdistribution hazard model accounting for non-CVD mortality as a competing risk. (A) Subdistribution hazard ratio (SHR) contour plot (isohazard curves; X marks the reference point, SHR = 1). (B) Predicted 60-month absolute CVD mortality risk over the same BMI/exposure space. (C) Predicted 60-month absolute risk by exposure at BMI 20, 30, and 40 kg/m². Dashed vertical lines in panels A and B mark these same three values.

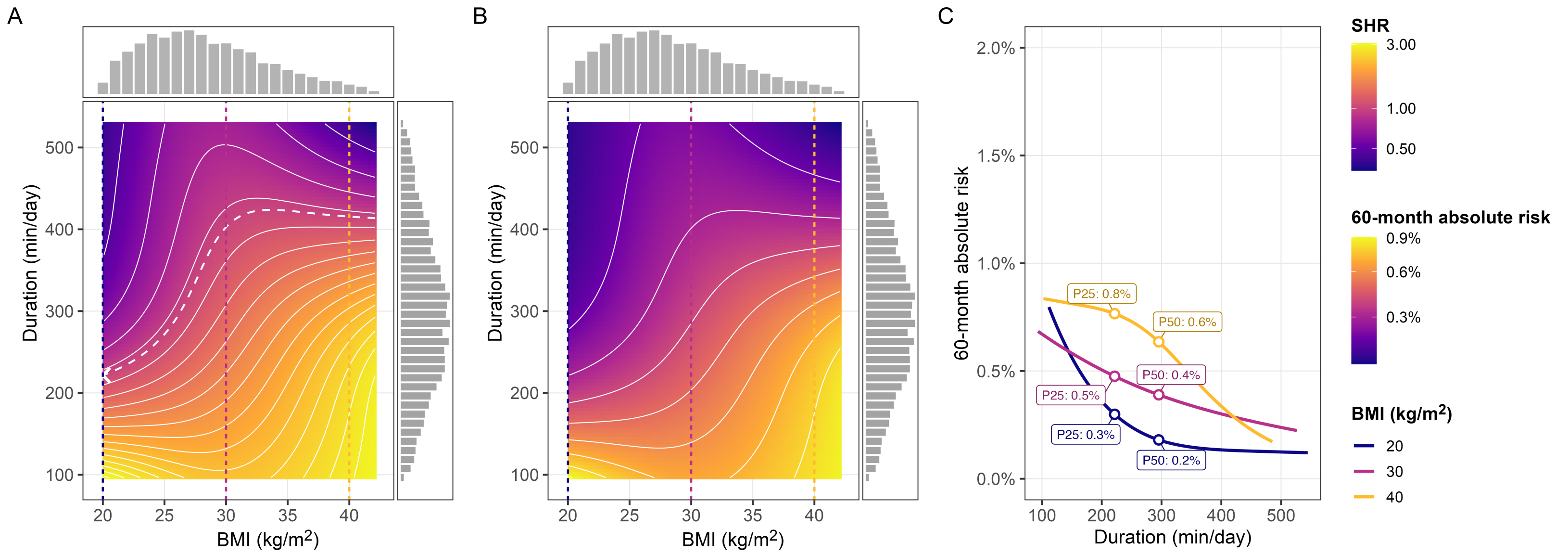

**FIG 20** Association between BMI and physical activity duration and CVD mortality, from a survey-weighted Fine-Gray subdistribution hazard model accounting for non-CVD mortality as a competing risk. (A) Subdistribution hazard ratio (SHR) contour plot (isohazard curves; X marks the reference point, SHR = 1). (B) Predicted 60-month absolute CVD mortality risk over the same BMI/exposure space. (C) Predicted 60-month absolute risk by exposure at BMI 20, 30, and 40 kg/m². Dashed vertical lines in panels A and B mark these same three values.

#### 7.7 Translating findings into public health messages

Table 6 adds further information to Figure 4 presented in the manuscript. The resulting intensity and volume changes are taken from [18]. These findings are derived from modelled changes using observational data with physical activity measured at a single timepoint and should thus be interpreted as illustrative examples that are not based on actual within-person changes in physical activity.

**TABLE 6** Estimated subdistribution hazard ratios for inactive and median activity profiles across age, sex, and adiposity interactions.

| | | SHR (95% CI) per scenario | | |
|---|---|---|---|---|
| Stratum | Activity level | Baseline | Baseline + 10 min brisk | Baseline + 10 min slow running |
| Intensity gradient × age | | | | |
| 45 years | inactive | Reference | 0.78 (0.72–0.84) | 0.75 (0.68–0.82) |
| 45 years | median | 0.34 (0.25–0.46) | 0.30 (0.21–0.42) | 0.30 (0.21–0.42) |
| 60 years | inactive | Reference | 0.88 (0.84–0.93) | 0.86 (0.81–0.92) |
| 60 years | median | 0.62 (0.47–0.81) | 0.60 (0.43–0.84) | 0.60 (0.42–0.85) |
| 75 years | inactive | Reference | 0.95 (0.93–0.97) | 0.95 (0.92–0.97) |
| 75 years | median | 0.88 (0.77–1.00) | 0.90 (0.75–1.08) | 0.90 (0.74–1.09) |
| Volume × age | | | | |
| 45 years | inactive | Reference | 0.83 (0.75–0.93) | 0.62 (0.46–0.83) |
| 45 years | median | 0.20 (0.08–0.47) | 0.19 (0.08–0.45) | 0.18 (0.07–0.44) |
| 60 years | inactive | Reference | 0.89 (0.81–0.98) | 0.73 (0.57–0.94) |
| 60 years | median | 0.31 (0.15–0.64) | 0.29 (0.14–0.63) | 0.27 (0.12–0.61) |
| 75 years | inactive | Reference | 0.92 (0.87–0.98) | 0.81 (0.69–0.94) |
| 75 years | median | 0.35 (0.16–0.75) | 0.32 (0.12–0.80) | 0.27 (0.08–0.93) |
| Intensity gradient × sex | | | | |
| Men | inactive | Reference | 0.94 (0.90–0.98) | 0.93 (0.89–0.97) |
| Men | median | 0.73 (0.58–0.92) | 0.69 (0.51–0.93) | 0.68 (0.50–0.94) |
| Women | inactive | Reference | 0.95 (0.92–0.98) | 0.94 (0.91–0.97) |
| Women | median | 0.76 (0.48–1.20) | 0.72 (0.37–1.43) | 0.72 (0.35–1.48) |
| Volume × sex | | | | |
| Men | inactive | Reference | 0.90 (0.82–0.98) | 0.75 (0.59–0.96) |
| Men | median | 0.33 (0.16–0.67) | 0.31 (0.15–0.66) | 0.29 (0.13–0.65) |
| Women | inactive | Reference | 0.92 (0.88–0.96) | 0.80 (0.71–0.90) |
| Women | median | 0.33 (0.19–0.56) | 0.30 (0.16–0.55) | 0.25 (0.11–0.56) |
| Intensity gradient × WtH | | | | |
| WtH = 0.5 | inactive | Reference | 0.92 (0.87–0.98) | 0.91 (0.85–0.97) |
| WtH = 0.5 | median | 0.67 (0.49–0.90) | 0.62 (0.42–0.92) | 0.61 (0.40–0.92) |
| WtH = 0.6 | inactive | Reference | 0.94 (0.91–0.96) | 0.93 (0.90–0.96) |
| WtH = 0.6 | median | 0.72 (0.58–0.91) | 0.68 (0.49–0.95) | 0.68 (0.48–0.96) |
| WtH = 0.7 | inactive | Reference | 0.96 (0.93–1.00) | 0.96 (0.92–1.00) |
| WtH = 0.7 | median | 0.84 (0.64–1.10) | 0.81 (0.55–1.19) | 0.81 (0.54–1.22) |
| Volume × WtH | | | | |
| WtH = 0.5 | inactive | Reference | 0.80 (0.72–0.90) | 0.56 (0.41–0.75) |
| WtH = 0.5 | median | 0.16 (0.07–0.35) | 0.15 (0.07–0.35) | 0.15 (0.07–0.35) |
| WtH = 0.6 | inactive | Reference | 0.90 (0.85–0.96) | 0.76 (0.65–0.89) |
| WtH = 0.6 | median | 0.36 (0.20–0.67) | 0.35 (0.18–0.68) | 0.32 (0.14–0.72) |
| WtH = 0.7 | inactive | Reference | 0.96 (0.89–1.03) | 0.89 (0.74–1.09) |
| WtH = 0.7 | median | 0.39 (0.18–0.83) | 0.34 (0.14–0.81) | 0.27 (0.09–0.80) |

This table summarises subdistribution hazard ratio (SHR) scenarios from the dose–response models across selected interactions and strata. For each activity example (inactive, defined as 14 mg and -2.9, and median, defined as 33 mg and -2.669 for volume and intensity, respectively), SHRs are shown for the baseline value, a scenario of adding 10 minutes of brisk walking, and a scenario of adding 10 minutes of slow running. The baseline × inactive combination serves as the Reference (SHR = 1.00). Abbreviations: WtH, waist-to-height ratio; SHR, subdistribution hazard ratio; 95% CI, 95% confidence interval.

### 8 Sensitivity analyses

#### 8.1 Landmark analyses beyond 24 months

We repeated all main models after censoring deaths occurring within the first two years of follow-up (landmark analysis) to minimise potential reverse causation (n = 8,422; 216 CVD deaths). Estimates were pooled across the five imputed datasets.

##### 8.1.1 Joint association of physical activity and age with CVD mortality

Figure 21 shows the results of the landmarking model for the interaction between physical activity and age.

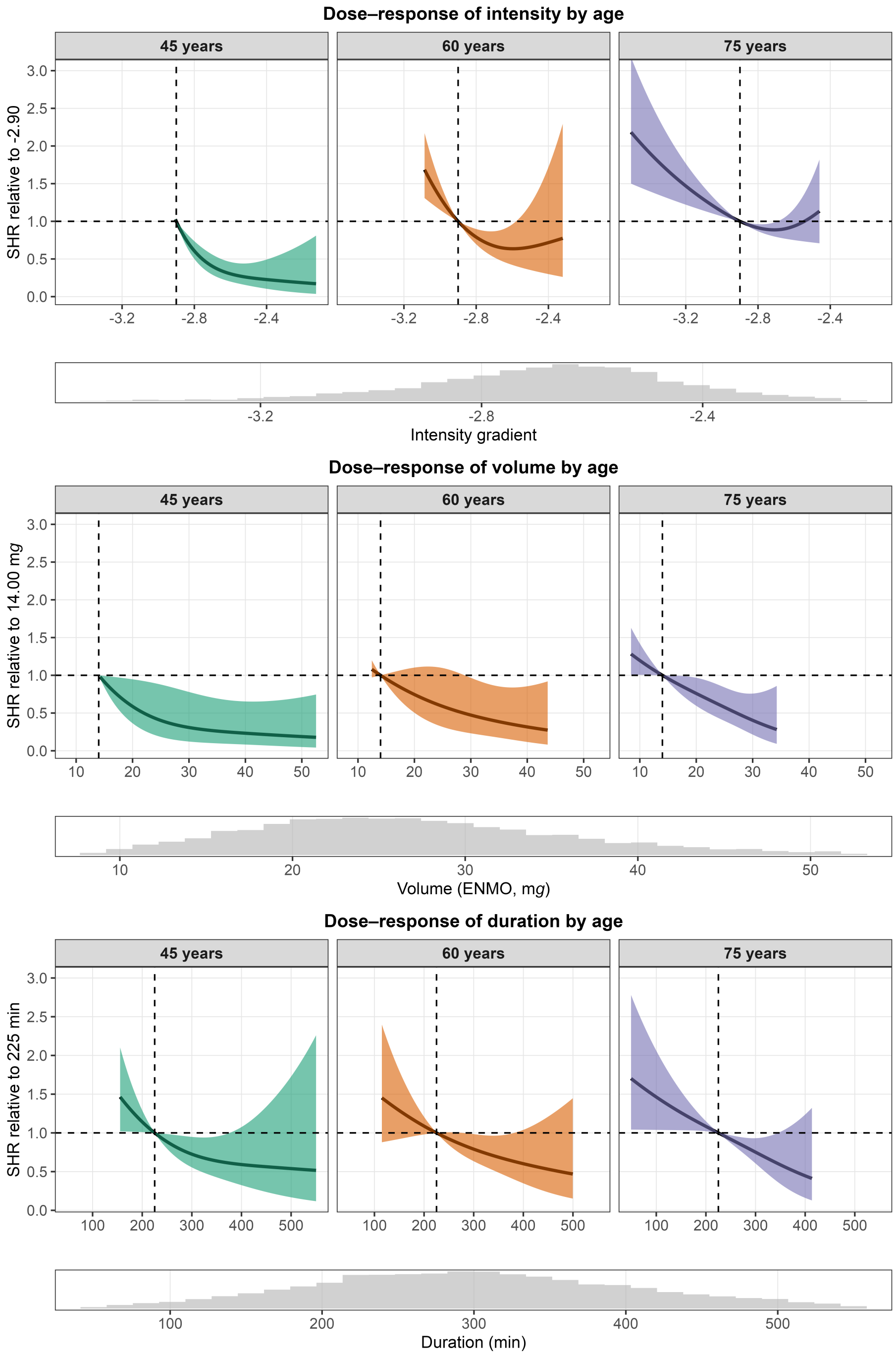

**FIG 21** Landmark model (≥ 24 months follow-up): Dose-response relationship between physical activity intensity (top), volume (middle), and duration (bottom) and cardiovascular disease mortality risk, predicted for ages 45, 60, and 75 years. Models included an interaction term between each physical activity metric and age. Histograms depict the distribution of the corresponding exposure variable.

###### 8.1.2 Joint association of physical activity and sex with CVD mortality

Figure 22 shows the results of the landmarking model for the interaction between physical activity and sex.

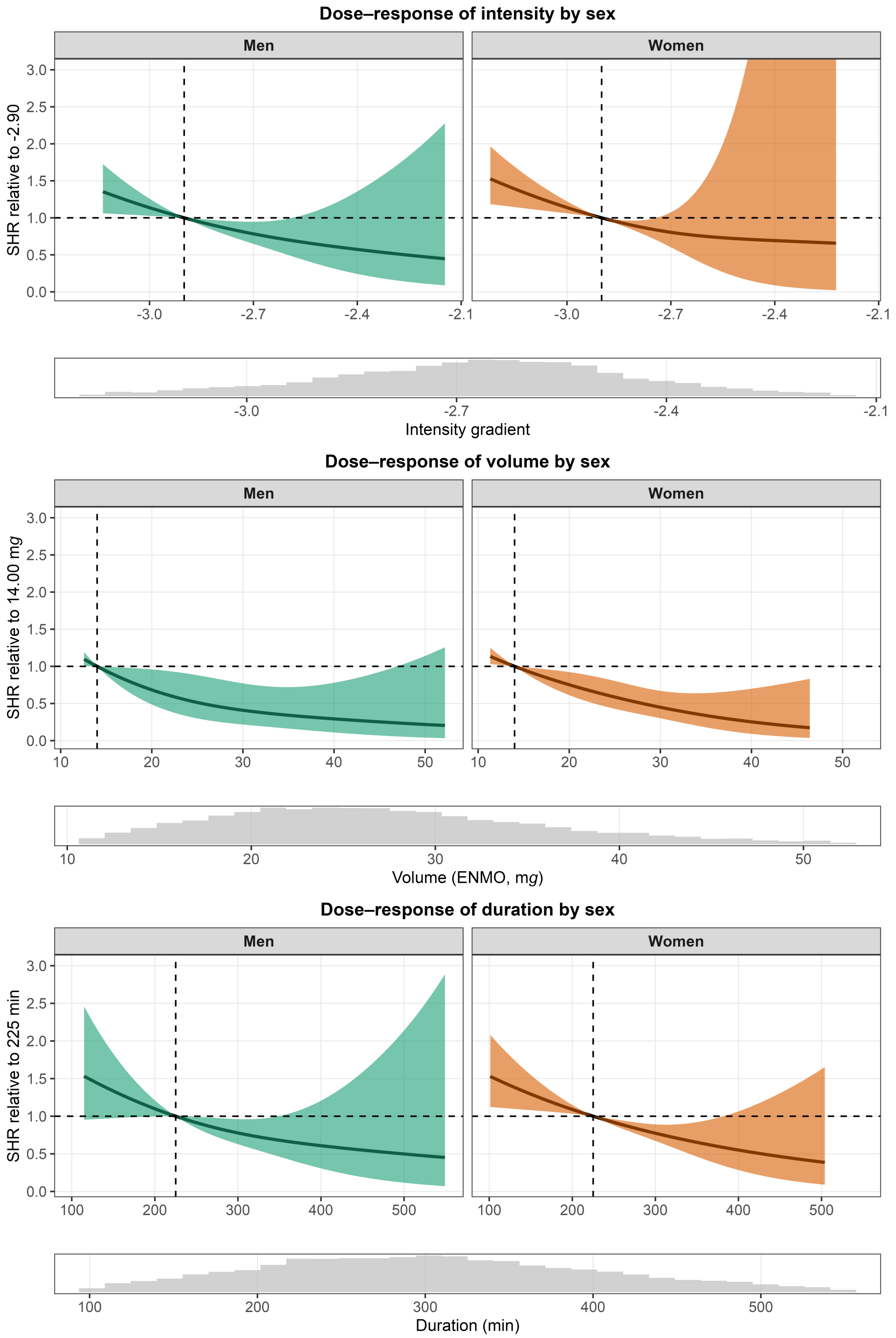

**FIG 22** Landmark model ($\geq$ 24 months follow-up): Dose-response relationship between physical activity intensity (top), volume (middle), and duration (bottom) and cardiovascular disease mortality risk, predicted for men and women. Models included an interaction term between each physical activity metric and sex. Histograms depict the distribution of the corresponding exposure variable.

###### 8.1.3 Joint association of physical activity and body morphology with CVD mortality

Figure 23 shows the results of the landmarking model for the interaction between physical activity and waist-to-height ratio.

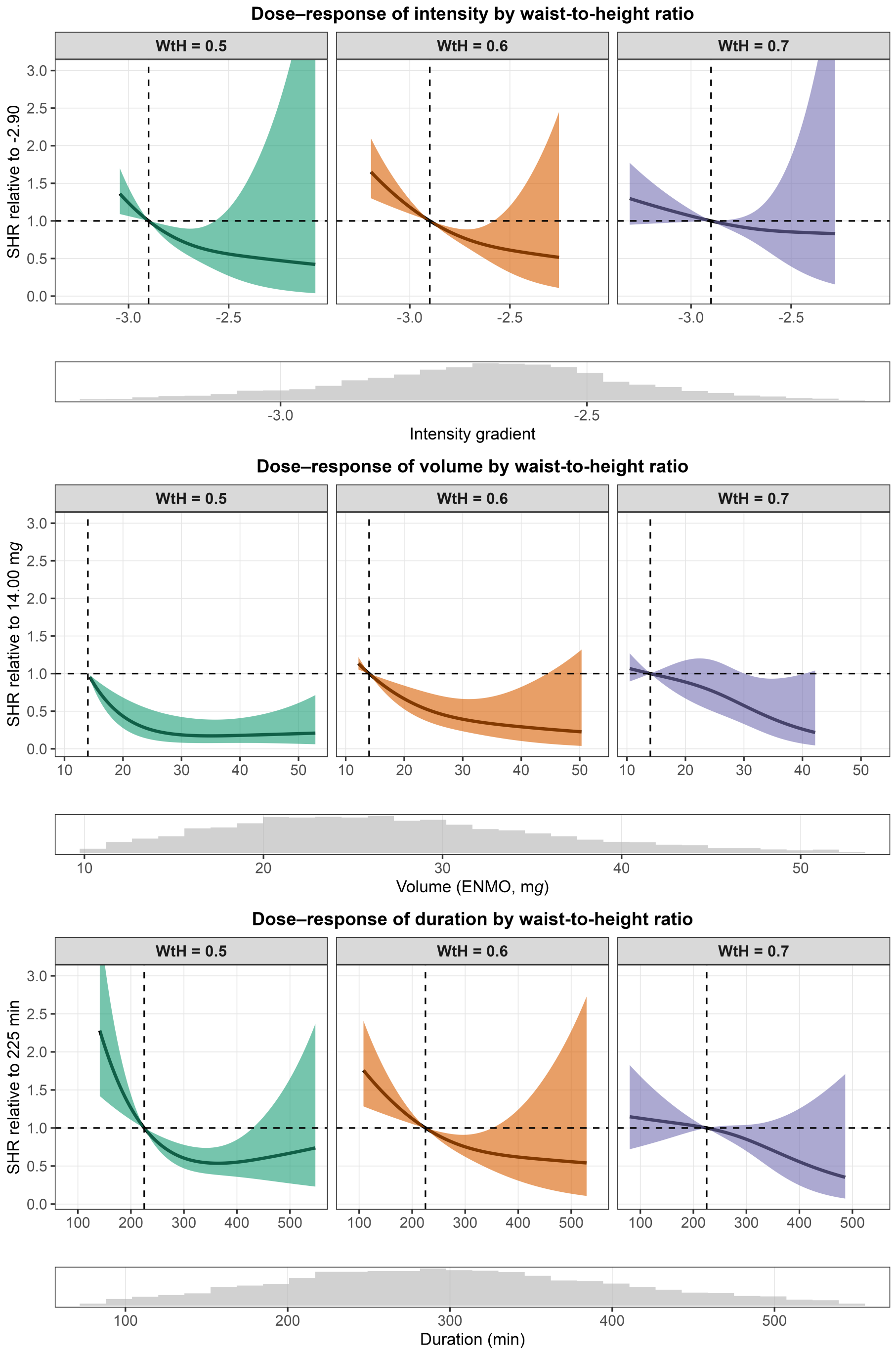

**FIG 23** Landmark model ($\geq$ 24 months follow-up): Dose-response relationship between physical activity intensity (top), volume (middle), and duration (bottom) and cardiovascular disease mortality risk, predicted for waist-to-height ratio (WtH) 0.5, 0.6, and 0.7. Models included an interaction term between each physical activity metric and waist-to-height ratio. Histograms depict the distribution of the corresponding exposure variable.

Figure 24 shows the results of the landmarking model for the interaction between physical activity and waist circumference.

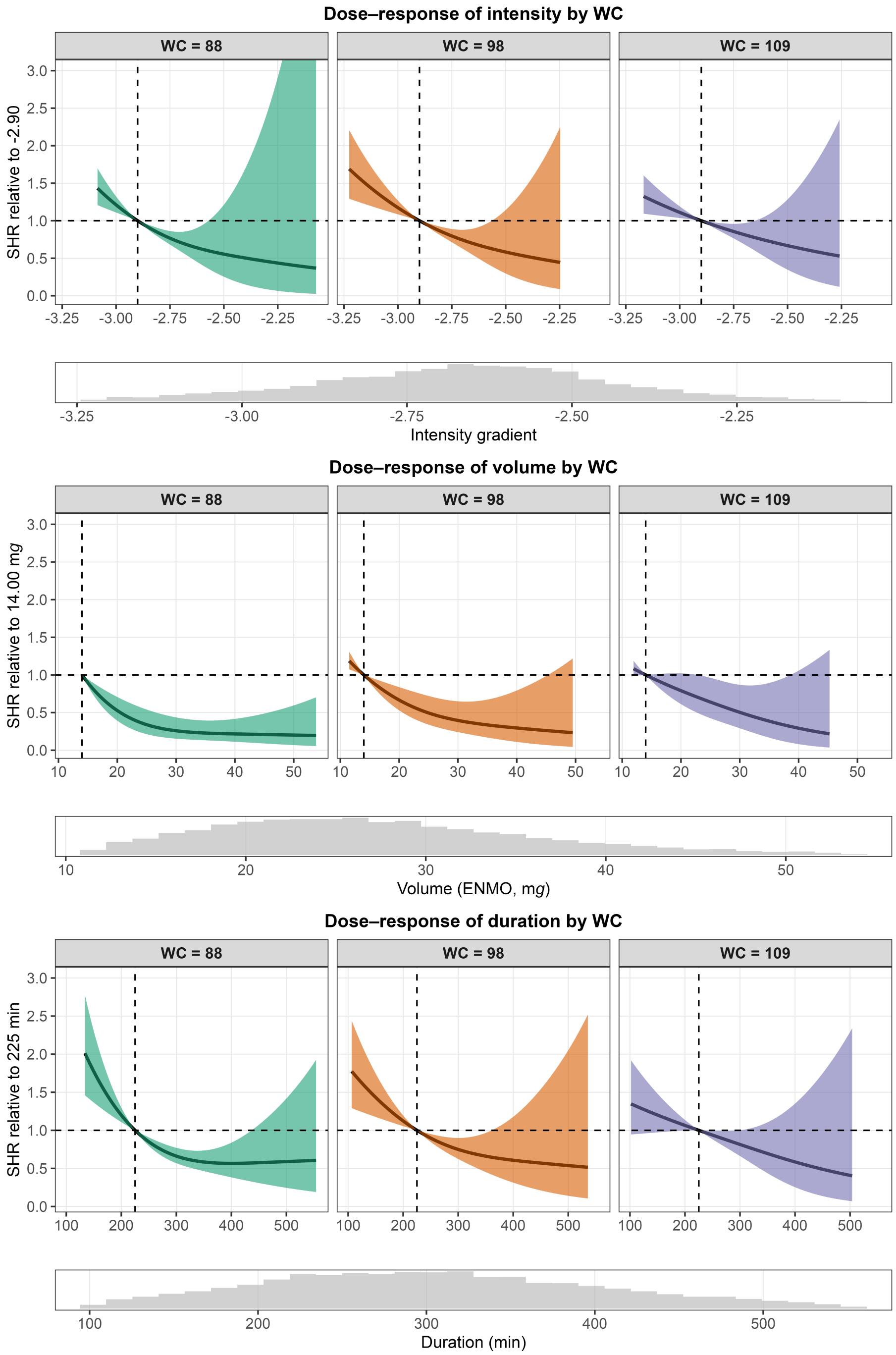

**FIG 24** Landmark model ($\geq$ 24 months follow-up): Dose-response relationship between physical activity intensity (top), volume (middle), and duration (bottom) and cardiovascular disease mortality risk, predicted for waist circumferences (WC) 88, 98, and 109 cm. Models included an interaction term between each physical activity metric and waist circumference. Histograms depict the distribution of the corresponding exposure variable.

#### 8.2 Excluding adults with prevalent cardiovascular disease

We also repeated all models after excluding all adults with prevalent cardiovascular disease to minimise potential reverse causation. Estimates were pooled across the five imputed datasets. Because prevalent CVD status was imputed for 25 participants, the analytic sample comprised 7,650–7,652 participants with 131 CVD deaths and 402–404 non-CVD deaths, depending on the imputed dataset.

##### 8.2.1 Joint association of physical activity and age with CVD mortality

Figure 25 shows the results for the interaction between physical activity and age without adults with prevalent cardiovascular disease.

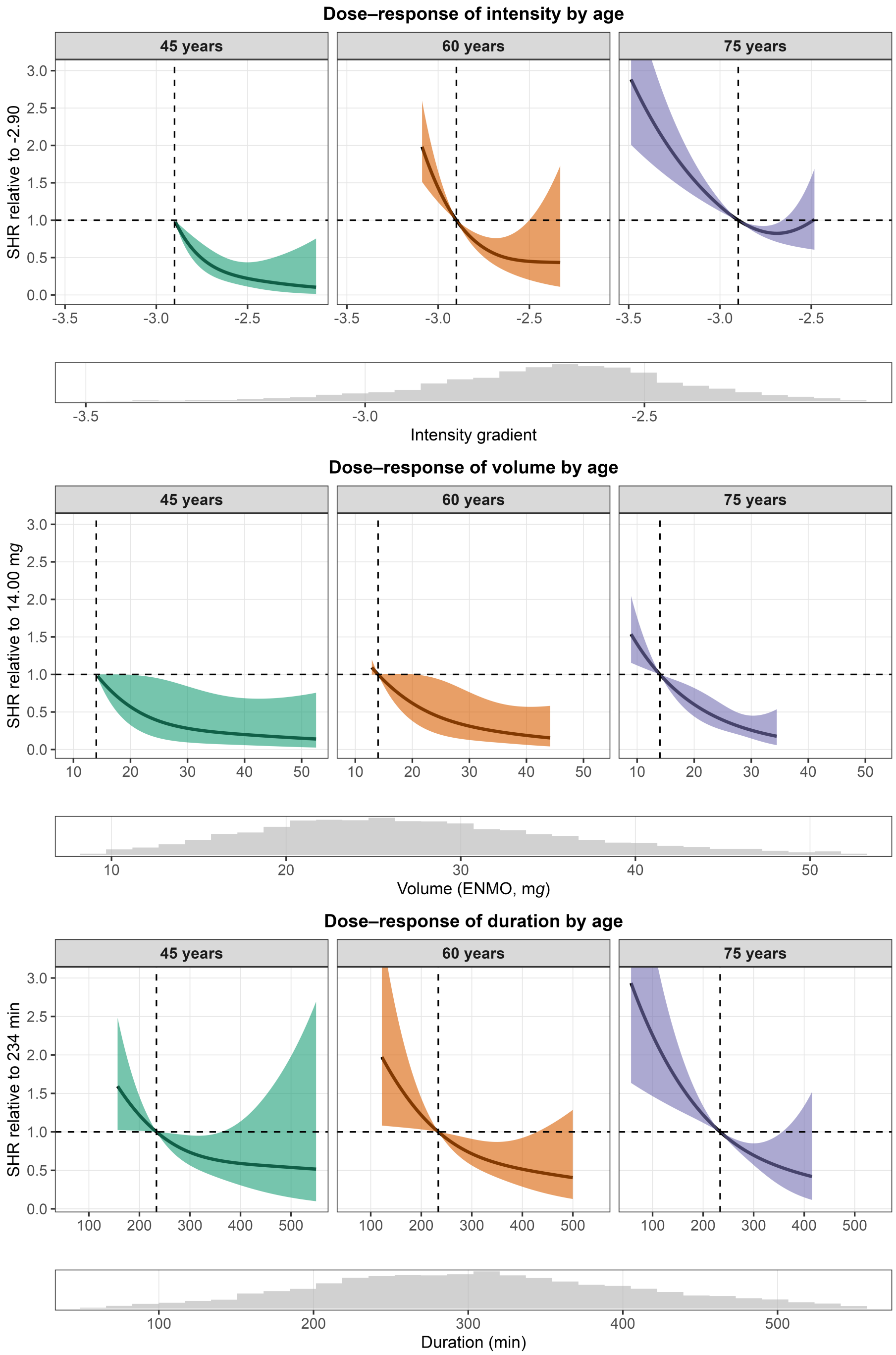

**FIG 25** Without prevalent cardiovascular disease: Dose-response relationship between physical activity intensity (top), volume (middle), and duration (bottom) and cardiovascular disease mortality risk, predicted for ages 45, 60, and 75 years. Models included an interaction term between each physical activity metric and age. Histograms depict the distribution of the corresponding exposure variable.

###### 8.2.2 Joint association of physical activity and sex with CVD mortality

Figure 26 shows the results for the interaction between physical activity and sex without adults with prevalent cardiovascular disease.

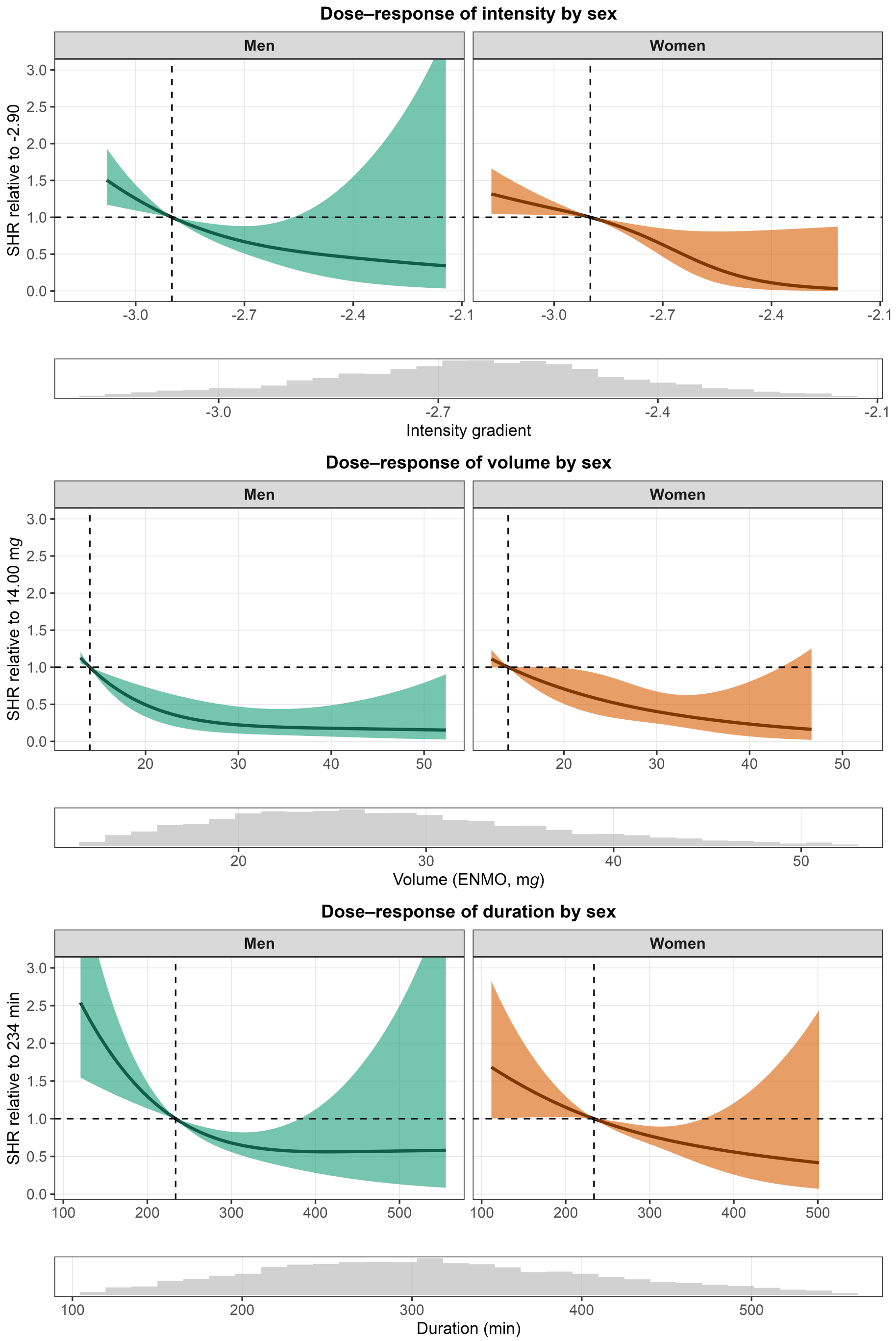

**FIG 26** Without prevalent cardiovascular disease: Dose-response relationship between physical activity intensity (top), volume (middle), and duration (bottom) and cardiovascular disease mortality risk, predicted for men and women. Models included an interaction term between each physical activity metric and sex. Histograms depict the distribution of the corresponding exposure variable.

##### 8.2.3 Joint association of physical activity and body morphology with CVD mortality

Figure 27 shows the results for the interaction between physical activity and waist-to-height ratio without adults with prevalent cardiovascular disease.

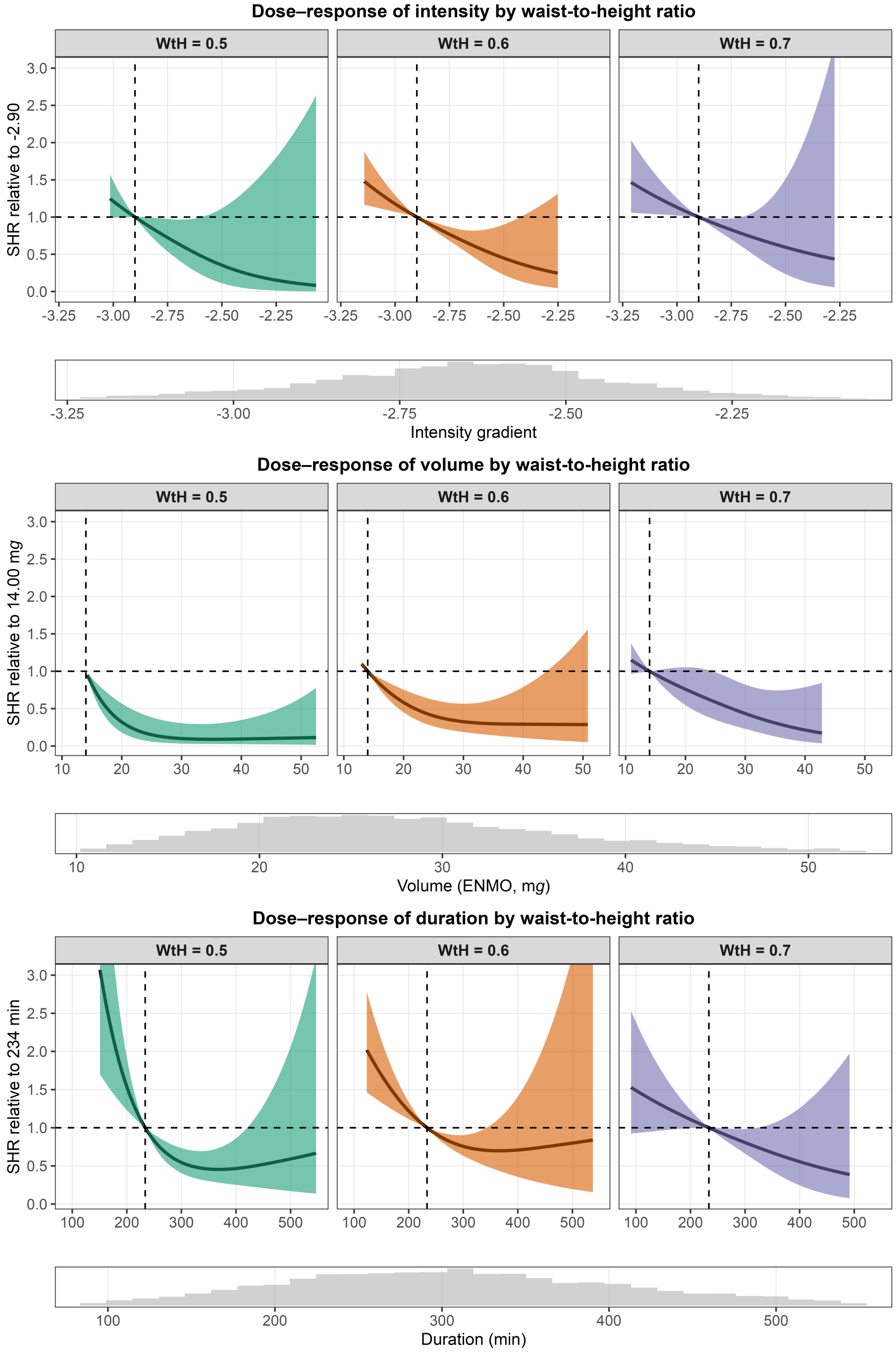

**FIG 27** Without prevalent cardiovascular disease: Dose-response relationship between physical activity intensity (top), volume (middle), and duration (bottom) and cardiovascular disease mortality risk, predicted for waist-to-height ratio (WtH) 0.5, 0.6, and 0.7. Models included an interaction term between each physical activity metric and waist-to-height ratio. Histograms depict the distribution of the corresponding exposure variable.

Figure 28 shows the results for the interaction between physical activity and waist circumference without adults with prevalent cardiovascular disease.

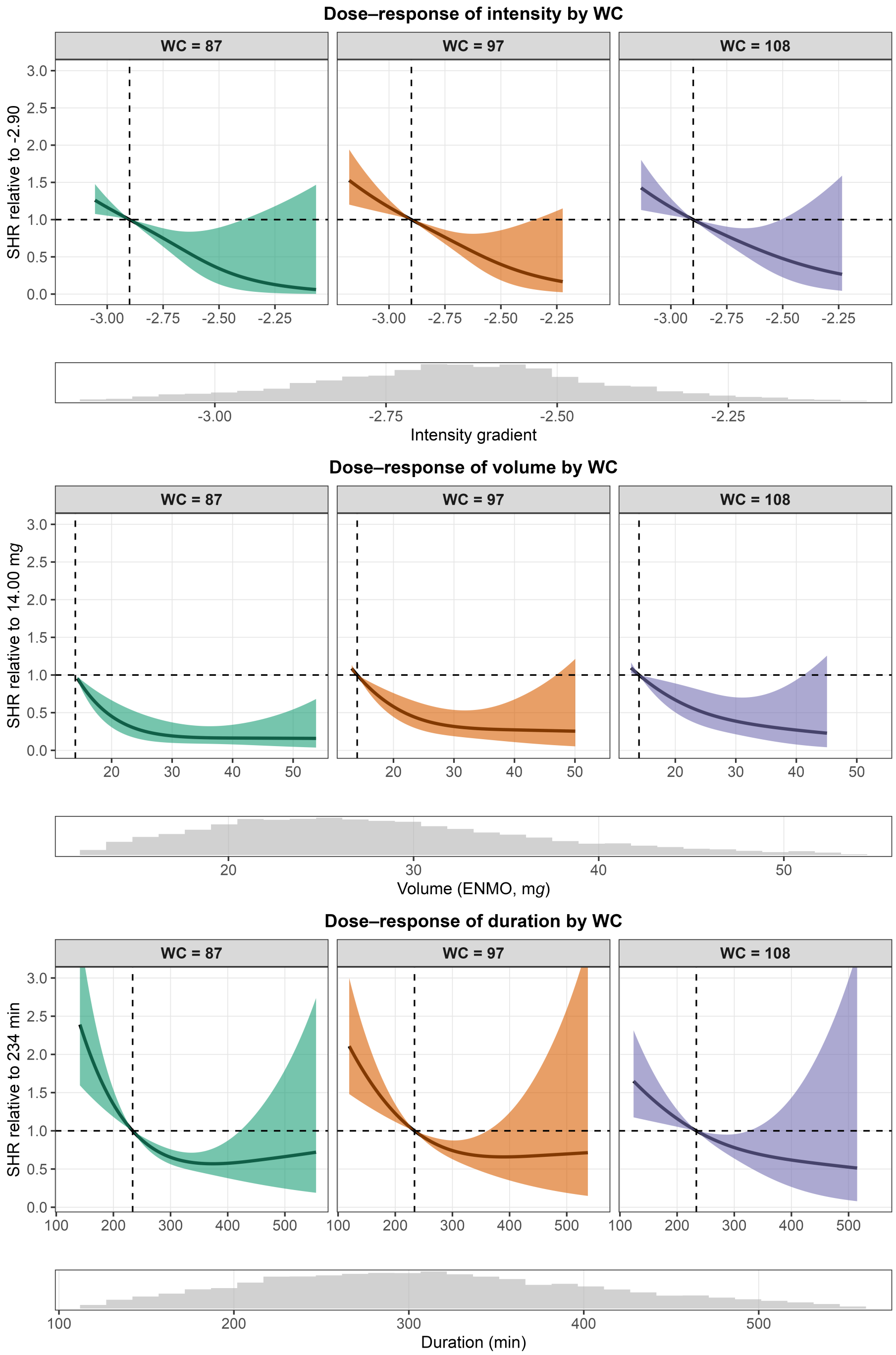

**FIG 28** Without prevalent cardiovascular disease: Dose-response relationship between physical activity intensity (top), volume (middle), and duration (bottom) and cardiovascular disease mortality risk, predicted for waist circumferences (WC) 88, 98, and 109 cm. Models included an interaction term between each physical activity metric and waist circumference. Histograms depict the distribution of the corresponding exposure variable.

#### 8.3 Complete-case analyses

This section presents results of the complete-case analyses (n=7,838, events=226). Models were adjusted for age (continuous), sex (binary), body mass index (continuous), education (categorical), income-to-poverty ratio (continuous), ethnicity (categorical), smoking status (categorical), congestive heart failure (binary), coronary heart disease (binary), myocardial infarction (binary), angina pectoris (binary), stroke (binary), cancer (binary), and diabetes (binary). Contrary to the main imputed models, we did not adjust for mean arterial pressure because of missing data. We included each cardiovascular condition individually into the model.

##### 8.3.1 Joint association of physical activity and age with CVD mortality

Figure 29 shows the complete-case results for the interaction between physical activity and age.

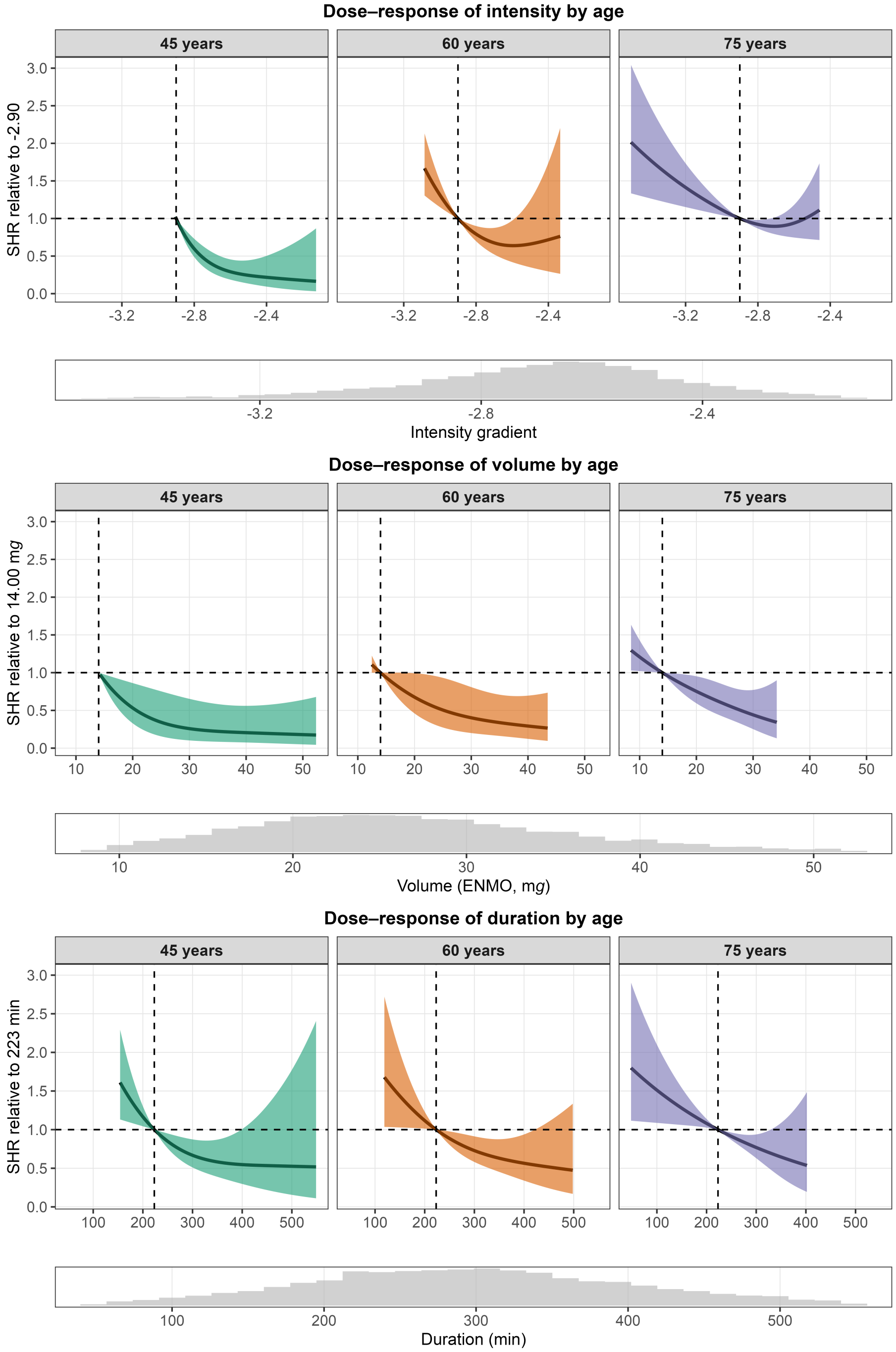

**FIG 29** Complete-case model: Dose-response relationship between physical activity intensity (top), volume (middle), and duration (bottom) and cardiovascular disease mortality risk, predicted for ages 45, 60, and 75 years. Models included an interaction term between each physical activity metric and age. Histograms depict the distribution of the corresponding exposure variable.

##### 8.3.2 Joint association of physical activity and sex with CVD mortality

Figure 30 shows the complete-case results for the interaction between physical activity and sex.

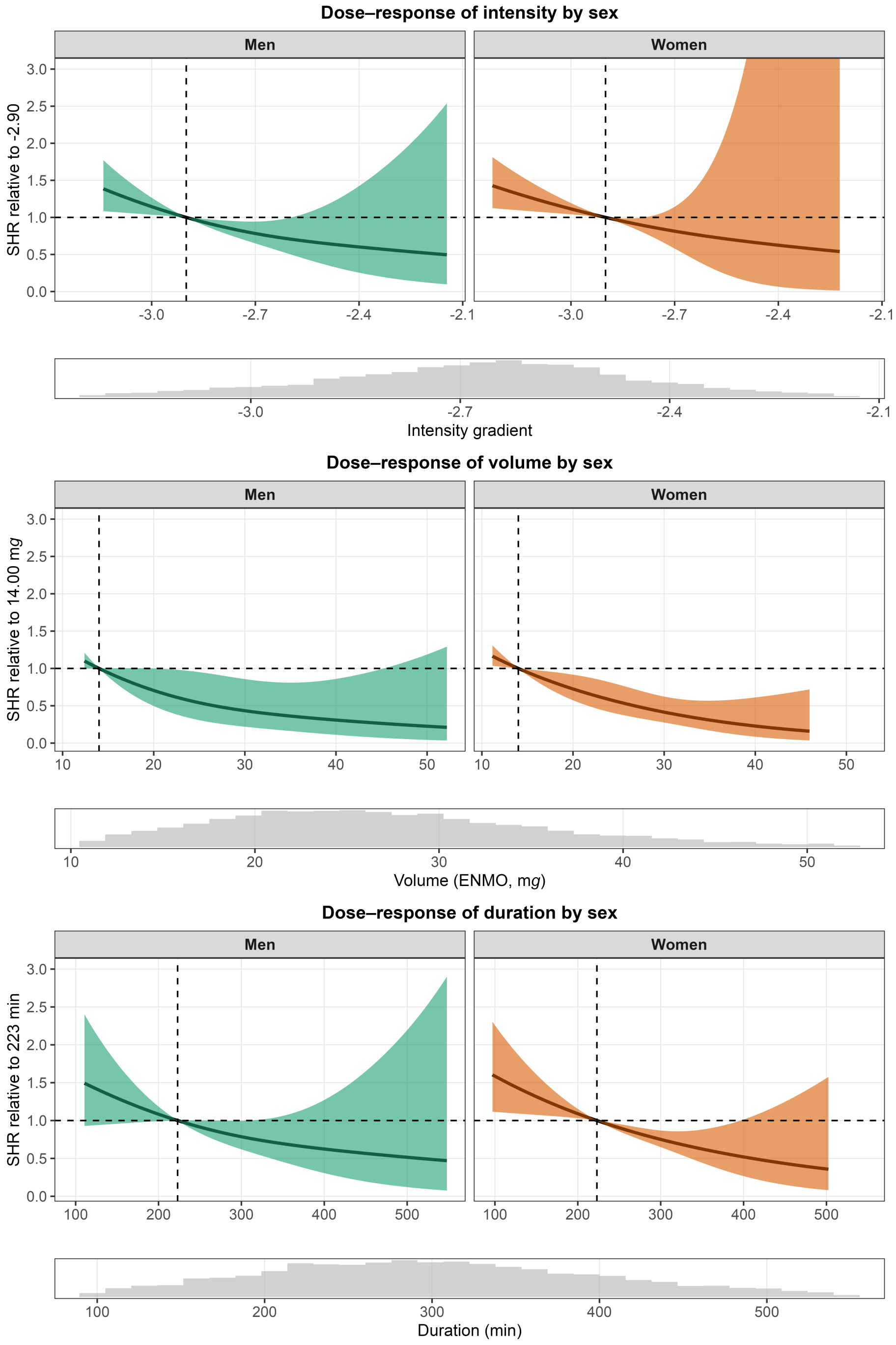

**FIG 30** Complete-case model: Dose-response relationship between physical activity intensity (top), volume (middle), and duration (bottom) and cardiovascular disease mortality risk, predicted for men and women. Models included an interaction term between each physical activity metric and sex. Histograms depict the distribution of the corresponding exposure variable.

##### 8.3.3 Joint association of physical activity and body morphology with CVD mortality

Figure 31 shows the complete-case results for the interaction between physical activity and waist-to-height ratio.

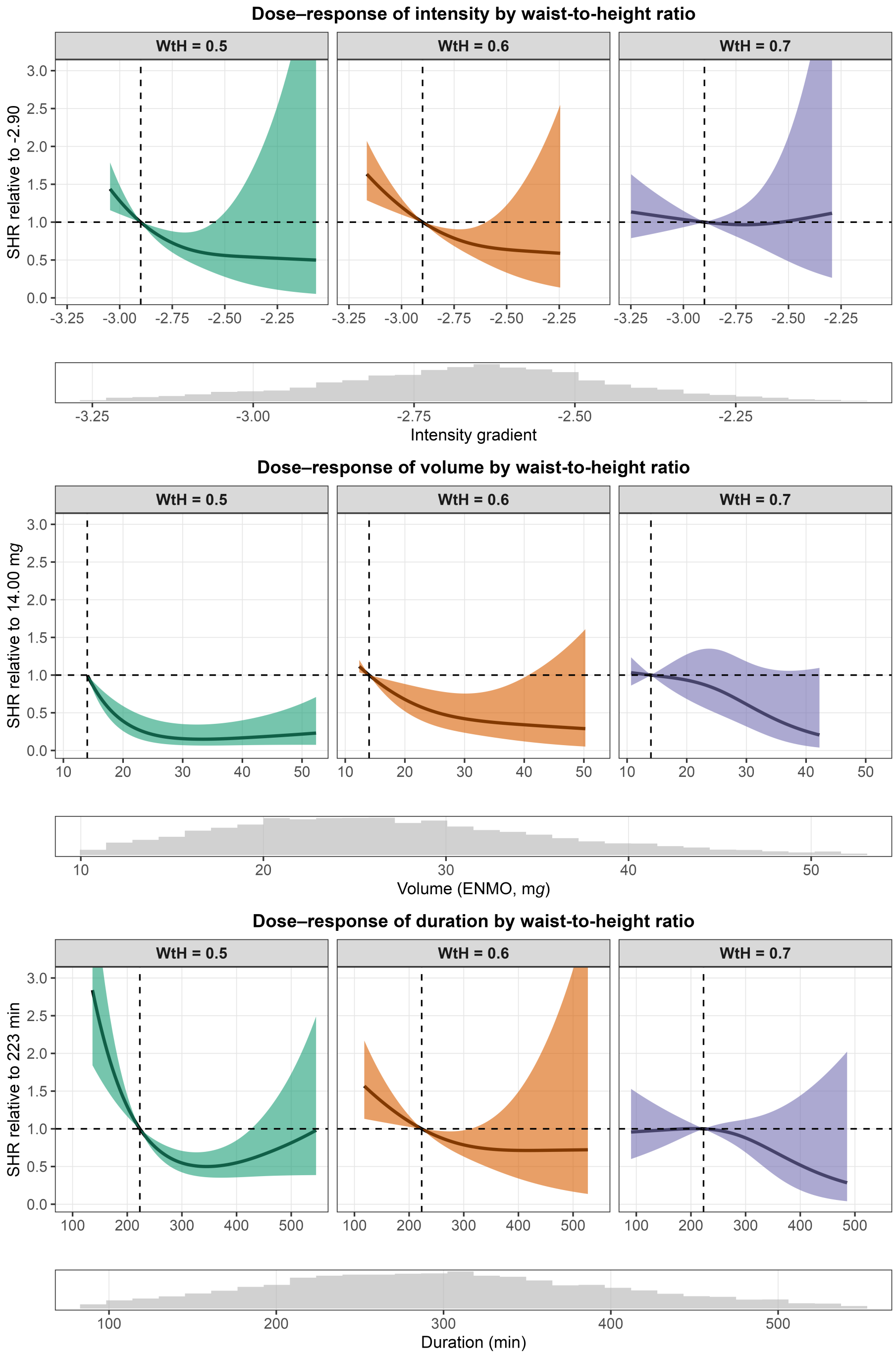

**FIG 31** Complete-case model: Dose-response relationship between physical activity intensity (top), volume (middle), and duration (bottom) and cardiovascular disease mortality risk, predicted for waist-to-height ratio (WtH) 0.5, 0.6, and 0.7. Models included an interaction term between each physical activity metric and WtH. Histograms depict the distribution of the corresponding exposure variable. These analyses were not adjusted for body mass index.

Figure 32 shows the complete-case results for the interaction between physical activity and waist circumference.

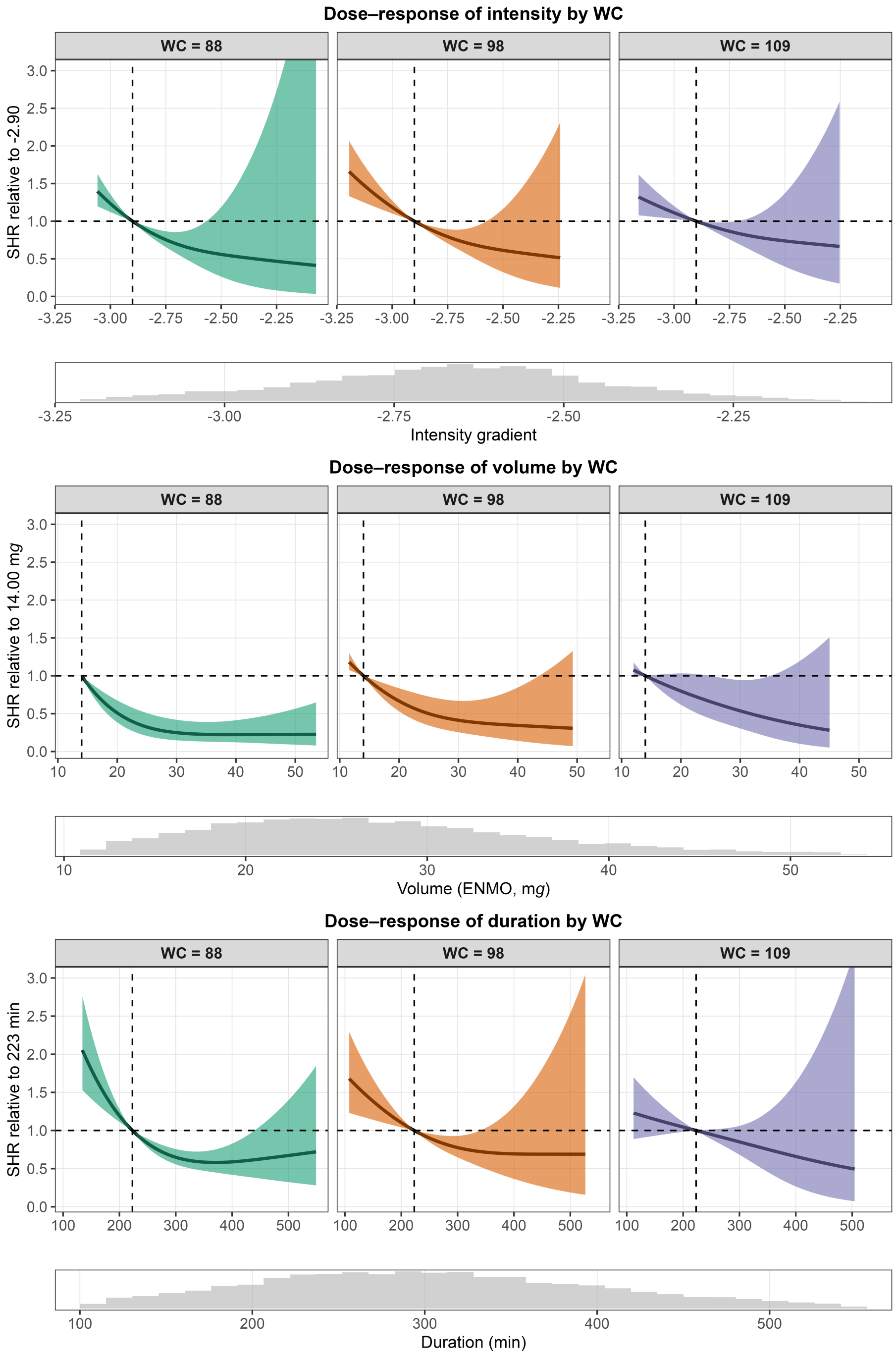

**FIG 32** Complete-case model: Dose-response relationship between physical activity intensity (top), volume (middle), and duration (bottom) and cardiovascular disease mortality risk, predicted for waist circumferences (WC) 88, 98, and 109 cm. Models included an interaction term between each physical activity metric and waist circumference. Histograms depict the distribution of the corresponding exposure variable. These analyses were not adjusted for body mass index.

163 Figure 33 shows the complete-case results for the interaction between physical activity and body mass index.

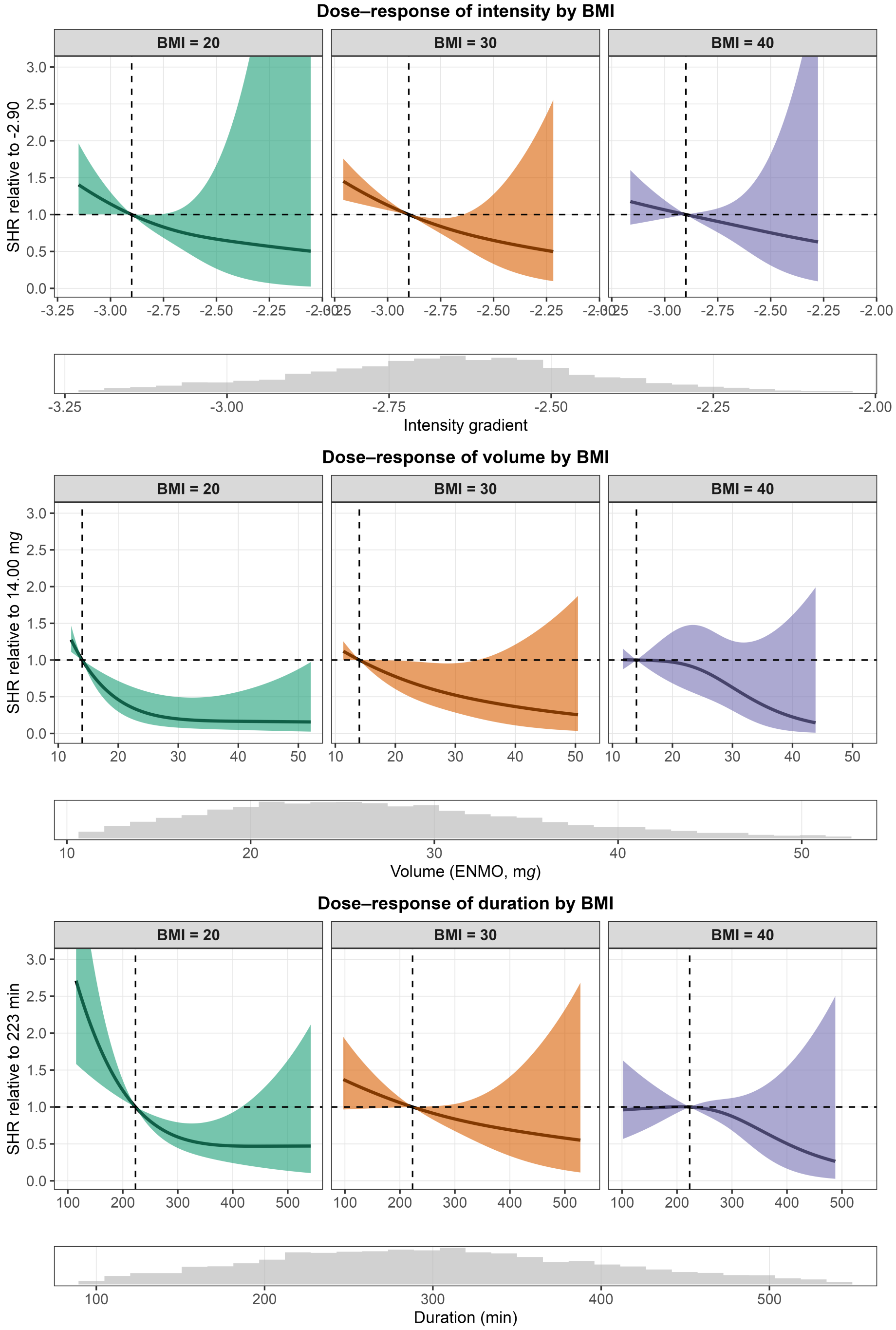

**FIG 33** Complete-case model: Dose-response relationship between physical activity intensity (top), volume (middle), and duration (bottom) and cardiovascular disease mortality risk, predicted for body mass index (BMI) 20, 30, and 40 kg/m$^2$. Models included an interaction term between each physical activity metric and BMI. Histograms depict the distribution of the corresponding exposure variable.

#### 8.4 Unadjusted for blood pressure and diabetes status

We repeated all main analyses, pooled across the five imputed datasets, without adjusting for mean arterial pressure and diabetes status. This was done because these variables might be on the causal pathway of physical activity and mortality risk. Overall, the sensitivity analyses were similar to the main analyses presented in the manuscript across all models. This suggests that additional adjustment for blood pressure and diabetes status might not have biased estimates of our main analyses.

##### 8.4.1 Joint association of physical activity and age with CVD mortality

Figure 34 shows the results of this sensitivity analysis for the interaction between physical activity and age.

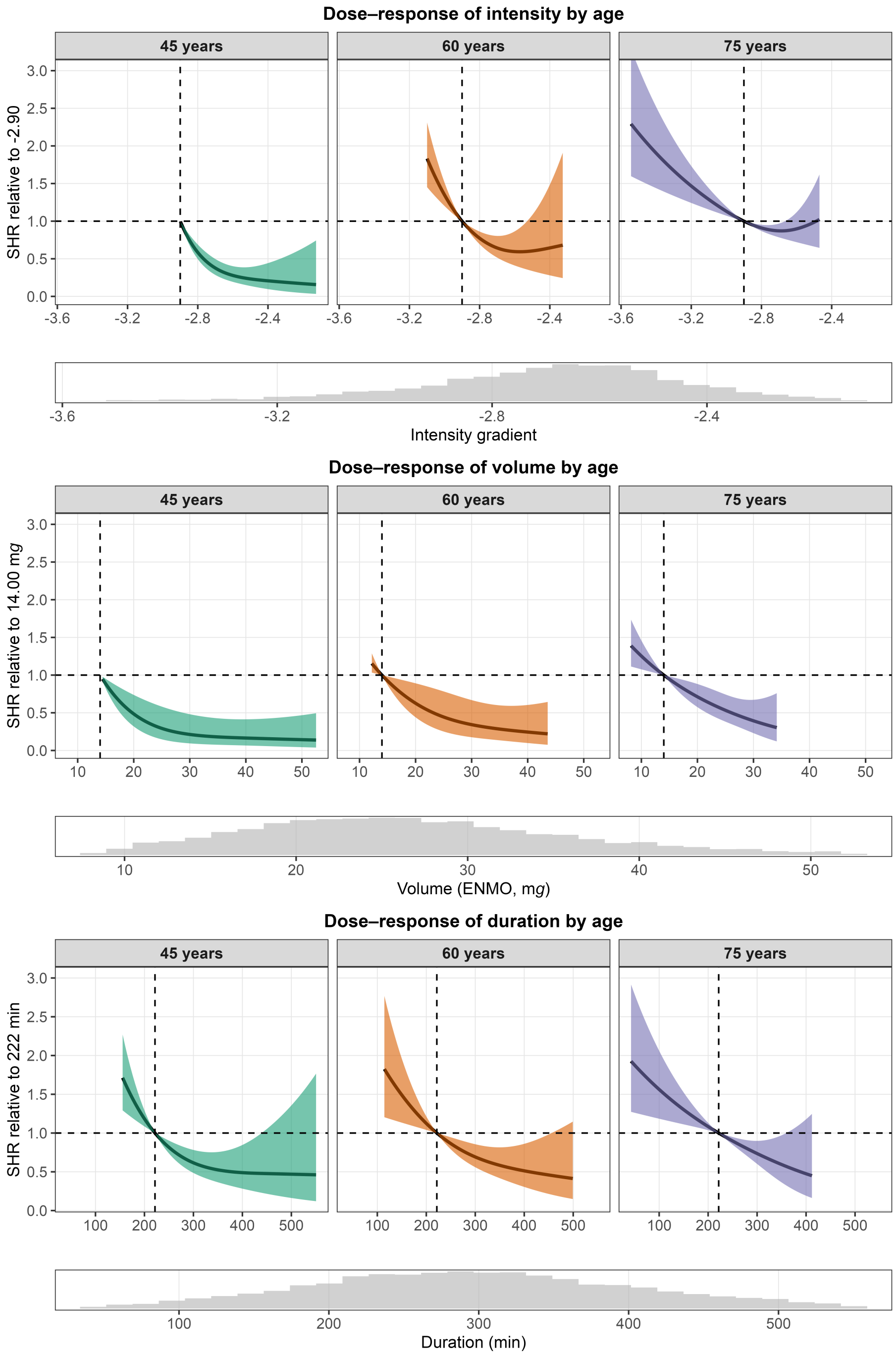

**FIG 34** Without adjusting for mean arterial pressure and diabetes status: Dose-response relationship between physical activity intensity (top), volume (middle), and duration (bottom) and cardiovascular disease mortality risk, predicted for ages 45, 60, and 75 years. Models included an interaction term between each physical activity metric and age. Histograms depict the distribution of the corresponding exposure variable.

###### 8.4.2 Joint association of physical activity and sex with CVD mortality

Figure 35 shows the results for the interaction between physical activity and sex.

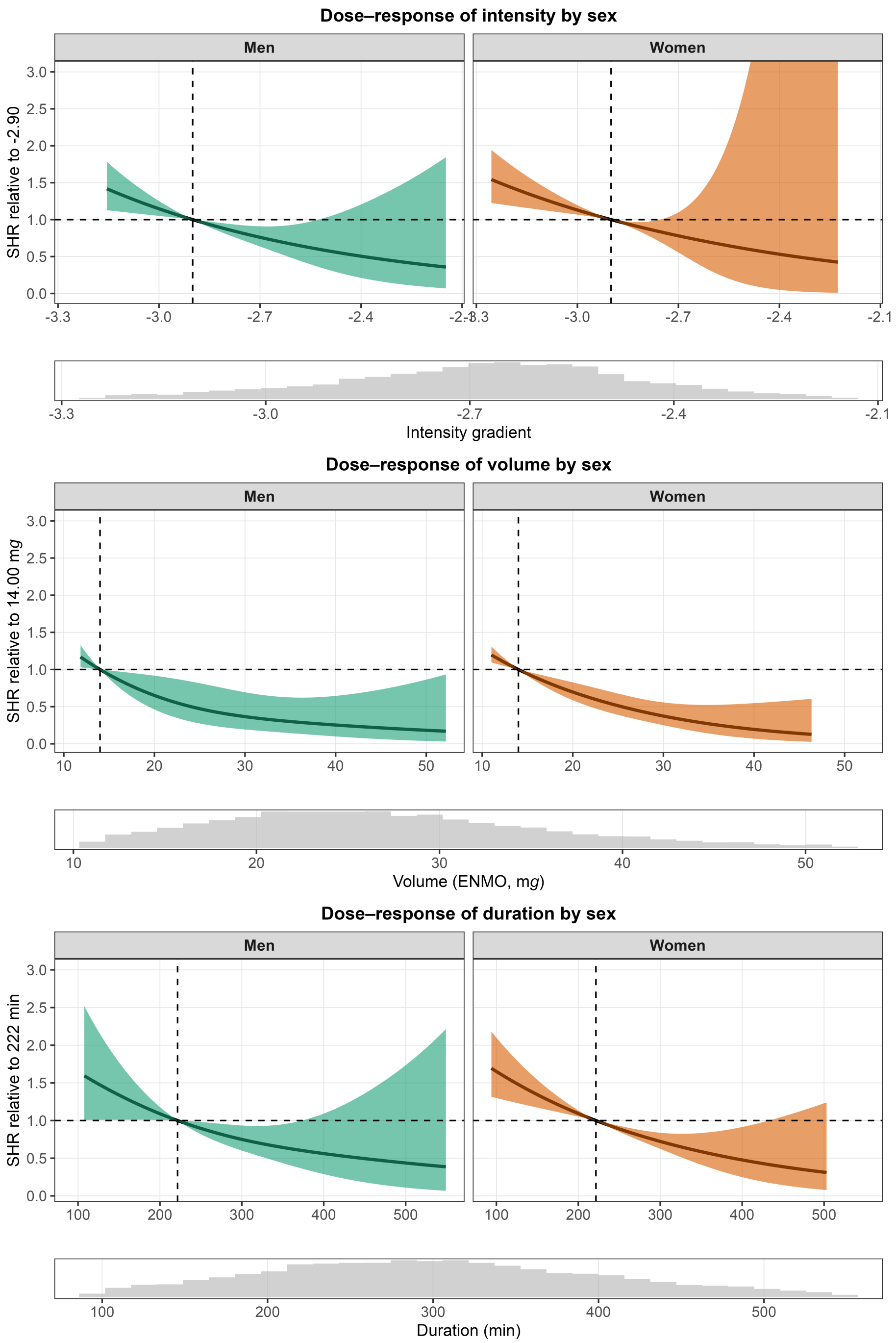

**FIG 35** Without adjusting for mean arterial pressure and diabetes status: Dose-response relationship between physical activity intensity (top), volume (middle), and duration (bottom) and cardiovascular disease mortality risk, predicted for men and women. Models included an interaction term between each physical activity metric and sex. Histograms depict the distribution of the corresponding exposure variable.

##### 8.4.3 Joint association of physical activity and body morphology with CVD mortality

Figure 36 shows the results for the interaction between physical activity and waist-to-height ratio.

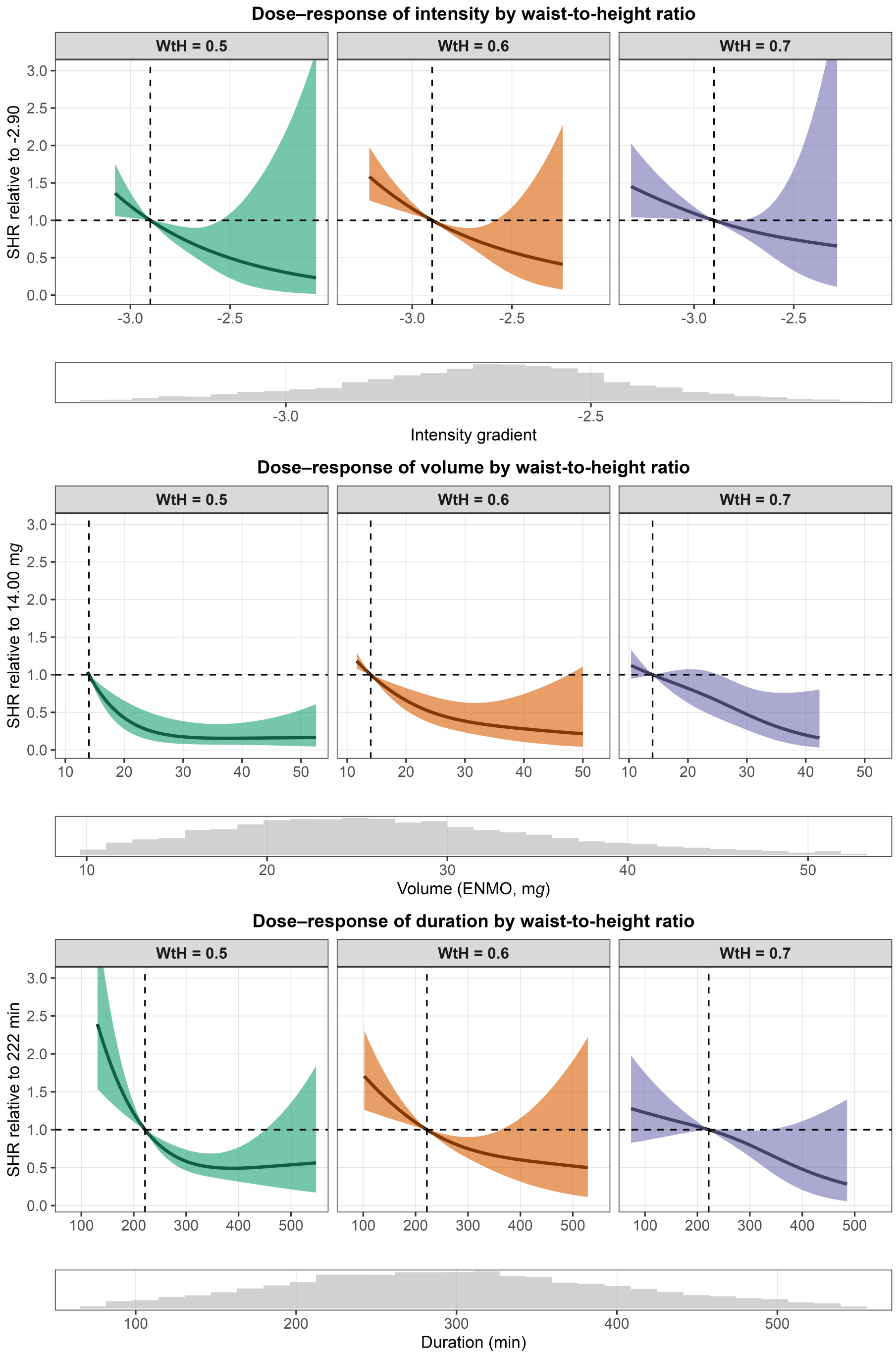

**FIG 36** Without adjusting for mean arterial pressure and diabetes status: Dose-response relationship between physical activity intensity (top), volume (middle), and duration (bottom) and cardiovascular disease mortality risk, predicted for waist-to-height ratio (WtH) 0.5, 0.6, and 0.7. Models included an interaction term between each physical activity metric and WtH. Histograms depict the distribution of the corresponding exposure variable.

Figure 37 shows the results for the interaction between physical activity and waist circumference.

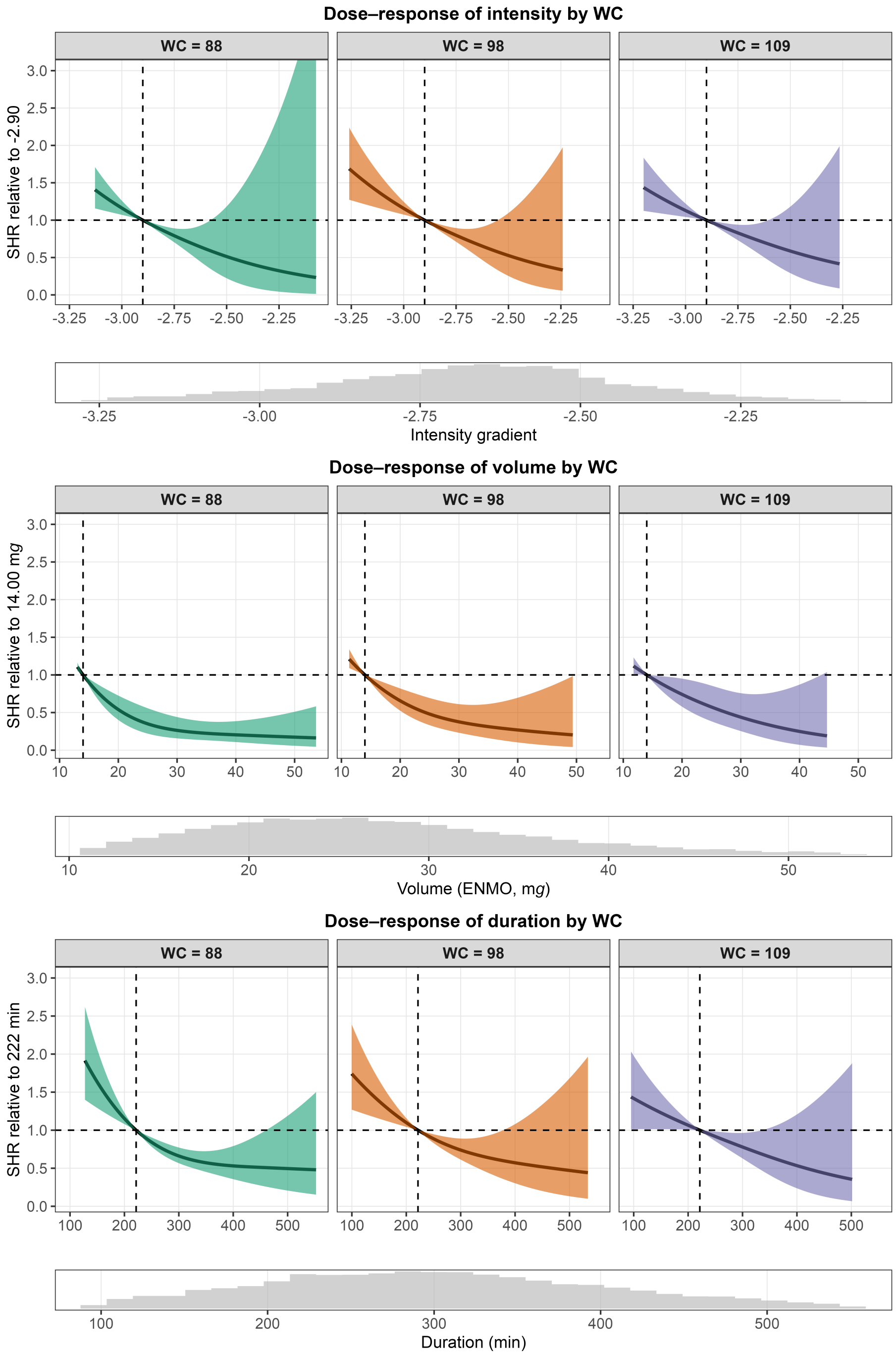

**FIG 37** Without adjusting for mean arterial pressure and diabetes status: Dose-response relationship between physical activity intensity (top), volume (middle), and duration (bottom) and cardiovascular disease mortality risk, predicted for waist circumferences (WC) 88, 98, and 109 cm. Models included an interaction term between each physical activity metric and waist circumference. Histograms depict the distribution of the corresponding exposure variable.

Figure 38 shows the results for the interaction between physical activity and body mass index.

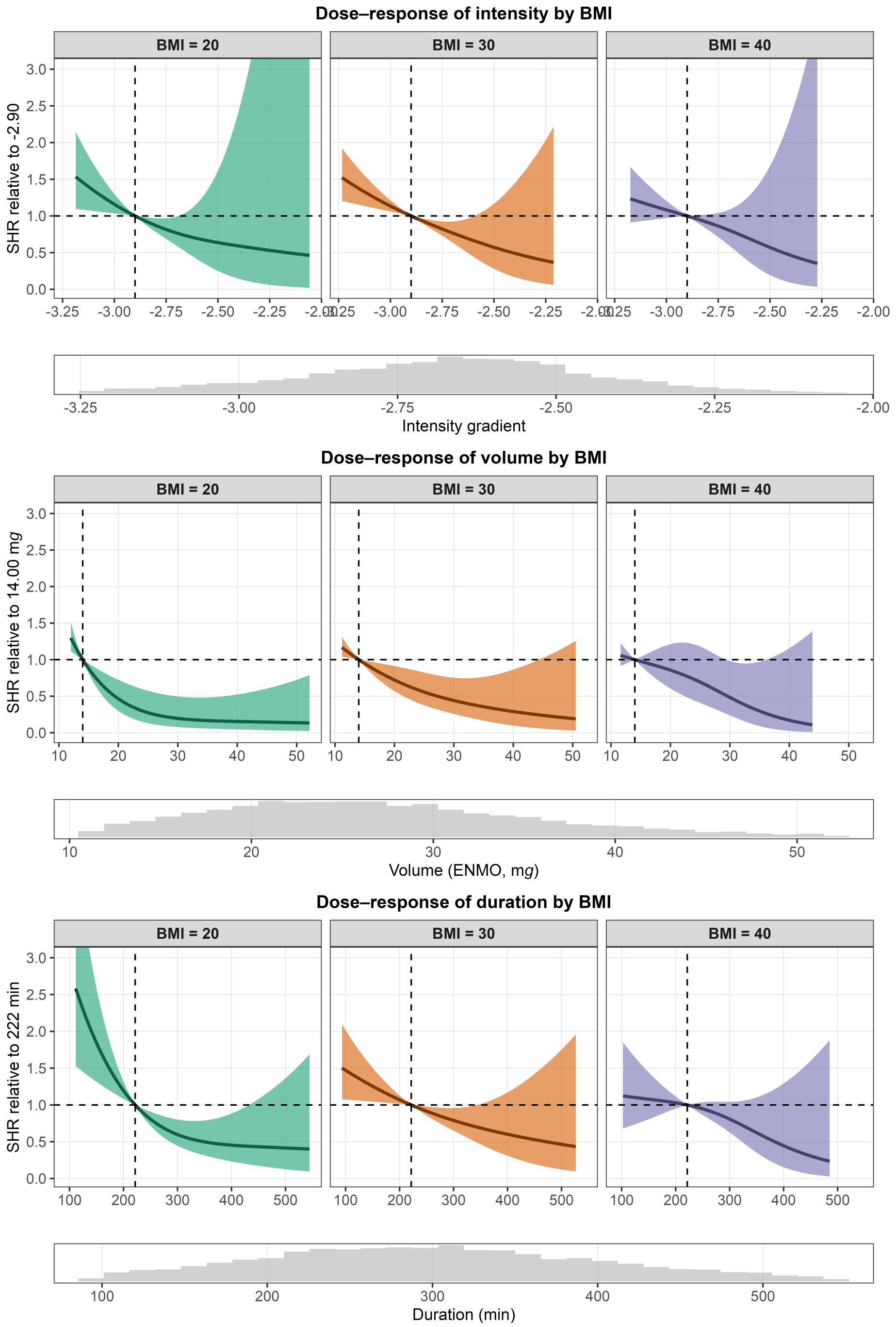

**FIG 38** Without adjusting for mean arterial pressure and diabetes status: Dose-response relationship between physical activity intensity (top), volume (middle), and duration (bottom) and cardiovascular disease mortality risk, predicted for body mass index (BMI) 20, 30, and 40 kg/m$^2$. Models included an interaction term between each physical activity metric and BMI. Histograms depict the distribution of the corresponding exposure variable.

#### 8.5 Using all-cause mortality as outcome

We repeated all main models using all-cause mortality as outcome. We included 8,607 participants of whom 847 died during a median follow-up of 81 (interquartile range: 69–94) months. Models were adjusted for the same covariates as in the main analyses.

An overview of the models together with test results for added value of the interaction term are provided in Table 7.

**TABLE 7** Diagnostics and interaction test results for all-cause mortality models.

| PA metric | Association modifier | All-cause events | Parameters | Events/param. | Interaction p-value |
|---|---|---|---|---|---|
| Intensity | Age | 847 | 23 | 36.8 | 0.031 |
| Volume | Age | 847 | 23 | 36.8 | 0.440 |
| Duration | Age | 847 | 23 | 36.8 | 0.357 |
| Intensity | Sex | 847 | 22 | 38.5 | 0.979 |
| Volume | Sex | 847 | 22 | 38.5 | 0.345 |
| Duration | Sex | 847 | 22 | 38.5 | 0.830 |
| Intensity | Waist-to-height ratio | 847 | 23 | 36.8 | 0.013 |
| Volume | Waist-to-height ratio | 847 | 23 | 36.8 | 0.029 |
| Duration | Waist-to-height ratio | 847 | 23 | 36.8 | 0.021 |
| Intensity | Body mass index | 847 | 23 | 36.8 | 0.159 |
| Volume | Body mass index | 847 | 23 | 36.8 | 0.042 |
| Duration | Body mass index | 847 | 23 | 36.8 | 0.043 |
| Intensity | Waist circumference | 847 | 23 | 36.8 | 0.034 |
| Volume | Waist circumference | 847 | 23 | 36.8 | 0.035 |
| Duration | Waist circumference | 847 | 23 | 36.8 | 0.025 |

##### 8.5.1 Joint association of physical activity and age with all-cause mortality

Figure 39 shows the results for the interaction between physical activity and age with all-cause mortality as outcome. We observed similar dose-response patterns in intensity, volume, and duration across age. Unlike for CVD mortality, the association with intensity among older adults did not level off at higher intensities.

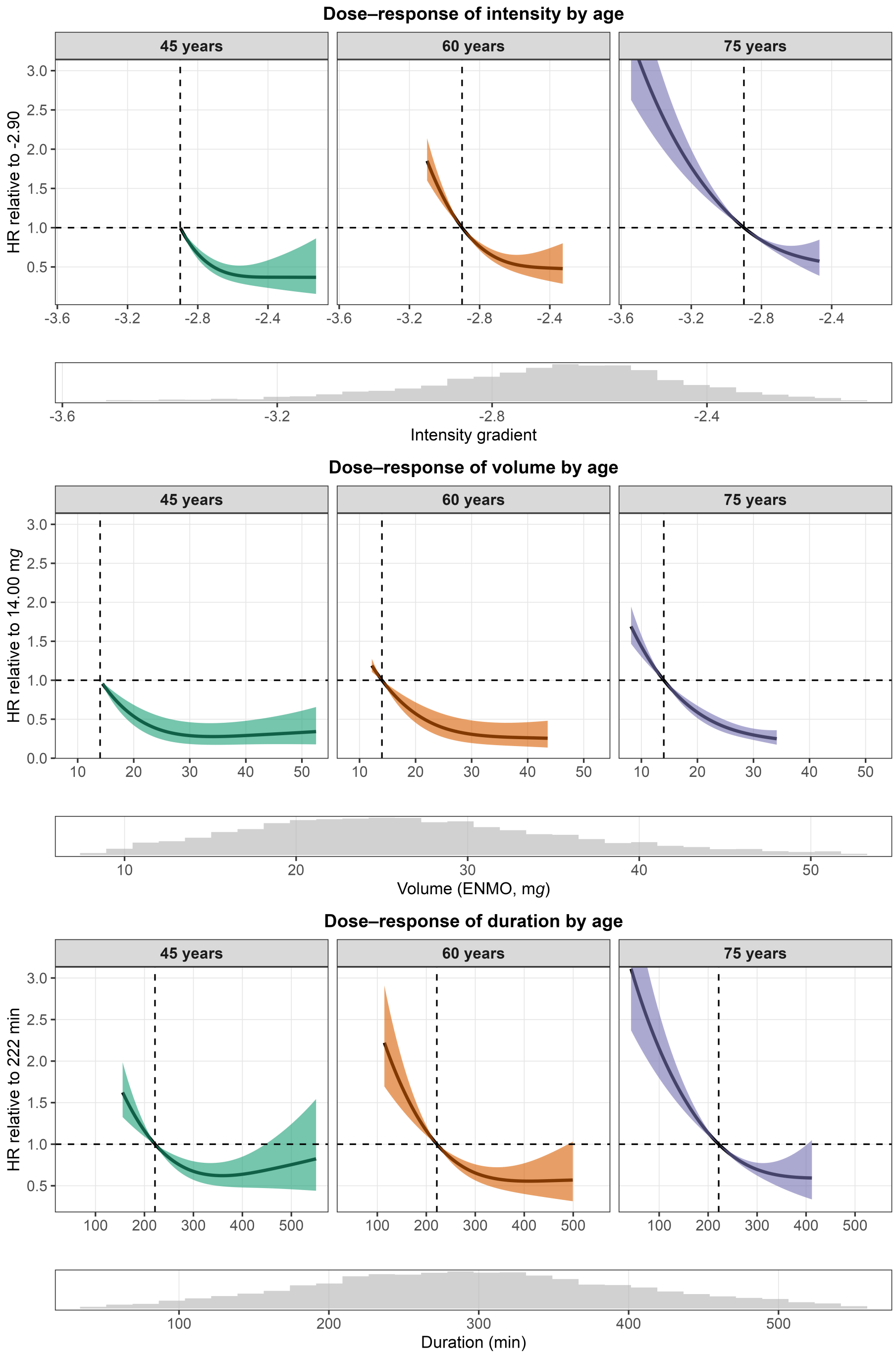

**FIG 39** All-cause mortality: Dose-response relationship between physical activity intensity (top), volume (middle), and duration (bottom) and all-cause mortality risk, predicted for ages 45, 60, and 75 years. Models included an interaction term between each physical activity metric and age. Histograms depict the distribution of the corresponding exposure variable.

##### 8.5.2 Joint association of physical activity and sex with all-cause mortality

Figure 40 shows the results for the interaction between physical activity and sex with all-cause mortality as outcome. Compared to dose-response curves for CVD mortality, those for all-cause mortality were similar with narrower confidence intervals.

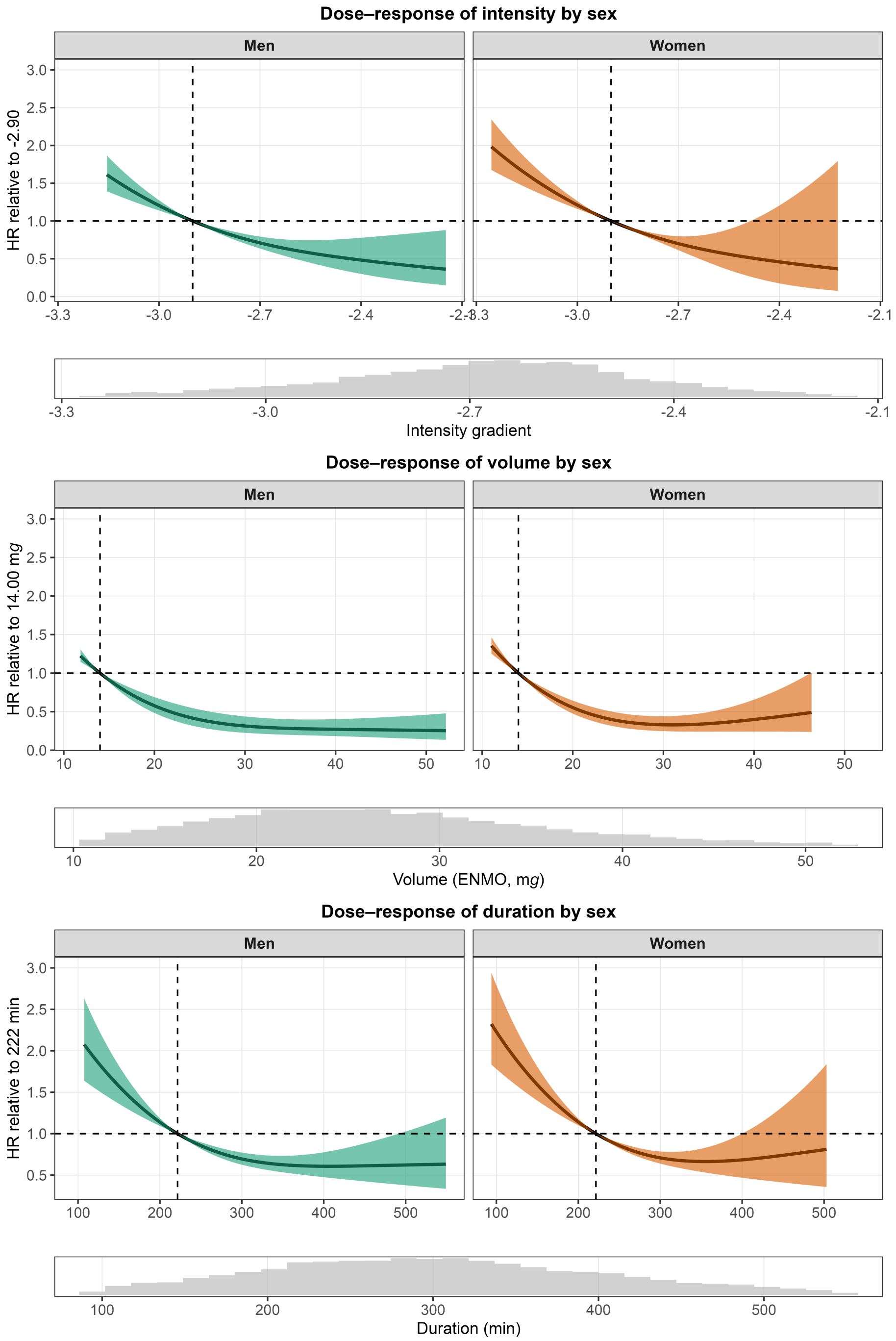

**FIG 40** All-cause mortality: Dose-response relationship between physical activity intensity (top), volume (middle), and duration (bottom) and all-cause mortality risk, predicted for men and women. Models included an interaction term between each physical activity metric and sex. Histograms depict the distribution of the corresponding exposure variable.

##### 8.5.3 Joint association of physical activity and body morphology with all-cause mortality

Figure 41 shows the association between physical activity and all-cause mortality stratified by waist-to-height ratio. Overall, the dose-response curves were largely similar in shape to those observed for CVD mortality but confidence intervals were narrower across all PA dimensions and waist-to-height ratio levels.

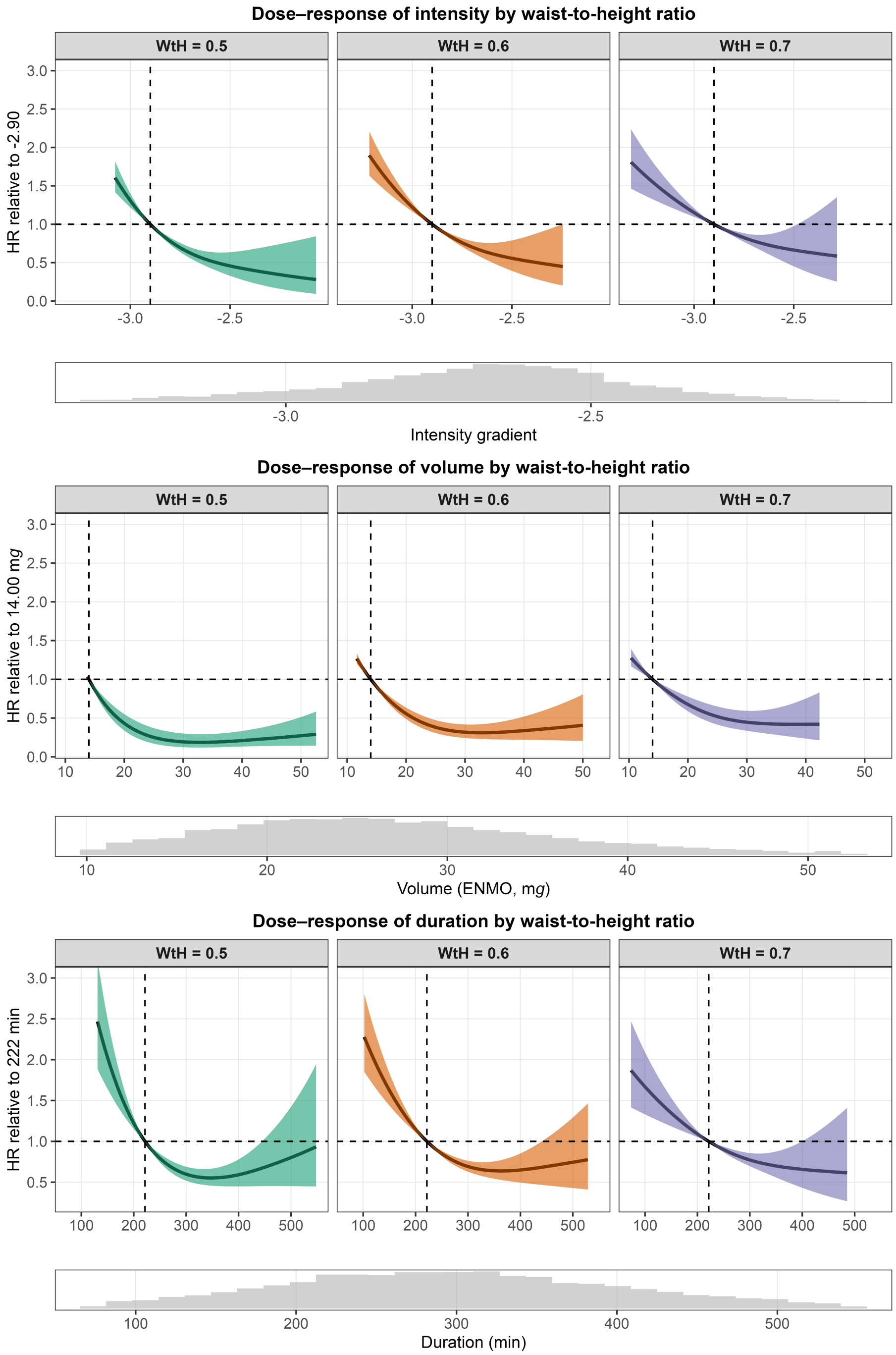

**FIG 41** All-cause mortality: Dose-response relationship between physical activity intensity (top), volume (middle), and duration (bottom) and all-cause mortality risk, predicted for waist-to-height ratio (WtH) 0.5, 0.6, and 0.7. Models included an interaction term between each physical activity metric and WtH. Histograms depict the distribution of the corresponding exposure variable.

Figure 42 shows the results for the interaction between physical activity and waist circumference with all-cause mortality as outcome. We observed similar dose-response shapes across all PA dimensions and waist circumference levels.

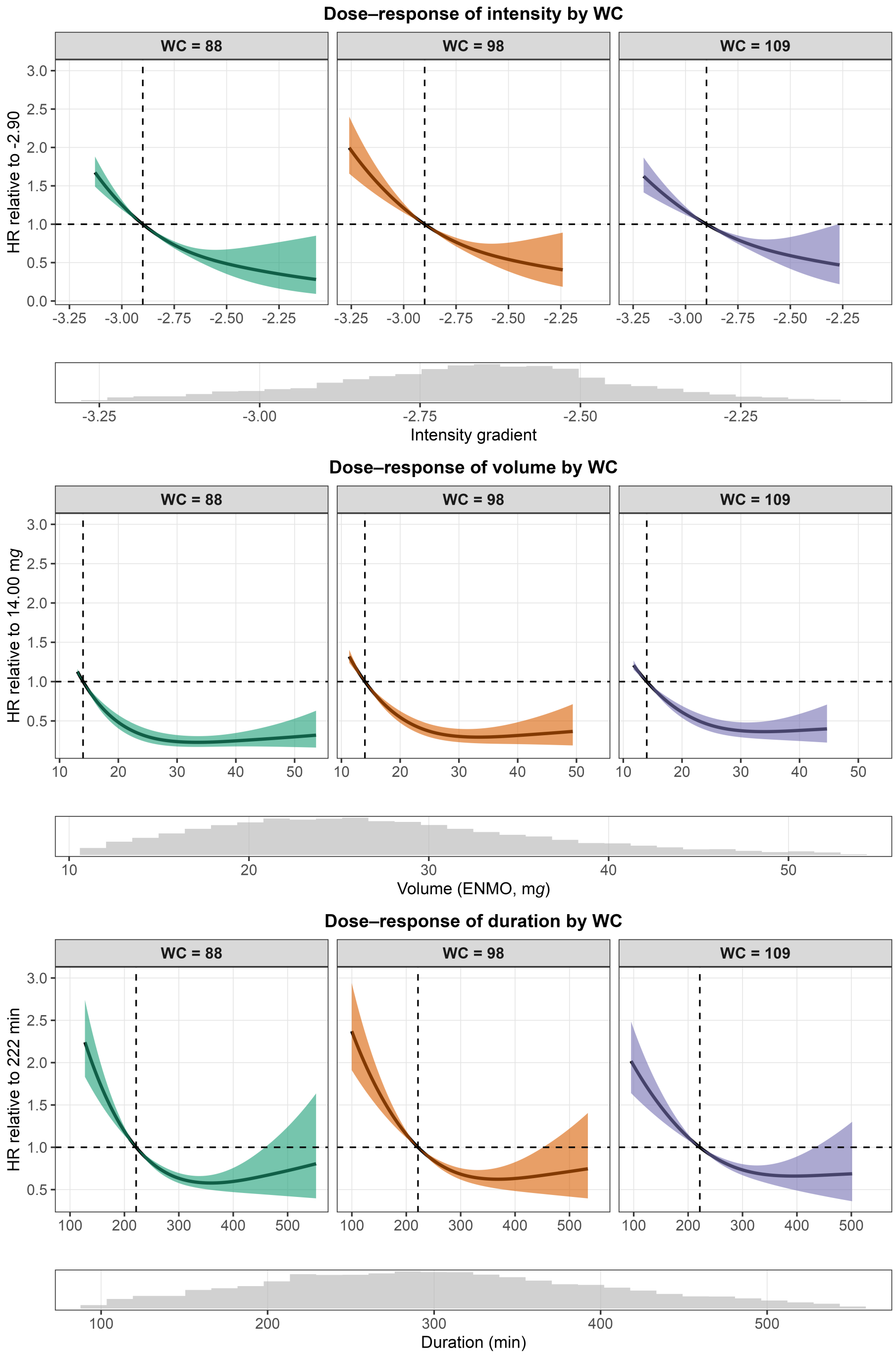

**FIG 42** All-cause mortality: Dose-response relationship between physical activity intensity (top), volume (middle), and duration (bottom) and all-cause mortality risk, predicted for waist circumferences (WC) 88, 98, and 109 cm. Models included an interaction term between each physical activity metric and waist circumference. Histograms depict the distribution of the corresponding exposure variable.

Figure 43 shows the results for the interaction between physical activity and body mass index with all-cause mortality as outcome. Dose-response curves were curvilinear rather than linear for intensity and more certain compared to the main analyses with CVD mortality as outcome. For volume and duration, dose-response shapes were comparable to the main models but with narrower confidence intervals. Yet, a widening of confidence intervals with increasing BMI was still visible.

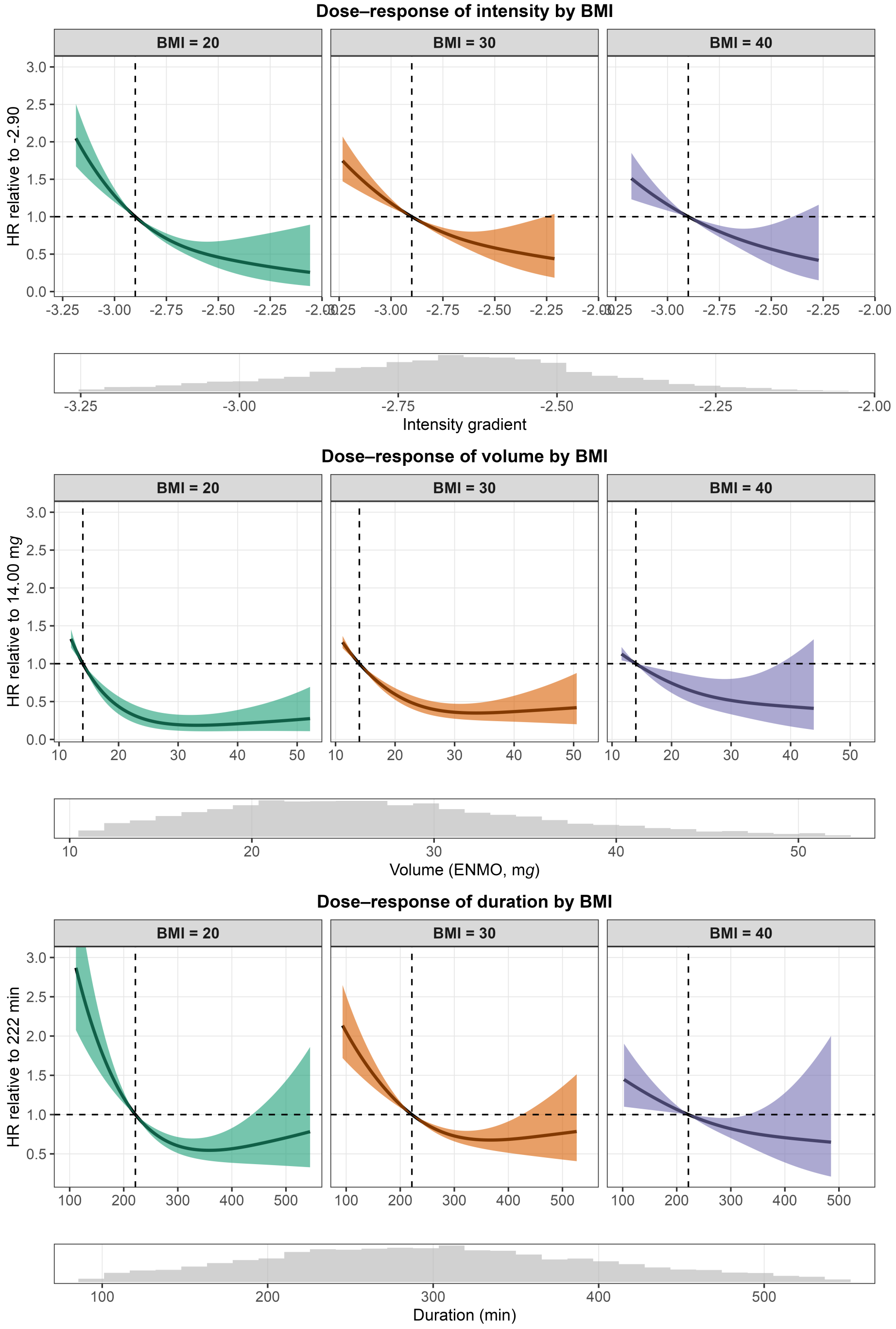

**FIG 43** All-cause mortality: Dose-response relationship between physical activity intensity (top), volume (middle), and duration (bottom) and all-cause mortality risk, predicted for body mass index (BMI) 20, 30, and 40 kg/$m^2$. Models included an interaction term between each physical activity metric and BMI. Histograms depict the distribution of the corresponding exposure variable.

### 9 GGIR configuration

Table 8 lists the GGIR settings used to process the accelerometer data (`config.csv`).

**TABLE 8** GGIR configuration.

| Parameter group | Argument | Value |
|---|---|---|
| not applicable | GGIRread_version | 1.0.10 |
| not applicable | GGIRversion | 3.3.9 |
| not applicable | LC_TIME_backup | en_US.UTF-8 |
| not applicable | R_version | R version 4.6.1 (2026-06-24) |
| not applicable | config_file_in_outputdir | /Volumes/NHANES/output_GGIR/output_NHANES/config.csv |
| not applicable | datadir | /Volumes/NHANES/data |
| not applicable | do.report | 2 |
| not applicable | f0 | 1 |
| not applicable | f1 | 9504 |
| not applicable | mode | 2 |
| not applicable | outputdir | /Volumes/NHANES/output_GGIR |
| not applicable | studyname | NHANES |
| params_247 | IVIS.activity.metric | 2 |
| params_247 | IVIS_acc_threshold | 20 |
| params_247 | IVIS_epochsize_seconds | c() |
| params_247 | IVIS_windowsize_minutes | 60 |
| params_247 | L5M5window | c(0, 24) |
| params_247 | LUX_cal_constant | c() |
| params_247 | LUX_cal_exponent | c() |
| params_247 | LUX_day_segments | c() |
| params_247 | LUXthresholds | c(0, 100, 500, 1000, 3000, 5000, 10000) |
| params_247 | M5L5res | 1 |
| params_247 | MX.ig.min.dur | 10 |
| params_247 | SRI2_WASOmin | 30 |
| params_247 | clevels | c(30, 150) |
| params_247 | cosinor | FALSE |
| params_247 | iglevels | c(0, 25, 50, 75, 100, 125, 150, 175, 200, 225, 250, 275, 300, 325, 350, 375, 400, 425, 450, 475, 500, 525, 550, 575, 600, 625, 650, 675, 700, 725, 750, 775, 800, 825, 850, 875, 900, 925, 950, 975, 1000, 1025, 1050, 1075, 1100, 1125, 1150, 1175, 1200, 1225, 1250, 1275, 1300, 1325, 1350, 1375, 1400, 1425, 1450, 1475, 1500, 1525, 1550, 1575, 1600, 1625, 1650, 1675, 1700, 1725, 1750, 1775, 1800, 1825, 1850, 1875, 1900, 1925, 1950, 1975, 2000, 2025, 2050, 2075, 2100, 2125, 2150, 2175, 2200, 2225, 2250, 2275, 2300, 2325, 2350, 2375, 2400, 2425, 2450, 2475, 2500, 2525, 2550, 2575, 2600, 2625, 2650, 2675, 2700, 2725, 2750, 2775, 2800, 2825, 2850, 2875, 2900, 2925, 2950, 2975, 3000, 3025, 3050, 3075, 3100, 3125, 3150, 3175, 3200, 3225, 3250, 3275, 3300, 3325, 3350, 3375, 3400, 3425, 3450, 3475, 3500, 3525, 3550, 3575, 3600, 3625, 3650, 3675, 3700, 3725, 3750, 3775, 3800, 3825, 3850, 3875, 3900, 3925, 3950, 3975, 4000, 8000) |
| params_247 | ilevels | c(0, 1, 2, 3, 4, 5, 6, 7, 8, 9, 10, 15, 20, 25, 30, 35, 40, 45, 50, 55, 60, 65, 70, 75, 80, 85, 90, 95, 100, 125, 150, 175, 200, 225, 250, 275, 300, 325, 350, 375, 400, 425, 450, 475, 500, 525, 550, 575, 600, 625, 650, 675, 700, 725, 750, 775, 800, 825, 850, 875, 900, 925, 950, 975, 1000, 1100, 1200, 1300, 1400, 1500, 1600, 1700, 1800, 1900, 2000, 8000) |
| params_247 | part2CR | FALSE |
| params_247 | part6CR | FALSE |
| params_247 | part6DFA | FALSE |
| params_247 | part6HCA | FALSE |
| params_247 | part6Window | c(start, end) |
| params_247 | qM5L5 | c(0.05, 0.25, 0.5, 0.75, 0.95) |
| params_247 | qlevels | c(0.333333333333333, 0.416666666666667, 0.5, 0.583333333333333, 0.625, 0.666666666666667, 0.75, 0.833333333333333, 0.916666666666667, 0.958333333333333, 0.96875, 0.979166666666667, 0.986111111111111, 0.989583333333333, 0.993055555555556, 0.99375, 0.994444444444444, 0.995138888888889, 0.995833333333333, 0.996527777777778, 0.997222222222222, 0.997916666666667, 0.998611111111111, 0.999305555555556) |
| params_247 | qwindow | c(0, 24) |
| params_247 | qwindow_dateformat | %d-%m-%Y |

*(continued)*

| Parameter group | Argument | Value |
|---|---|---|
| params_247 | winhr | c(16, 14, 12, 10, 9, 8, 6, 5, 4, 2, 1, 0.75, 0.5, 0.333333333333333, 0.25, 0.166666666666667, 0.15, 0.133333333333333, 0.116666666666667, 0.1, 0.0833333333333333, 0.0666666666666667, 0.05, 0.0333333333333333, 0.0166666666666667) |
| params_cleaning | TimeSegments2ZeroFile | c() |
| params_cleaning | data_cleaning_file | c() |
| params_cleaning | data_masking_strategy | 1 |
| params_cleaning | do.imp | TRUE |
| params_cleaning | excludefirst.part4 | FALSE |
| params_cleaning | excludefirstlast | FALSE |
| params_cleaning | excludefirstlast.part5 | FALSE |
| params_cleaning | excludelast.part4 | FALSE |
| params_cleaning | hrs.del.end | 0 |
| params_cleaning | hrs.del.start | 0 |
| params_cleaning | includecrit.part6 | c(0.666666666666667, 0.666666666666667) |
| params_cleaning | includedaycrit | 16 |
| params_cleaning | includedaycrit.part5 | 0.666666666666667 |
| params_cleaning | includenightcrit | 16 |
| params_cleaning | includenightcrit.part5 | 0 |
| params_cleaning | max_calendar_days | 0 |
| params_cleaning | maxdur | 9 |
| params_cleaning | minimum_MM_length.part5 | 23 |
| params_cleaning | ndayswindow | 7 |
| params_cleaning | nonWearEdgeCorrection | TRUE |
| params_cleaning | nonwearFilterWindow | c() |
| params_cleaning | nonwearFiltermaxHours | c() |
| params_cleaning | nonwear_approach | 2023 |
| params_cleaning | segmentDAYSPTcrit.part5 | c(0.9, 0) |
| params_cleaning | segmentWEARcrit.part5 | 0.5 |
| params_cleaning | strategy | 1 |
| params_cleaning | study_dates_dateformat | %d-%m-%Y |
| params_cleaning | study_dates_file | c() |
| params_general | acc.metric | ENMO |
| params_general | configtz | UTC |
| params_general | dataFormat | raw |
| params_general | dayborder | 0 |
| params_general | desiredtz | UTC |
| params_general | do.parallel | TRUE |
| params_general | expand_tail_max_hours | c() |
| params_general | extEpochData_timeformat | %d-%m-%Y %H:%M:%S |
| params_general | idloc | 6 |
| params_general | maxNcores | 17 |
| params_general | maxRecordingInterval | c() |
| params_general | overwrite | TRUE |
| params_general | part5_agg2_60seconds | FALSE |
| params_general | print.filename | TRUE |
| params_general | recordingEndSleepHour | c() |
| params_general | recording_split_ignore_edges | FALSE |
| params_general | recording_split_overlap | 0 |
| params_general | recording_split_timeformat | %d/%m/%Y %H:%M |
| params_general | recording_split_times | c() |
| params_general | sensor.location | wrist |
| params_general | use_trycatch_serial | FALSE |
| params_general | windowsizes | c(5, 900, 3600) |
| params_metrics | actilife_LFE | FALSE |
| params_metrics | do.anglex | FALSE |
| params_metrics | do.angley | FALSE |
| params_metrics | do.anglez | TRUE |
| params_metrics | do.bfen | FALSE |
| params_metrics | do.bfx | FALSE |

*(continued)*

| Parameter group | Argument | Value |
|---|---|---|
| params_metrics | do.bfy | FALSE |
| params_metrics | do.bfz | FALSE |
| params_metrics | do.brondcounts | FALSE |
| params_metrics | do.dev_roll_med_acc_x | FALSE |
| params_metrics | do.dev_roll_med_acc_y | FALSE |
| params_metrics | do.dev_roll_med_acc_z | FALSE |
| params_metrics | do.en | FALSE |
| params_metrics | do.enmo | TRUE |
| params_metrics | do.enmoa | FALSE |
| params_metrics | do.hfen | FALSE |
| params_metrics | do.hfenplus | FALSE |
| params_metrics | do.hfx | FALSE |
| params_metrics | do.hfy | FALSE |
| params_metrics | do.hfz | FALSE |
| params_metrics | do.lfen | FALSE |
| params_metrics | do.lfenmo | FALSE |
| params_metrics | do.lfx | FALSE |
| params_metrics | do.lfy | FALSE |
| params_metrics | do.lfz | FALSE |
| params_metrics | do.mad | FALSE |
| params_metrics | do.neishabouricounts | FALSE |
| params_metrics | do.roll_med_acc_x | FALSE |
| params_metrics | do.roll_med_acc_y | FALSE |
| params_metrics | do.roll_med_acc_z | FALSE |
| params_metrics | do.zcx | FALSE |
| params_metrics | do.zcy | FALSE |
| params_metrics | do.zcz | FALSE |
| params_metrics | hb | 15 |
| params_metrics | lb | 0.2 |
| params_metrics | n | 4 |
| params_metrics | zc.hb | 3 |
| params_metrics | zc.lb | 0.25 |
| params_metrics | zc.order | 2 |
| params_metrics | zc.sb | 0.01 |
| params_metrics | zc.scale | 1 |
| params_output | criterror | 3 |
| params_output | dec_config | . |
| params_output | dec_reports | . |
| params_output | do.part2.pdf | FALSE |
| params_output | do.part2.png | FALSE |
| params_output | do.part3.pdf | FALSE |
| params_output | do.sibreport | TRUE |
| params_output | do.visual | FALSE |
| params_output | dofirstpage | TRUE |
| params_output | epochvalues2csv | FALSE |
| params_output | method_research_vars | c() |
| params_output | old_visualreport | FALSE |
| params_output | outliers.only | FALSE |
| params_output | require_complete_lastnight_part5 | FALSE |
| params_output | save_dashboard_parquet | FALSE |
| params_output | save_ms5raw_format | RData |
| params_output | save_ms5raw_without_invalid | FALSE |
| params_output | save_ms5rawlevels | TRUE |
| params_output | sep_config | , |
| params_output | sep_reports | , |
| params_output | storefolderstructure | FALSE |
| params_output | timewindow | c(MM, WW) |
| params_output | viewingwindow | 1 |
| params_output | visualreport | TRUE |

*(continued)*

| Parameter group | Argument | Value |
|---|---|---|
| params_output | visualreport_focus | day |
| params_output | visualreport_hrsPerRow | 36 |
| params_output | visualreport_validcrit | 0 |
| params_output | visualreport_without_invalid | TRUE |
| params_output | week_weekend_aggregate.part5 | FALSE |
| params_phyact | boutcriter | 0.8 |
| params_phyact | boutcriter.in | 0.9 |
| params_phyact | boutcriter.lig | 0.8 |
| params_phyact | boutcriter.mvpa | 0.8 |
| params_phyact | boutdur.in | c(10, 20, 30) |
| params_phyact | boutdur.lig | c(1, 5, 10) |
| params_phyact | boutdur.mvpa | c(1, 5, 10) |
| params_phyact | frag.metrics | c() |
| params_phyact | mvpadur | c(1, 5, 10) |
| params_phyact | mvpathreshold | c(40, 100, 400) |
| params_phyact | part6_threshold_combi | 40_100_400 |
| params_phyact | threshold.lig | 40 |
| params_phyact | threshold.mod | 100 |
| params_phyact | threshold.vig | 400 |
| params_rawdata | backup.cal.coef | retrieve |
| params_rawdata | chunksize | 1 |
| params_rawdata | do.cal | TRUE |
| params_rawdata | dynrange | 6 |
| params_rawdata | frequency_tol | 0.1 |
| params_rawdata | imputeTimegaps | TRUE |
| params_rawdata | interpolationType | 1 |
| params_rawdata | minimumFileSizeMB | 2 |
| params_rawdata | minloadcrit | 168 |
| params_rawdata | nonwear_range_threshold | 150 |
| params_rawdata | printsummary | FALSE |
| params_rawdata | rmc.bitrate | c() |
| params_rawdata | rmc.check4timegaps | TRUE |
| params_rawdata | rmc.col.acc | c(2, 3, 4) |
| params_rawdata | rmc.col.temp | c() |
| params_rawdata | rmc.col.time | 1 |
| params_rawdata | rmc.col.wear | c() |
| params_rawdata | rmc.configtz | c() |
| params_rawdata | rmc.dec | . |
| params_rawdata | rmc.desiredtz | c() |
| params_rawdata | rmc.doresample | FALSE |
| params_rawdata | rmc.dynamic_range | c() |
| params_rawdata | rmc.firstrow.acc | 1 |
| params_rawdata | rmc.firstrow.header | c() |
| params_rawdata | rmc.format.time | %Y-%m-%d %H:%M:%OS |
| params_rawdata | rmc.header.length | c() |
| params_rawdata | rmc.header.structure | c() |
| params_rawdata | rmc.headername.recordingid | c() |
| params_rawdata | rmc.headername.sf | c() |
| params_rawdata | rmc.headername.sn | c() |
| params_rawdata | rmc.noise | 0.013 |
| params_rawdata | rmc.origin | 1970-01-01 |
| params_rawdata | rmc.scalefactor.acc | 1 |
| params_rawdata | rmc.sf | 80 |
| params_rawdata | rmc.unit.acc | g |
| params_rawdata | rmc.unit.temp | C |
| params_rawdata | rmc.unit.time | UNIXsec |
| params_rawdata | rmc.unsignedbit | TRUE |
| params_rawdata | spherecrit | 0.3 |
| params_sleep | HASIB.algo | vanHees2015 |

*(continued)*

| Parameter group | Argument | Value |
|---|---|---|
| params_sleep | HASPT.algo | HDCZA |
| params_sleep | HASPT.ignore.invalid | FALSE |
| params_sleep | HDCZA_roll_windowsize | 5 |
| params_sleep | HDCZA_threshold | c() |
| params_sleep | HorAngle_threshold | 60 |
| params_sleep | LowAcc_threshold | 0.014 |
| params_sleep | SRI1_smoothing_frac | c() |
| params_sleep | SRI1_smoothing_wsize_hrs | c() |
| params_sleep | Sadeh_axis | NA |
| params_sleep | anglethreshold | 5 |
| params_sleep | colid | 1 |
| params_sleep | coln1 | 2 |
| params_sleep | consider_marker_button | FALSE |
| params_sleep | def.noc.sleep | 1 |
| params_sleep | guider_cor_do | FALSE |
| params_sleep | guider_cor_maxgap_hrs | 2 |
| params_sleep | guider_cor_meme_frac_in | 0.4 |
| params_sleep | guider_cor_meme_frac_out | 0.9 |
| params_sleep | guider_cor_meme_min_dys | 3 |
| params_sleep | guider_cor_meme_min_hrs | 1 |
| params_sleep | guider_cor_min_frac_sib | 0.5 |
| params_sleep | guider_cor_min_hrs | 2 |
| params_sleep | ignorenonwear | TRUE |
| params_sleep | impute_marker_button | FALSE |
| params_sleep | loglocation | c() |
| params_sleep | longitudinal_axis | c() |
| params_sleep | nap_markerbutton_max_distance | 30 |
| params_sleep | nap_markerbutton_method | 0 |
| params_sleep | nap_model | c() |
| params_sleep | nnights | c() |
| params_sleep | oakley_threshold | 20 |
| params_sleep | possible_nap_dur | c() |
| params_sleep | possible_nap_edge_acc | Inf |
| params_sleep | possible_nap_gap | 0 |
| params_sleep | possible_nap_window | c() |
| params_sleep | relyonguider | FALSE |
| params_sleep | sib_must_fully_overlap_with_TimeInBed | c(TRUE, TRUE) |
| params_sleep | sleepefficiency.metric | 1 |
| params_sleep | sleeplogsep | c() |
| params_sleep | sleepwindowType | SPT |
| params_sleep | spt_max_gap_dur | 60 |
| params_sleep | spt_max_gap_ratio | 1 |
| params_sleep | spt_min_block_dur | 30 |
| params_sleep | timethreshold | 5 |

The Quarto template used to create this supplementary material document has been developed by [19] and [20].

205 # References

206 1. Davies NM, Dickson M, Davey Smith G, Windmeijer F, Berg GJ van den. The causal effects of education on adult health, mortality and income: Evidence from mendelian randomization and the raising of the school leaving age. International Journal of Epidemiology. 2023;52:1878–86. doi:10.1093/ije/dyad104

207 2. Stringhini S, Carmeli C, Jokela M, Avendaño M, Muennig P, Guida F, et al. Socioeconomic status and the 25 × 25 risk factors as determinants of premature mortality: A multicohort study and meta-analysis of 1·7 million men and women. The Lancet. 2017;389:1229–37. doi:10.1016/s0140-6736(16)32380-7

208 3. Martinez-Gomez D, Luo M, Huang Y, Rodríguez-Artalejo F, Ekelund U, Sotos-Prieto M, et al. Physical activity and all-cause mortality by age in 4 multinational megacohorts. JAMA Network Open. 2024;7:e2446802. doi:10.1001/jamanetworkopen.2024.46802

209 4. Hamer M, Kivimaki M, Steptoe A. Longitudinal patterns in physical activity and sedentary behaviour from mid-life to early old age: A substudy of the whitehall II cohort. Journal of Epidemiology and Community Health. 2012;66:1110–5. doi:10.1136/jech-2011-200505

210 5. Lantz PM, House JS, Lepkowski JM, Williams DR, Mero RP, Chen J. Socioeconomic factors, health behaviors, and mortality. JAMA. 1998;279:1703. doi:10.1001/jama.279.21.1703

211 6. Wang DD, Li Y, Bhupathiraju SN, Rosner BA, Sun Q, Giovannucci EL, et al. Fruit and vegetable intake and mortality. Circulation. 2021;143:1642–54. doi:10.1161/circulationaha.120.048996

212 7. Ashwell M, Gunn P, Gibson S. Waist-to-height ratio is a better screening tool than waist circumference and BMI for adult cardiometabolic risk factors: Systematic review and meta-analysis. Obesity Reviews. 2011;13:275–86. doi:10.1111/j.1467-789x.2011.00952.x

213 8. Singh N, Areal AT, Breitner S, Zhang S, Agewall S, Schikowski T, et al. Heat and cardiovascular mortality: An epidemiological perspective. Circulation Research. 2024;134:1098–112. doi:10.1161/circresaha.123.323615

214 9. Mornas A, Deshayes TA, Bartlett AA, Chaseling G, Jay O, Marzolini S, et al. A scoping review of recommendations for adults with cardiovascular disease to remain physically active during hot weather. European Journal of Preventive Cardiology. 2025. doi:10.1093/eurjpc/zwaf519

215 10. Migueles JH, Rowlands AV, Huber F, Sabia S, Hees VT van. GGIR: A research community-driven open source r package for generating physical activity and sleep outcomes from multi-day raw accelerometer data. Journal for the Measurement of Physical Behaviour. 2019;2:188–96. doi:10.1123/jmpb.2018-0063

216 11. Rowlands AV, Edwardson CL, Davies MJ, Khunti K, Harrington DM, Yates T. Beyond cut points: Accelerometer metrics that capture the physical activity profile. Medicine & Science in Sports & Exercise. 2018;50:1323–32. doi:10.1249/MSS.0000000000001561

217 12. Harrell Jr FE, Beck C, Dupont C. Hmisc: Harrell miscellaneous [Internet]. 2026. Available from: https://CRAN.R-project.org/package=Hmisc

218 13. Latouche A, Allignol A, Beyersmann J, Labopin M, Fine JP. A competing risks analysis should report results on all cause-specific hazards and cumulative incidence functions. Journal of Clinical Epidemiology. 2013;66:648–53. doi:https://doi.org/10.1016/j.jclinepi.2012.09.017

219 14. Austin PC, Fine JP. Practical recommendations for reporting fine-gray model analyses for competing risk data. Statistics in Medicine. 2017;36:4391–400. doi:https://doi.org/10.1002/sim.7501

220 15. Rao JNK, Scott AJ. On chi-squared tests for multiway contingency tables with cell proportions estimated from survey data. The Annals of Statistics [Internet]. 1984 [cited 2026 Aug 24];12:46–60. Available from: http://www.jstor.org/stable/2241033

221 16. Eekhout I, Wiel MA van de, Heymans MW. Methods for significance testing of categorical covariates in logistic regression models after multiple imputation: Power and applicability analysis. BMC Medical Research Methodology. 2017;17:129. doi:10.1186/s12874-017-0404-7

222 17. Ashwell M, Gunn P, Gibson S. Waist-to-height ratio is a better screening tool than waist circumference and BMI for adult cardiometabolic risk factors: Systematic review and meta-analysis. Obesity Reviews. 2011;13:275–86. doi:10.1111/j.1467-789x.2011.00952.x

223 18. Rowlands AV, Kingsnorth AP, Hansen BH, Fairclough SJ, Boddy LM, Maylor BD, et al. Enhancing clinical and public health interpretation of accelerometer-assessed physical activity with age-referenced values based on UK biobank data. Journal of Sport and Health Science. 2025;14:100977. doi:https://doi.org/10.1016/j.jshs.2024.100977

224 19. Overleaf. LaTeX template for research and review papers in ASM journals [Internet]. 2025. Available from: https://www.overleaf.com/latex/templates/latex-template-for-research-and-review-papers-in-asm-journals/kvdbthwnzcwn

225 20. Sovacool K. Kelly-sovacool/quarto-manuscript [Internet]. GitHub repository; 2025. Available from: https://github.com/kelly-sovacool/quarto-manuscript
